# Latent subtypes of manic or irritable episode symptoms in two population-based cohorts

**DOI:** 10.1101/2021.04.14.21255394

**Authors:** Ryan Arathimos, Chiara Fabbri, Evangelos Vassos, Katrina A S Davis, Oliver Pain, Alexandra Gillett, Jonathan R I Coleman, Ken Hanscombe, Saskia Hagenaars, Bradley Jermy, Anne Corbett, Clive Ballard, Dag Aarsland, Byron Creese, Cathryn M Lewis

## Abstract

**Background:** Episodic changes in mood characterise disorders such as bipolar disorder, which includes distinct periods of manic excitability or irritability, along with additional symptoms experienced during these periods. Common clinical understanding informs diagnostic criteria and epidemiological studies reflect clinical thresholds.

**Aims:** To use a data-driven approach to defining groupings of symptoms experienced during periods of manic or irritable mood, which could inform understanding of mood disorders and guide case classification by identifying subgroups with homogeneous clinical/functional outcomes.

**Methods:** We used latent class analysis (LCA) to conduct an exploration of the latent structure in symptom responses in the UK Biobank and PROTECT studies, by investigating how symptoms, experienced during periods of manic or irritable mood, formed latent subgroups. We tested associations of latent subgroups with sociodemographic characteristics, diagnoses of psychiatric disorders and polygenic risk scores (PRS).

**Results:** Five latent classes were identified that captured patterns of symptoms experienced during periods of manic or irritable mood (N=42,183) in UK Biobank. We identified one class that experienced disruptive episodes of mostly irritable mood that was largely comprised of cases of depression/anxiety, and a class of individuals with increased confidence/creativity that reported lower disruptiveness and lower functional impairment. The five latent classes were replicated in an independent cohort, the PROTECT study (N=4,445), with similar distinctions between classes.

**Conclusion:** Our data-driven approach to grouping individuals identified distinct latent classes. A dimensional classification of mood disorders informed by our findings will be able to better assess or subtype these disorders in future studies.

## Introduction

### Background

Mood disorders are common in the general population (1,2) and lead to significant impairment in the individual, as well as direct and indirect costs to society (3). The episodic nature and intra-individual symptom heterogeneity of these conditions can make diagnosis based on subjective symptom reports challenging(4). All mood disorders are characterised by changes in mood according to ICD-10(5) and DSM-5 (6) diagnostic criteria. DSM-5 specifies that bipolar disorder diagnosis requires a distinct period of abnormally and persistently elevated, euphoric, or irritable mood must occur, in the presence of a specified number of additional symptoms and usually some degree of impairment. The additional symptoms in DSM-5 encompass: 1)inflated self-esteem or grandiosity, 2)decreased need for sleep, 3)increased talkativeness, 4)racing thoughts, 5)being easily distracted, 6)increased goal-directed activity or psychomotor agitation, and 7)engagement in activities that hold the potential for painful consequences (6). Bipolar disorder type I and type II are differentiated by the presence of mania in type I, compared to hypomania (a state with many of the symptoms of mania but milder, with less disruption to life) in type II.

### Data-driven classifications

Epidemiological studies of bipolar spectrum disorders use questionnaires to ascertain symptoms, with various approaches proposed (7–9). In the UK Biobank (10), questions based on DSM criteria were used to assess presence and severity of symptoms (11)(12), with responses used to assess potential case status for mood disorders. Whereas both diagnostic and epidemiological classifications reflect common clinical understanding of mood disorders, they do not usually make use of more data-driven approaches to justify such classifications. Further explorations of mental health definitions could aid epidemiological studies to refine the cases into more homogenous groups for investigation(13). Precise phenotypes (or disease endotypes) will be instrumental in the shift to precision medicine and patient-specific tailored treatments, based on a more data-centric approach to disease taxonomy, with various frameworks and solutions already proposed (14–16).

Latent class analysis (LCA) is a model-based probabilistic method of identifying homogenous groups (classes) based on patterns present in a set of categorical indicator variables. Previous studies have used this data-driven approach to identify subtypes of disease based on symptoms data. A general-population study of both manic/irritable and psychotic episode symptoms (n=1846) identified five classes differentiated by the presence of irritability and psychotic experiences, as well as differential associations with sociodemographic and clinical characteristics (17). Other clustering methods have also been employed to inform data-driven distinctions between mood disorders, such as with longitudinal patterns of mood to identify individuals with bipolar disorder type I (18). Previous studies conducting LCA of symptoms have often lacked replication in external datasets or been performed in small samples.

### Aims

In this study, we conducted a data-driven exploratory analysis of latent structure in reported symptoms experienced during manic or irritable episodes. Our aims were two-fold: 1) to identify latent classes with homogeneous clinical characteristics and functional outcomes that may have clinical or biological relevance independent of diagnostic categories; 2) to investigate the correspondence of latent classes with reported psychiatric diagnoses and genetic liability, in order to aid in refining commonly used epidemiological definitions of probable bipolar disorder.

## Methods

### Study populations

#### UK Biobank

Study participants for the discovery analysis were drawn from the UK Biobank. Briefly, the UK Biobank is a prospective cohort study of over 500,000 individuals across the UK. Participants were aged 40-69 years at recruitment in 2006–2010 (10). Genotype data was available for all UK Biobank participants (19). Ethical approval was provided by the Research Ethics Committee (REC reference 11/NW/0382). In a follow-up, participants who had agreed to be recontacted were invited to complete an online mental health questionnaire (MHQ) in 2017, resulting in additional phenotypic data in 157,366 UKB participants (12).

### Phenotype data

To characterise probable history of mood disorders, UK Biobank included questions based on the approach used in the Structured Clinical Interview for DSM-IV Axis I Disorders (SCID-I) at baseline and in the MHQ (see **Supplementary Methods**). Participants answered questions on ever having experienced a manic/irritable episode, as in **Box 1**.

#### Box 1

*MHQ questions*

Period of manic/hyper mood (field #20501)

*“Have you ever had a period of time when you were feeling so good*, “*high”*, “*excited", or* “*hyper” that other people thought you were not your normal self or you were so* “*hyper” that you got into trouble?*”

Period of irritable mood (field #20502)

*“Have you ever had a period of time when you were so irritable that you found yourself shouting at people or starting fights or arguments?”*.

Participants that answered positively to either or both above questions were asked about symptoms experienced during these episodes (field #20548), selecting all that might apply

*Please try to remember a period when you were in a* “*high” or* “*irritable” state and select all of the following that apply:*

- *I was more talkative than usual*,
- *I was more restless than usual, my thoughts were racing*,
- *I needed less sleep than usual*,
- *My thoughts were racing*,
- *I was more creative or had more ideas than usual*,
- *I was easily distracted*,
- *I was more confident than usual*,
- *I was more active than usual*

Participants who answered positive to above fields were then asked about:

- The longest **duration** of any such episode (field #20492): brief (<24 hours); moderate (>24 hours but <1 week); or extended (>1 week).
- The **disruptiveness** of the episode (field #20493): not disruptive or disruptive if participants reported that the episode required treatment, caused problems with work,

Sociodemographic data on participant sex, age, smoking status, alcohol intake frequency, Townsend deprivation index (TDI, a measure of area-level deprivation as a proxy for socioeconomic status) and education level were extracted from participant responses to the baseline questionnaire (see **Supplementary Methods)**.

In the MHQ, participants reported past diagnoses by a professional (field #20544) of several disorders, which were used to define six broad diagnostic categories; attention deficit hyperactivity disorder (ADHD), autism spectrum disorder (ASD), generalised anxiety disorder (GAD), depression, schizophrenia/psychosis, and mania/bipolar disorder (see **Supplementary Methods**).

Linked electronic health records to Hospital Episode Statistics (HES), which contain hospital diagnoses recorded with the International Classification of Diseases, 10th Revision (ICD-10) up until June 2020, were used to derive cases status for 3 broad disorder definitions; depression, schizophrenia/psychotic disorder, and mania/bipolar disorder (see **Supplementary Methods**).

### Polygenic risk scores (PRS)

Genetic data pre-processing and sample exclusions are described in **Supplementary Methods**. We calculated PRS using PRSice v2 (20,21), with clumping (r2< 0.1 and 500kb window) and a p-value threshold of 1 (all SNPs included) for all analyses. PRS were residualized for the first 6 genetic principal components (PCs) and scaled to a mean of zero and standard deviation of 1. Summary results from GWAS of anxiety disorder(22), ADHD(23), ASD(24), major depression(25), bipolar disorder(26) and schizophrenia(27) were used, from studies that did not include UK Biobank participants in the discovery sample (**Table S1**).

#### PROTECT – Replication sample

We attempted replication of findings in the Platform for Research Online to Investigate the Genetics and Cognition in Aging (PROTECT) study. Briefly, the PROTECT study is a UK-based online participant registry with continuous, ongoing recruitment beginning in 2015 which tracks the cognitive health of older adults. Study participants must be >50 years old, have no diagnosis of dementia, and must have access to a computer/internet. Beginning in 2015, 14,836 PROTECT study participants were invited to complete the same online MHQ as UK Biobank participants, as a pilot of the questionnaire before roll-out in the UK Biobank.

### Phenotype data

Participant responses to the MHQ in PROTECT were extracted from baseline questionnaires using the same derivation process as UK Biobank. Sociodemographic variables on sex, age, smoking, education level and alcohol consumption frequency were derived from responses to baseline questionnaires.

### Genetic data

A subset of PROTECT study participants provided a saliva sample for genotyping. Genetic data pre-processing and sample exclusions are described in **Supplementary Methods**. The total number of individuals with genetic data following exclusions was 8,272. PRS were calculated as for UK Biobank, residualized for the first 6 genetic PCs and rescaled.

### Statistical analysis

#### Latent class analysis

Latent class analysis (LCA) is a model-based method that estimates the distribution of an underlying unobserved categorical variable, hypothesised to explain the patterns of association between a set of discrete variables. The estimated categorical variable describes “classes” or subgroups of individuals within the data. The method estimates the posterior probabilities of an individual belonging to a particular latent class. LCA was run using the poLCA package(28) in R, which uses the maximum likelihood method. Models with increasing numbers of classes, beginning at 2 and up to 7, were compared for best fit using the Bayesian Information Criterion (BIC) and Akaike Information Criterion (AIC). The relative entropy (a measure of classification certainty ranging between 0 and 1) was used to assess separation between classes(29).

#### Multinomial logistic regression

Multinomial logistic regression was used to test for association between class membership, as the outcome (based on most likely class membership probability) and sociodemographic variables, disorder diagnoses (self-reported or hospital) and polygenic risk scores. Posterior probabilities of class membership were used as weights. Relative risk ratios (RR) were estimated for each class, compared to a reference class. For categorical variables (education attainment, smoking and alcohol consumption), dummy coding was used for each level, with the reference level of each being all combined remaining levels.

## Results

In the UK Biobank MHQ, 42,183 participants responded positively to the questions on a manic or irritable episode and completed the episode symptoms questions (**Table S2**). Characteristics of this analytical subset and all MHQ respondents are shown in **Table 1**.

**Table 1.** Comparison of sociodemographic characteristics in the subset of participants in the latent class analysis (LCA subset) with responses to the stem question on ever having experienced a period of manic or irritable mood as well as the subsequent questions of symptoms, and the participants who completed the Mental Health Questionnaire (MHQ - whole sample) in the UK Biobank.

|  |  | LCA subset |  | Full MHQ sample |  |
| --- | --- | --- | --- | --- | --- |
|  |  | N | percent | N | percent |
| **Total** |  | 42183 | - | 151159 | - |
| **Sex** | Female | 24402 | 58 | 85557 | 57 |
|  | Male | 17779 | 42 | 65597 | 43 |
|  | NA/Missing | 2 | 0.005 | 5 | 0.003 |
| **Education** | University degree | 18820 | 45 | 68467 | 45 |
|  | A levels NVQ HNC or HND | 14411 | 34 | 49677 | 33 |
|  | O levels or CSE | 5901 | 14 | 21169 | 14 |
|  | None | 2654 | 6.3 | 10424 | 6.9 |
|  | NA/ Missing | 397 | 0.94 | 1422 | 0.94 |
| **Alcohol consumption** | Daily | 9245 | 22 | 35176 | 23 |
|  | Weekly | 20973 | 50 | 76917 | 51 |
|  | Occasionally | 9371 | 22 | 30484 | 20 |
|  | Never | 2551 | 6 | 8459 | 5.6 |
|  | NA/ Missing | 43 | 0.1 | 123 | 0.081 |
| **Smoking** | Current | 2877 | 6.8 | 7303 | 4.8 |
|  | Past | 17124 | 41 | 56483 | 37 |
|  | Never | 22076 | 52 | 87037 | 58 |
|  | NA/ Missing | 106 | 0.25 | 336 | 0.22 |
|  |  | **Mean** | **SD** | **Mean** | **SD** |
| **TDI\*** |  | -1.4 | 3.0 | -1.7 | 2.8 |
|  | NA/ Missing | 78 |  | 196 |  |
| **Age** |  | 53.9 | 7.8 | 55.9 | 7.7 |
|  | NA/ Missing | 2 |  | 5 |  |

\*TDI – Townsend Deprivation Index values before rank normalisation (high values = increased deprivation).

### Latent class analysis

LCA was applied to the eight binary symptoms, as indicators, in the subset of participants reporting a manic and/or irritable episode (N=42,183). As the number of classes increased, BIC and AIC both continuously decreased, with no minimum attained (**Table S3**). Elbow/scree plots (30) (**Figure S1**) indicated that either a 4 or 5-class model was the optimum model. Plotting the conditional probabilities for each indicator symptom showed that the additional class in the 5-class model was distinct from the other 4 (**Figure S2**). We therefore selected the 5-class model as the optimum model.

The conditional probabilities of the eight indicator symptoms in each of the five latent classes are shown in **Figure 1A**. Individuals in the first class (3.2%) had a high probability of reporting all symptoms and was therefore labelled the “extensively affected” (EA) class. The second class (9.8%) was labelled “focused creative” (FC), as individuals reported being more active, talkative, confident and creative. Individuals in the third class (11.5%), had high probabilities of being more active, talkative, restless, easily distracted and having racing thoughts. This class was labelled the “active restless” (AR). Individuals in the fourth class (31.6% of the sample) had a high probability of reporting racing thoughts, feeling more restless and being more easily distracted. This class was labelled the “inactive restless” (IR). The fifth class (43.9%) had low probabilities of reporting all symptoms and was therefore labelled the “minimally affected” (MA) and was used as the reference class in downstream analyses.

**Figure 1.**
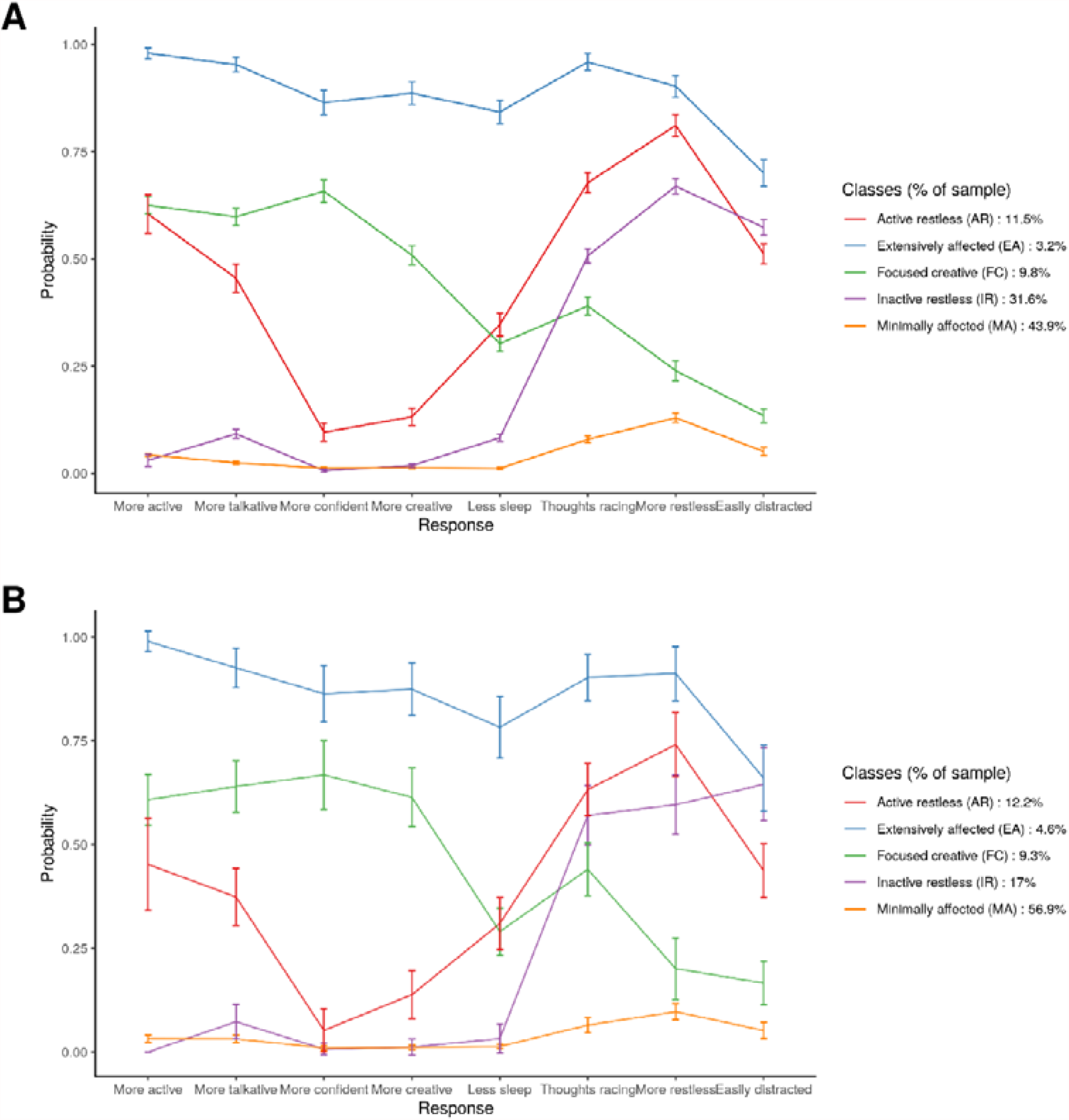
Conditional probabilities of each response (symptom) in, (A) the UK Biobank optimum 5-class latent model (N= 42,183), (B) the PROTECT replication study optimum 5-class latent model (N=4,445).

Distributions of responses to the original stem question of ever experiencing a period of manic and/or irritable mood by most likely class membership indicated that the inactive restless and the minimally affected were comprised mostly of individuals reporting an irritable episode. The active restless was comprised of individuals reporting an irritable episode and (to a lesser extent) both a manic and an irritable episode, whereas the focused creative was comprised of individuals reporting an irritable, a manic, or both a manic and an irritable episode. The extensively affected was comprised mostly of individuals reporting both a manic and an irritable episode. (**Figure 2, Figure S3, Table S4**)

**Figure 2.**
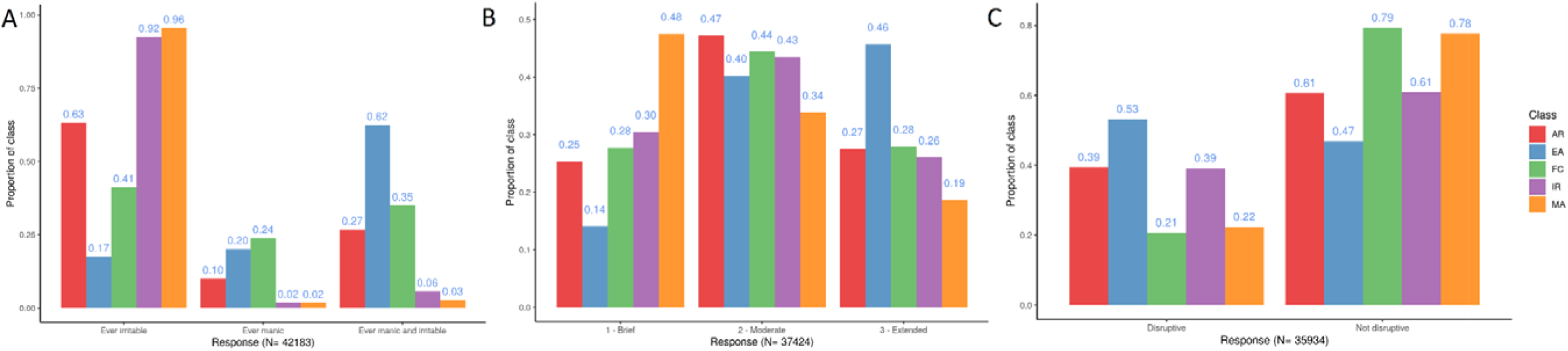
Distributions of responses to (A) the stem question on ever experiencing a period of manic or irritable mood, (B) the subsequent question on episode duration in a subset of N=37,424 participants with available data, and (C) the subsequent question on episode disruptiveness in a subset of N=35,934 participants with available data, by most likely class membership, in the optimum 5-class model. Classes are AR = active restless class, EA = extensively affected class, FC = focused creative class, IR = inactive restless class, MA = minimally affected class.

### Associations with episode duration and disruptiveness

For responses to episode duration (N=37,424; brief, moderate or extended duration), individuals in the minimally affected were more likely to report brief duration, while those in the extensively affected mostly reported extended duration (**Figure 2(B)**). Episode duration patterns did not differ among the remaining three classes substantially. Associations of episode duration with each class when using the minimally affected as reference largely reflected the observations from the most likely class membership (**Figure S5, Table S5-S6**).

Episode disruptiveness (N=35,934) showed a similar pattern to duration, with the highest proportion of reported disruption in the extensively affected (53%) and lowest in the focused creative (21%) and minimally affected (22%) (**Figure 2(C)** and **Table S7**). Individuals reporting disruptive episodes were more likely to be in the inactive restless and the active restless, and far more likely to be in the extensively affected (**Figure S7, Table S8**). Notably, levels of non-response to the questions on episode duration and disruptiveness were high (N=4,759 and N=6,249 respectively, **Figures S4, S6)**.

### Associations with sociodemographic characteristics

Associations with sociodemographic characteristics were investigated in a subset of N=41,620 individuals (**Tables S9-S17, Figures S8-S16**). Being male was associated with an increased risk of being in all classes when compared to the minimally affected, with a particularly high risk for the focused creative. Higher educational attainment was associated with increased risk of being in the extensively affected and the focused creative. For alcohol intake, individuals in the extensively affected and active restless were less likely to drink, whereas those in the focused creative were more likely to drink daily. There was an increased risk of current smoking for the extensively affected and a smaller increase for remaining classes. For TDI, there was an increased risk of being in the extensively affected with increasing TDI score (increased deprivation) and smaller but significant increases in risk for the other classes, when compared to the minimally affected.

### Associations with self-reported diagnoses of psychiatric disorders

The self-reported diagnoses of six psychiatric disorders differed substantially between the latent classes (N=42,183). Over half of individuals (54.9%) did not report a diagnosis of any of the self-reported disorders studied: ADHD, GAD, ASD, mania/bipolar disorder, depression and schizophrenia/psychosis. Amongst those that did report one or more diagnoses (**Figure S17**), individuals reporting a diagnosis of either depression or GAD (or a combination of both) were the most numerous and were mostly members of either the minimally affected or the inactive restless. Individuals with a diagnosis of mania/bipolar disorder, either alone or in combination with one or more of the remaining disorders, were mostly members of the extensively affected. Diagnosis of any of the six disorders was associated with increased risk of being in the extensively affected (**Figure 3(A)**), with the highest increases in risk observed for mania/bipolar disorder and schizophrenia/psychosis. Diagnosis of depression and GAD was associated with increased risk of being in the inactive restless, with weaker evidence for increased risk of being in this class for mania/bipolar disorder and schizophrenia/psychosis. Diagnosis of all six disorders was associated with increased risk of being in the focused creative and the active restless, with the strongest associations for each class observed for mania/bipolar disorder (**Figures S18, Tables S18-20**).

**Figure 3.**
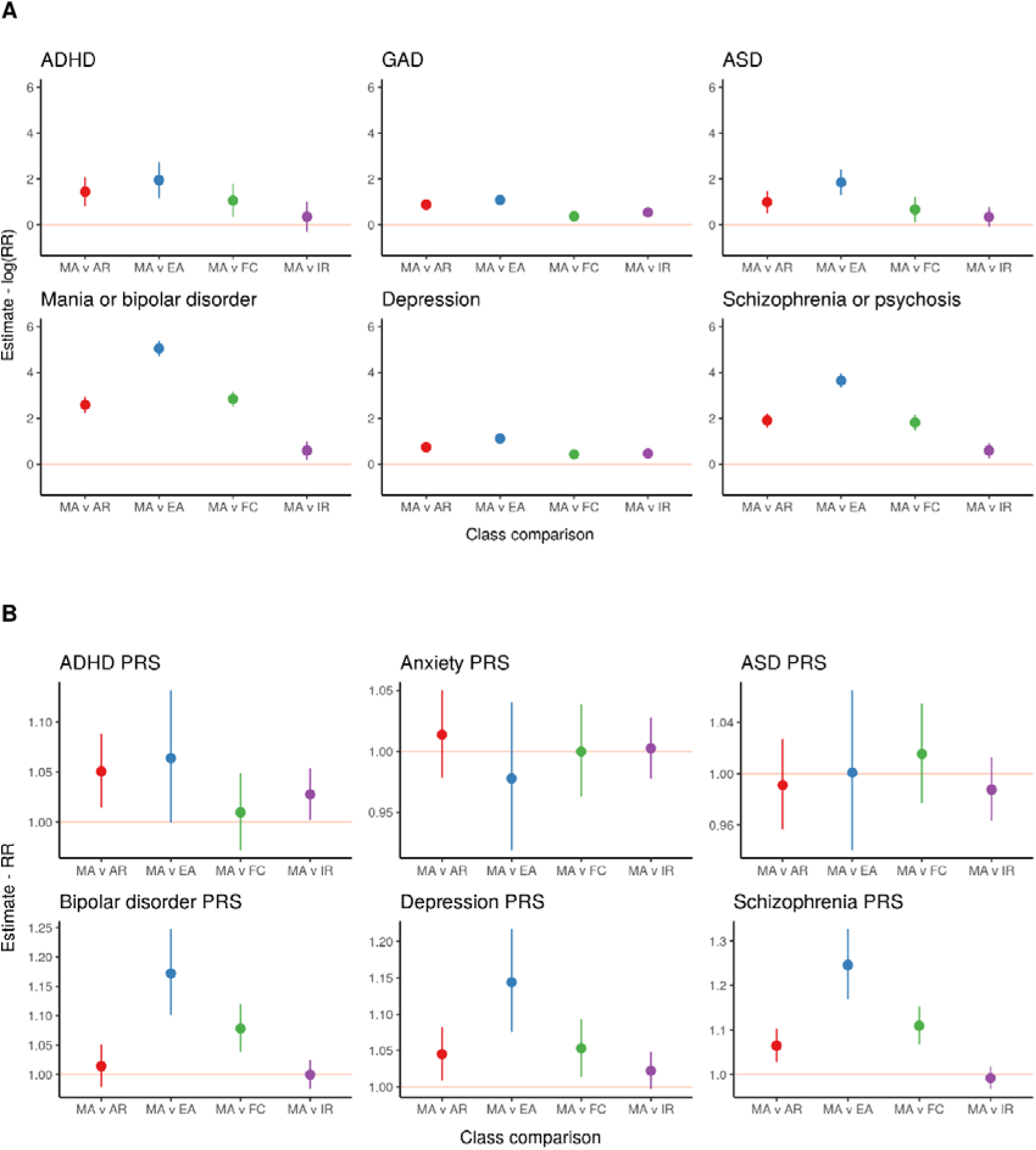
(A) Associations of self-reported diagnoses of six disorders with most likely class membership, weighted for the probability of inclusion of an individual in that class. Effect estimates are presented as natural log risk ratio (RR) of inclusion in each class (relative to the reference class) for cases of each disorder. (B) Associations of polygenic risk scores (PRS) of six disorders with most likely class membership in a subset of N=33,604 with genetic data, weighted for the probability of inclusion of an individual in that class. Effect estimates are presented as risk ratios (RR) of inclusion in each class (relative to the reference class) per standard deviation (SD) increase in standardised polygenic risk score for each disorder. The “minimally affected” class is used as the reference (comparison) class in all analyses. Classes are AR = active restless class, EA = extensively affected class, FC = focused creative class, IR = inactive restless class, MA = minimally affected class.

Observed differences between classes when examining ICD-10 diagnoses of depression, mania/bipolar disorder and schizophrenia/psychotic disorder extracted from hospital records (N=36,258) largely corroborated findings of the analysis of self-reported diagnoses. However, the number of cases of hospital diagnoses for all three disorders was low (**Figures S24-25, Tables S21-23**).

We also explored associations with personality traits (personality disorder diagnosis and neuroticism score) and overlap of each class with cases of probable bipolar disorder type I and II, as defined by Davis et al. and Smith et al. (**Supplementary Results Section A, Tables S24-25, Figures S19-23)**.

### Associations with PRS of psychiatric disorders

Polygenic risk scores (PRS) of psychiatric traits discriminated between classes (N=33,604) (**Figure 3(B)** and **Tables S26-S27**). PRS of schizophrenia was associated with increased risk of being in the extensively affected, the focused creative and the active restless classes. For bipolar disorder PRS there was an increased risk of being in the extensively affected and the focused creative. Depression PRS conferred an increased risk of being in the extensively affected, the focused creative and the active restless. Results for ADHD were weaker, with an increased risk of being in the active restless and to a lesser degree, the inactive restless. Anxiety and ASD showed no significant increase in observed risk of being in any of the classes. These results contrast with the high proportion of GAD and ASD diagnoses reported by the extensively affected, but might also reflect lower power of the PRS, which explains only a fraction of the variance in each trait.

### Replication

#### Latent class analysis

In the PROTECT replication cohort, there were N=4,445 participants with positive responses to the questions on ever experiencing a manic or irritable episode, ∼10% of the sample size of UK Biobank (**Tables S28**-**S29)**.

Comparing latent class models with increasing numbers of classes; indicated that a 5-class model was again the optimum model, with an almost identical patterns of condition probabilities for the symptom indicators (**Figure 1B, Table S30**). The size of some classes was notably different to the discovery cohort (31.6% vs 17% for the inactive restless and 43.9% vs 56.9% for the minimally affected). Distributions of responses to the stem question of ever experiencing a period of manic and/or irritable mood were also similar to the discovery results. The inactive restless and minimally affected comprised mostly of individuals reporting an irritable episode, whereas the extensively affected was comprised mostly of both manic and irritable episodes. The focused creative and active restless were more mixed (**Figure S26, Table S31**).

#### Associations of latent classes in PROTECT

Similar associations to the discovery analyses were found between episode duration (N=3,706) and episode disruptiveness (N=3,290) with the five latent classes in PROTECT (**Figures S27-S30, Table S32-35**). Associations with sociodemographic characteristics (N=4,411) suggested similar distinctions between classes to the discovery analyses, although associations were often weaker and of smaller magnitude (**Figure S31-38, Table S36-39**). For self-reported diagnoses of disorders (N=4421), there were an adequate number of cases (n>20) to analyse four disorders; depression, schizophrenia/psychosis, mania/bipolar disorder and GAD (**Figures S39-42**). There was increased risk of being in all classes with a diagnosis of depression or GAD that mirrored the associations found in the discovery analysis. A diagnosis of schizophrenia/psychosis or mania/bipolar disorder led to an increased risk of being in the extensively affected in particular (**Figures S43-46, Table S40**). For PRS of six disorders (N=1494), directions of effect were mostly consistent with the discovery cohort but with confidence intervals that overlapped the null, except for an increased risk of being in the extensively affected for bipolar disorder PRS (**Figure S47, Table S41-42**).

## Discussion

Using questions designed to discriminate participants with bipolar disorder, we have identified latent structure in participants reporting symptoms experienced during periods of manic or irritable mood. In both the main discovery cohort and the replication cohort, the experiences of these participants fall into five latent classes, with membership associated with episode duration, episode disruptiveness, sociodemographics, diagnoses of psychiatric disorders, and genetic risk of those disorders. These classes likely encompass a broad range of disorders, sub-diagnostic threshold conditions and non-pathological experiences.

The extensively affected class comprises individuals who are the most markedly clinically affected, with particular enrichment for diagnosis of bipolar disorder and schizophrenia but also including cases of depression, anxiety, ADHD and ASD. The inactive restless class comprises individuals endorsing aspects of depression and anxiety, but not schizophrenia, bipolar disorder, ADHD or ASD. The active restless class encompasses aspects of all disorders, to a lesser extent than the extensively affected class. The focused creative class comprises individuals with aspects of mostly bipolar disorder and schizophrenia, and to a lesser extent than the inactive restless class, anxiety, depression and ADHD/ASD. Genetic analyses using PRS corroborate these findings, suggesting the focused creative class has higher genetic liability for bipolar disorder and schizophrenia and the inactive restless has a higher genetic liability for depression and ADHD. The minimally affected class may contain a large number of individuals reporting episodes that may be considered normal variations in mood, with episodes of brief duration and low disruptiveness, with no increase in risk of disorder diagnosis or PRS. The class may comprise individuals that experience symptoms that are not captured using the pre-defined responses (in the questionnaire). As this class was the most numerous one, our findings underline the lack of specificity of the stem question in distinguishing clinically relevant periods of manic or irritable mood.

We have not assessed the degree to which cyclical definitions, when using PRS calculated from GWAS of disorders that use questionnaire-based definitions of the traits under study, influence results. The association between PRS and a latent subtype may simply be confirmatory of its inclusion in the discovery GWAS of the disorder (particularly for bipolar disorder). Similarly, the same cyclical definitions for self-reported diagnosis may be present if DSM-5 criteria select based on presence of symptoms similar to those in the LCA.

Contrasting dimensions of mood disorder symptomatology were evident between classes; with some subgroups displaying more disruptive symptoms and some less disruptive symptoms with more positive functional outcomes. The active restless class and the inactive restless class included disorganized, unproductive and unfocused characteristics, whereas the focused creative class included more creative characteristics, with higher education levels (similarly to the extensively affected class) and lower levels of episode disruptiveness. Some psychiatric disorders have been suggested to share genetics with traits such as educational attainment(31) and creativity(32,33). Given the subjective self-reported responses to the questions in the MHQ, participants own perception of their condition and impact of symptoms on their lives (or lack of awareness/insight) may mean that they perceive their episodes of manic/irritable mood less negatively (34,35)(36), but would not explain the more objective characteristic of higher educational attainment observed in the extensively affected and focused creative classes. Given the age of study participants, reported creative episodes and higher educational attainment in these two classes may precede onset and diagnosis of bipolar disorder, where the average age of onset for mood disorders is 29-43[IQR: 35-40] years of age (2). Sub-threshold episodes of elevated mood experienced earlier in life, may precede later-life bipolar disorder diagnosis and explain the observation. Further investigations into age of onset and age at which episodes were experienced may aid in resolving these questions.

Symptom groupings in the LCA suggested some redundancy between possible responses in the questionnaire. Symptoms did not all contribute equally to class separation; for example, increased confidence and creativity appeared to differentiate the focused creative and extensively affected classes from the other classes but did not separate out across classes. The five classes suggest that just four responses, one from each symptom group, would suffice to distinguish the classes from each other, with groupings of (1) increased active/talkative, (2) increased confident/creative, (3) increased restless/thoughts-racing/distracted and (4) less sleep. The observed co-occurrence of these symptoms does not preclude that individual symptoms from within groupings may further differentiate subtypes within each latent class. These results may also inform research for future updates of the diagnostic classification systems. Since symptoms do not contribute equally, a weighted approach to diagnostic criteria, rather than the current simple summation of number of symptoms present may be considered. Findings may also support a more dimensional classification of mood disorders in epidemiological research.

### Strengths and limitations

There are a number of strengths to the present study. Firstly, the use of a large, well-characterised cohort, the UK Biobank, provided a large sample size in which to conduct the present analyses and ensures that results of this study will inform future mental health research in what is an extensively studied and continuously expanding dataset. Secondly, the use of a model-based method enabled an agnostic bottom-up approach to defining latent subtypes that mitigates investigator bias of pre-defined criteria and uses the data to inform selection of the number of optimum subgroups. Finally, the replication of the identified latent classes in an independent dataset, PROTECT, demonstrates robustness and replicability of the findings in external cohorts.

There are several limitations to the present study. Firstly, the relative entropy of the optimum model in UK Biobank and PROTECT was below 0.7 indicating that subgroups may not be particularly homogenous, with some “fuzziness” between classes. To account for this, we have weighted associations with the probability of belonging to each class in multinomial regressions. Entropy is usually not considered a model selection criterion and varies depending on the data under study(29). Secondly, the study is limited by the scope of the questions asked of participants in UK Biobank regarding the manic or irritable episodes experienced. Responses were dependent on the selection of multiple-choice answers presented, and it is possible that other questions better characterise participants experiences, ultimately defining classes differently. However, since DSM-5 uses similar symptom reports, the value of any additional questions may have limited clinical relevance. Thirdly, given the use of two UK-based volunteer cohorts in restricted age-groups (generally >50 years of age), generalisability beyond this population is unknown. Finally, conclusions are limited by small sample sizes in the hospital diagnoses of disorders, and in the replication dataset where there was likely low statistical power to fully replicate associations found in the discovery.

We have used a data-driven approach, with replication in an external sample to derive latent groupings of symptoms experienced during episodes of manic or irritable mood. Our findings underline the heterogeneity of mental health disorders, but also the variation in symptoms experienced during episodes of manic/irritable mood. Findings support a dimensional classification of mood disorders and results will inform future studies of mood disorders by guiding the decisions to collect appropriate self-reported symptom data and better define subtypes of disorders with more homogenous characteristics for investigation.

## Data Availability

Data is available from the UK Biobank and PROTECT studies on request from eligible researchers.

## Acknowledgements/Funding

This paper represents independent research coordinated by the University of Exeter and King’s College London and is funded in part by the National Institute for Health Research (NIHR) Biomedical Research Centre at South London and Maudsley NHS Foundation Trust and King’s College London. This research was also supported by the NIHR Collaboration for Leadership in Applied Health Research and Care South West Peninsula, the NIHR BioResource Centre Maudsley and the NIHR Exeter Clinical Research Facility. The views expressed are those of the authors and not necessarily those of the NHS, the NIHR or the Department of Health and Social Care or King’s College London. We gratefully acknowledge the participation of all National Institute for Health Research (NIHR) BioResource, the NIHR BioResource Centre Maudsley, Biomedical Research Centre at South London and Maudsley NHS Foundation Trust and King’s College London volunteers, and thank the BioResource staff for their help with volunteer recruitment. Further information can be found at https://www.maudsleybrc.nihr.ac.uk/facilities/bioresource/. This work was funded in part by the University of Exeter through the MRC Proximity to Discovery: Industry Engagement Fund (External Collaboration, Innovation and Entrepreneurism: Translational Medicine in Exeter 2 (EXCITEME2) ref. MC_PC_17189). Genotyping of PROTECT was performed at deCODE Genetics. The authors acknowledge use of the research computing facility at King’s College London, Rosalind (https://rosalind.kcl.ac.uk), which is delivered in partnership with the NIHR Biomedical Research Centre at South London & Maudsley and Guy’s & St. Thomas’ NHS Foundation Trusts, and part-funded by capital equipment grants from the Maudsley Charity (Grant Ref. 980) and Guy’s & St. Thomas’ Charity (TR130505). Chiara Fabbri was supported by Fondazione Umberto Veronesi (https://www.fondazioneveronesi.it). Saskia Hagenaars was supported by the Medical Research Council (MR/S0151132). This research has been conducted using the UK Biobank Resource under Application Number 18177.

## Author Contributions

RA conducted the analyses and prepared the original draft. CF, EV, KASD, OP, SH, BJ, KH, JRIC were involved in data interpretation. AG was involved in methodology development and data interpretation. AC, CB, DA, BC, were involved in data acquisition/collection and data curation. CML was involved in project conceptualization and data interpretation. All authors reviewed, edited and approved the final draft.

