## Supplementary material for "Latent subtypes of manic or irritable episode symptoms in two population-based cohorts" (from another version of this preprint: version 1, 2021-04-20)

Arathimos et al.

#### Contents

**1 Supplementary Methods** **2**

**2 Supplementary Results** **4**

2.1 Results section A . . . . . . . . . . 4

**3 Supplementary Figures** **5**

**4 Supplementary Tables** **52**

### 1 Supplementary Methods

##### 1.0.1 UK Biobank

###### Phenotype data

In order to characterise probable history of bipolar disorder, questions based on the Structured Clinical Interview for DSM-IV Axis I Disorders (SCID-I) were included in the baseline questionnaire of UK Biobank. The questions included two stem questions on ever having experienced a manic/hyper/high episode, and ever having experienced an irritable episode; “Have you ever had a period of time lasting at least two days when you were feeling so good, “high”, excited or “hyper” that other people thought you were not your normal self or you were so “hyper” that you got into trouble?”; “Have you ever had a period of time lasting at least two days when you were so irritable that you found yourself shouting at people or starting fights or arguments?” The question on symptoms present during these episodes contained 4 possible responses, in addition to fields “All of the above” and “None of the above”; “Please try to remember a period when you were in a “high” or “irritable” state and select which of the following apply: I was more active than usual, I was more talkative than usual, I needed less sleep than usual, I was more creative or had more ideas than usual.” Follow-up questions asked participants to report on episode duration and episode disruptiveness; “What is the longest time period that these”high" or “irritable” periods have lasted?" with responses “At least two days but less than a week”, “Less than a week” , “A week or more”; and “How much of a problem have these”high" or “irritable” periods caused you?" with responses “No problems” and “Needed treatment or caused problems with work, relationships, finances, the law or other aspects of life”. Both the question of episode duration and episode disruptiveness included response options for “Do not know” or “Prefer not to answer”.

Sociodemographic data on participant sex, age, smoking status, alcohol intake frequency, Townsend deprivation index (TDI – as a measure of socioeconomic status) and education level were extracted from participant responses to the baseline questionnaire. Smoking status was defined as either “Never”, “Past” or “Current”. Alcohol intake frequency was defined as either “Never”, “Occasionally”, “Weekly” or “Daily”. The rank-based inverse normal transformation was applied to the TDI scores to transform a skewed distribution. Education level was defined as either “University degree”, “A-level, NVQ, HNC or HND”, “O-levels or CSE” or “None”.

In the MHQ, participants reported past diagnoses by a professional (field #20544) of several disorders. From the reports, six broad classes of disorders were defined; attention deficit hyperactivity disorder (ADHD), autism spectrum disorder (ASD), generalised anxiety disorder (GAD), depression, schizophrenia/psychosis, and mania/bipolar disorder. Cases of ‘schizophrenia/psychosis’ were defined by combining positive responses to reports of schizophrenia with reports of “any other type of psychosis or psychotic illness”. Cases of ‘generalised anxiety disorder’ (GAD) were defined by positive responses to “anxiety, nerves or generalised anxiety disorder”. Cases of ‘mania/bipolar disorder’ were defined by positive responses to “Mania, hypomania, bipolar or manic-depression”. Cases of ‘ASD’ were defined by positive responses to “Autism, Asperger’s or autistic spectrum disorder”. Cases of personality disorder were also derived from responses to the same question in the MHQ. Due to the low number of cases, self-reported personality disorder was examined separately under the category of personality traits, which also included derived neuroticism score which was based on 12 neurotic behaviour domains reported in the baseline questionnaire, previously described by Smith et al.

Linked electronic health records to Hospital Episode Statistics (HES) which contain diagnoses recorded with the International Classification of Diseases, 10th Revision (ICD-10) beginning in 1997 in England and from 1999 in Wales, up until June 2020, were used to derive cases status for 3 broad disorder definitions (termed “ICD diagnoses” or “ICD cases”). Individuals that had been assigned a primary or secondary diagnosis for depressive episode or recurrent depressive disorder (F32; F33), schizophrenia, schizotypal disorder, schizoaffective disorder and delusional/psychotic disorders (F20-F29), mood disorders including bipolar, cyclothymia, dysthymia, hypomania or mania (F30; F31; F34; F38; F39) were considered ICD cases for depression, schizophrenia/psychotic disorder, and mania/bipolar disorder, respectively. Psychiatric data from Scotland was not available (as it was currently linked), so some cases of the above disorders may have been missed.

**Genetic data**

Genotypes in UK Biobank were assayed using two different arrays (chips), the Affymetrix UK BiLEVE Axiom or Affymetrix UK Biobank Axiom array. Preliminary QC on genetic data was performed internally by UK Biobank for all participants. Using genetic principal components (PCs) provided by the UKB, 4-means clustering on the first two PCs was performed to identify individuals of European ancestry. QC was then performed using PLINK v1.9 to remove: variants with missingness > 0.02 (before individual QC), individuals with missingness > 0.02, individuals whose self-reported sex was discordant from their genetic sex, variants with missingness > 0.02 (after individual QC), variants departing from Hardy-Weinberg Equilibrium (p < 10e-8), and variants with minor allele frequency (MAF) < 0.01. Relatedness kinship estimates provided by the UK Biobank were used to identify pairs of related individuals using KING (r2 > 0.044) and the GreedyRelated algorithm was used to remove one individual from each pair, preferentially retaining individuals that survived previous QC. FlashPCA was used to generate PCs for the sub-set of individuals of European ancestry that survived QC.

##### 1.0.2 PROTECT

The PROTECT study is a UK-based online participant registry with continuous, ongoing recruitment. The PROTECT study acts as a registry as well as a study with its own aims and outcomes. The PROTECT study aims are primarily to establish risk factors for cognitive ageing and dementia. Recruitment in to the study was on the basis of local and national publicity with the majority of participants joining on the launch of the study in November 2015.

**Genetic data**

In PROTECT, genotypes were assayed in two batches using a variant of the Illumina Infinium Global Screening Array. The total numbers of participants in the combined genotyped data was 9146. Iterative filtering for call rate at 98% completeness (for individuals and SNPs) resulted in the exclusion of 84 samples. Relatedness was estimated using KING 2.2.3, followed by extraction of a list of individuals that contained no pairs of individuals with a 1st-, 2nd- or 3rd-degree relationships. Variants with Hardy-Weinberg Equilibrium p-value < 0.00001 were excluded. Individuals whose gender estimated from the genetic data did not match that reported by the study participants were excluded. Principal components (PCs) were calculated for the unrelated subset of the data using EIGENSOFT 6.1.4 after pruning. K-means clustering (assuming 4 distinct clusters) was used on the first two derived principal components to define a cluster of European ancestry individuals. Principal components were then recalculated for the cluster of individuals of European ancestry, with outlier individuals removed by EIGENSOFT if exceeding a sigma threshold of 30. Finally, individuals with excess heterozygosity (unusual patterns of genome-wide heterogeneity) calculated using the ibc function in plink v1.90 were excluded. The total number of individuals excluded was 790, resulting in a sample size of 8272.

### 2 Supplementary Results

#### 2.1 Results section A

##### 2.1.1 Associations of latent subtypes with personality traits

Associations of latent classes with self-reported diagnosis of personality disorder (N=42,183) indicated a greatly increased risk of being in all classes when compared to the minimally affected class (Figure S19-20 and Table S24). For derived neuroticism score, a one SD increase in score was associated with an increased risk of being in the extensively affected class and smaller but significant risk increases for all other classes, when compared to the minimally affected class (Figure S21 and Table S25).

##### 2.1.2 Comparisons with probable definitions of bipolar disorder

We compared the overlap of each class with cases of probable bipolar disorder type I and type II, as defined by Davis et al.from responses in the MHQ, and Smith et al. from responses to similar questions at baseline. The majority of cases of probable bipolar disorder type I defined by Davis et al. (N=931) were members of either the active restless (36.3%), or the extensively affected class (31.8%) class. For probable bipolar disorder type II (N=455), the majority of cases were members of the focused creative class (49%) (Figure S22). For the definition of probable bipolar disorder type I by Smith et al (N=248 MHQ responders), the majority of cases were members of the extensively affected class (42.7%) or the active restless class (17.7%). For probable bipolar disorder type II (N=252 MHQ responders), the majority were members of the focused creative class (21.8%) or the minimally affected class (12.7%) (Figure S23). Notably a large proportion of cases of probable bipolar disorder type II were missing class assignment, indicating that despite reporting symptoms in the baseline questionnaire that conferred them probable case status, they did not report a manic or irritable episode in the follow-up MHQ or did not answer the specific question.

### 3 Supplementary Figures

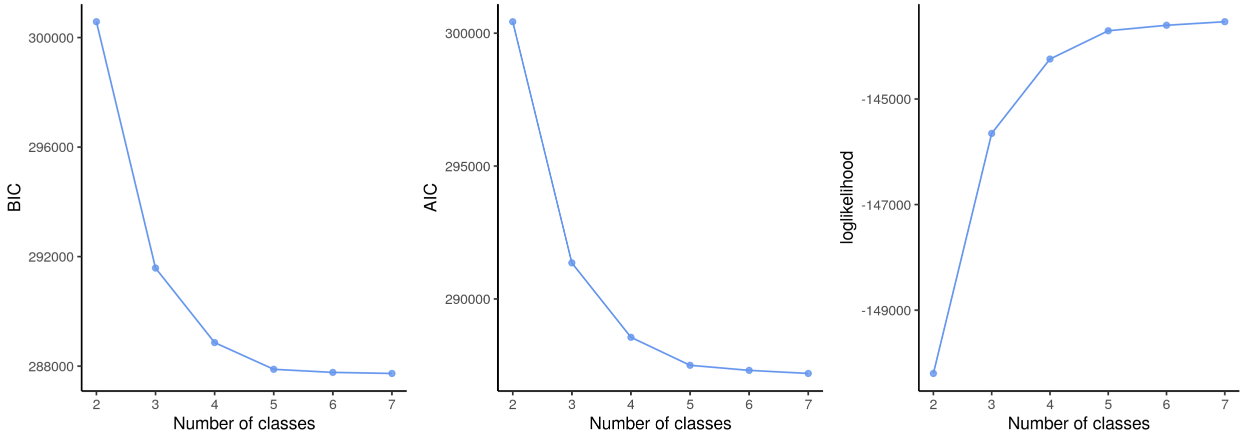


Figure S1 Elbow plots of fit statistics Bayesian Information Criterion (BIC) and Akaike Information Criterion (AIC), with log likelihood, from models with increasing number of classes (2:7 classes).

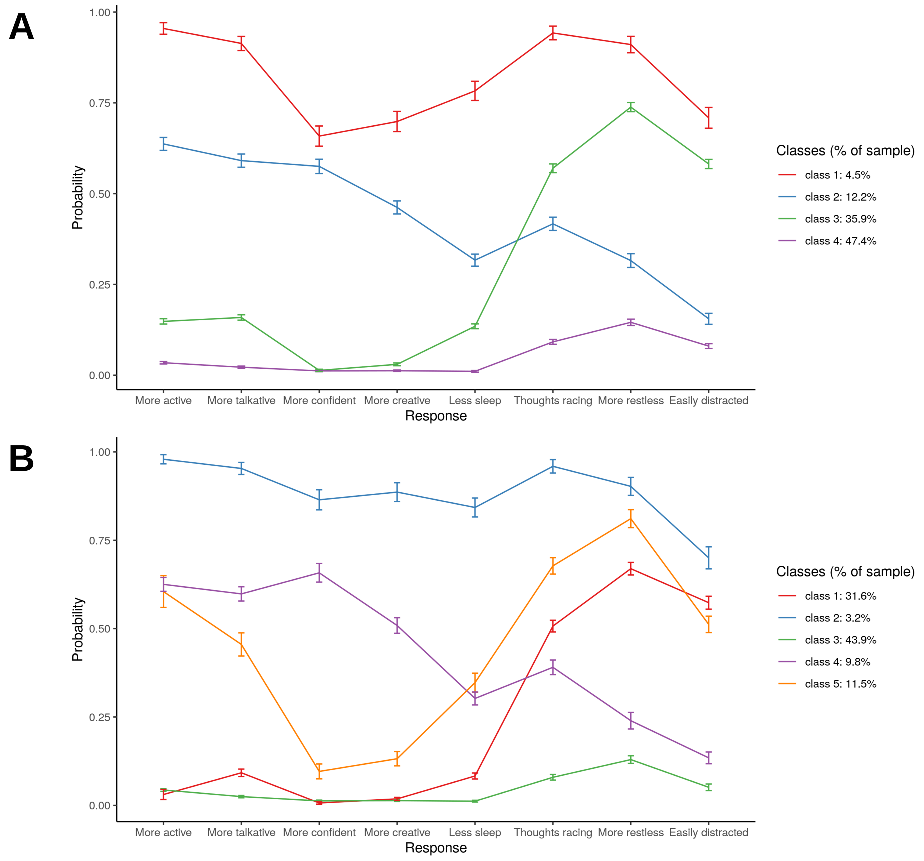


Figure S2 Comparison of conditional probabilities for each indicator symptom in the latent models with (A) 4 classes and (B) 5 classes. Note classes are unordered based on the default output from the LCA model.

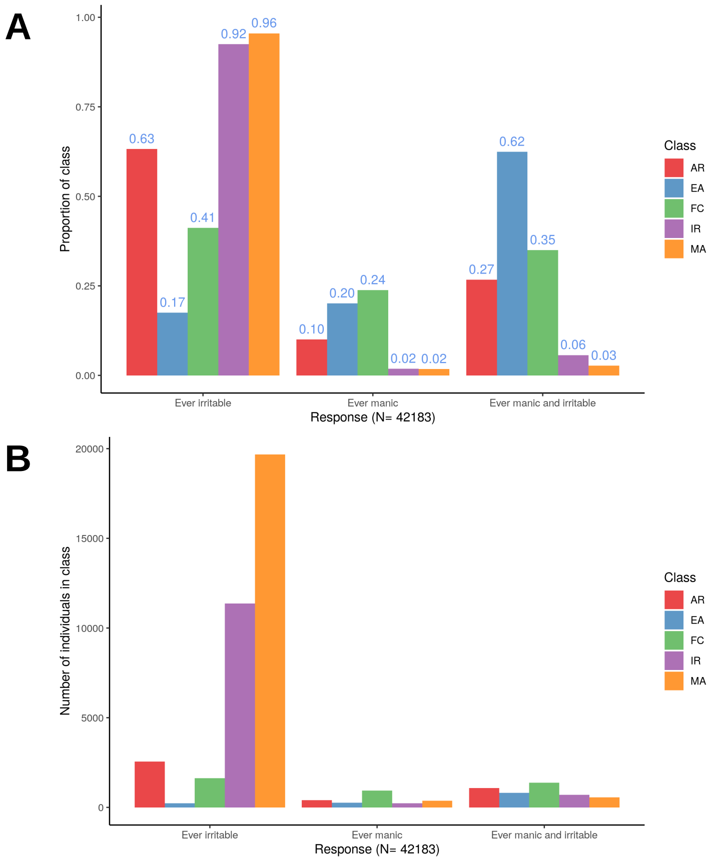


Figure S3 Distribution of responses to manic or irritable stem question by most likely class membership in the optimum 5-class model, where A) the proportion of responses in each class, and B) the absolute number of responses in each class.

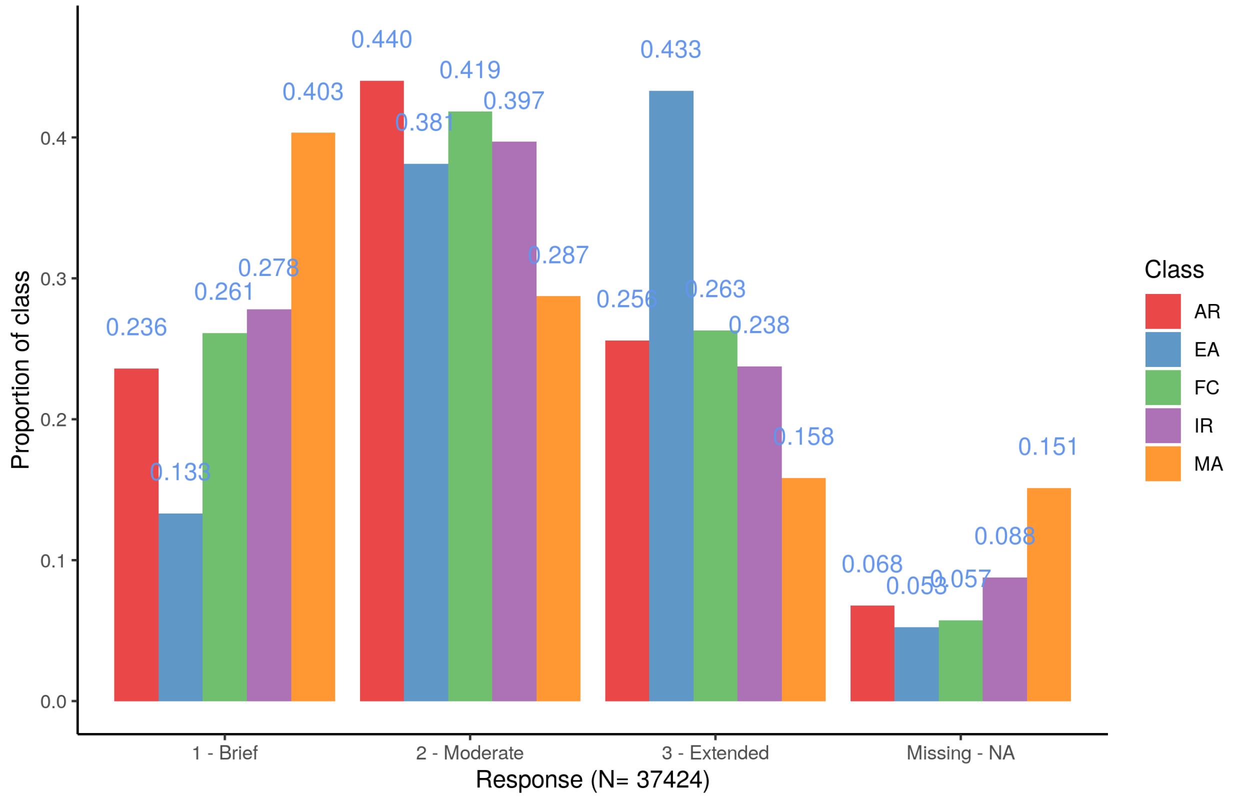


Figure S4 Distributions of responses to episode duration by most likely class membership, where "1 - Brief" <24 hours, "2 - Moderate"; duration >24 hours and < 1 week, and "3 - Extended"; duration > 1 week.

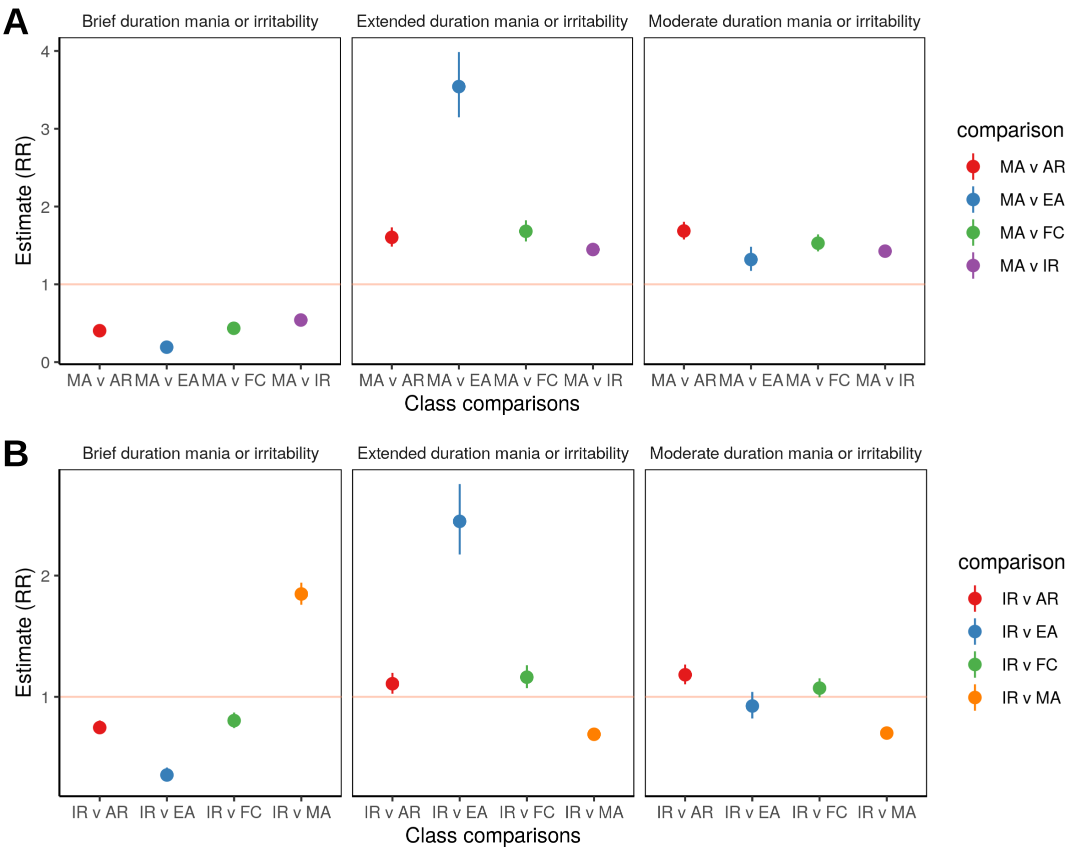


Figure S5 Associations of responses to episode duration question by most likely class membership in the optimum 5-class model.

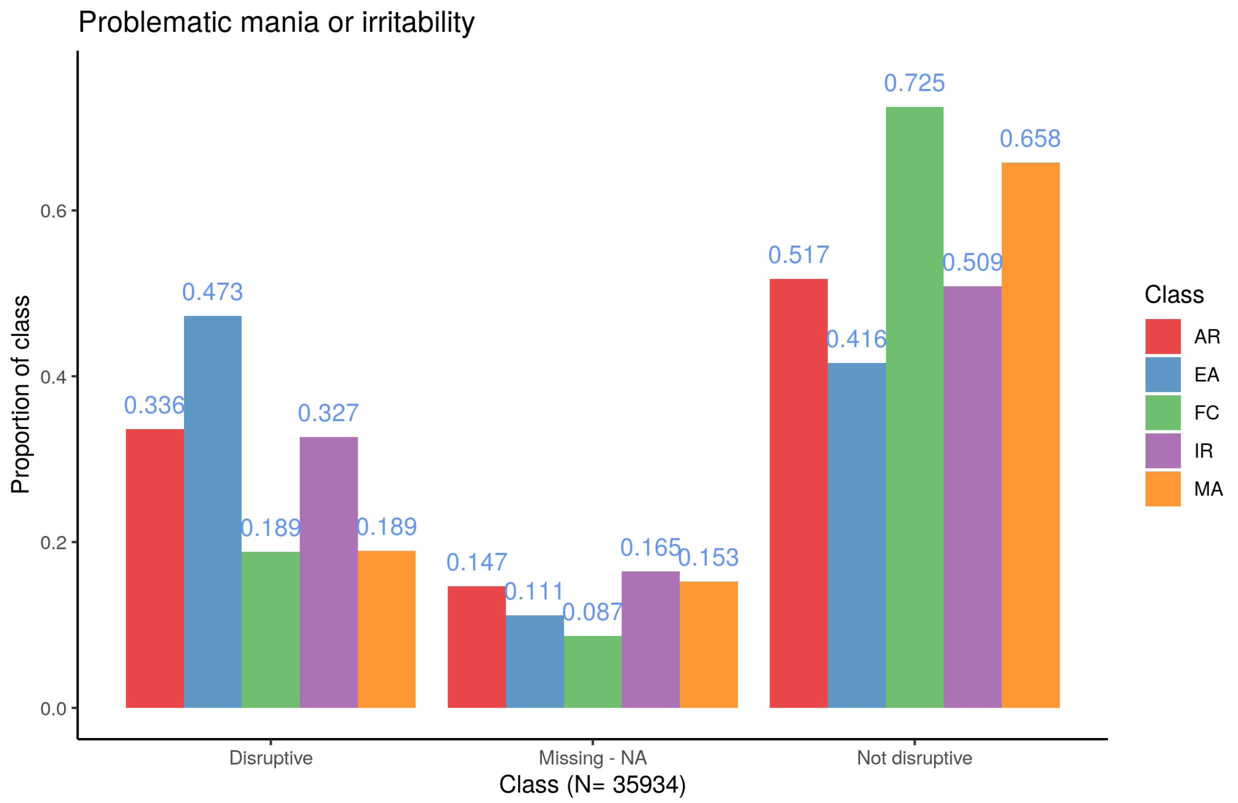


Figure S6 Distributions of responses to episode disruptiveness by most likely class membership.

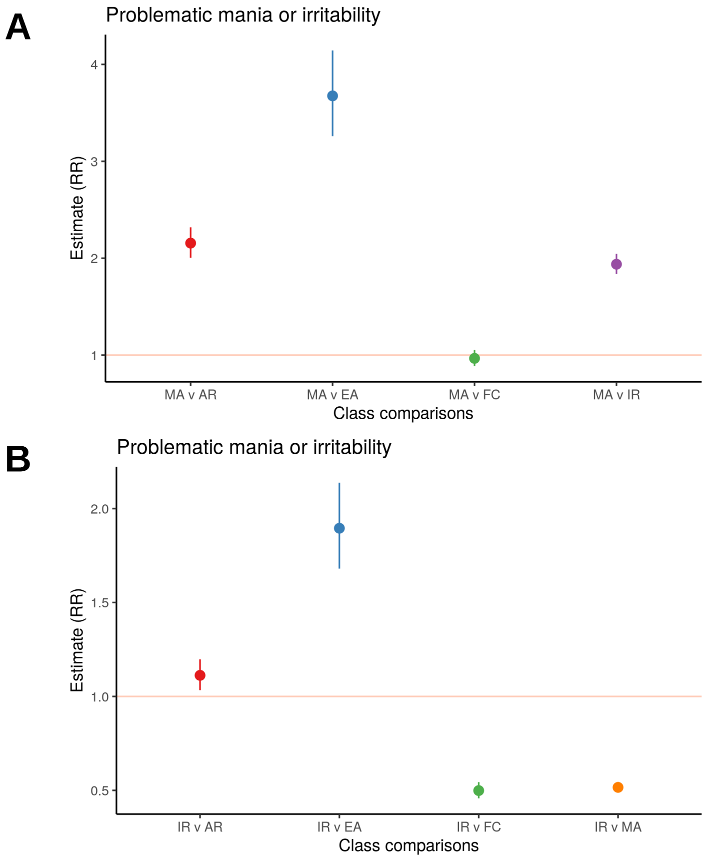


Figure S7 Associations of responses to episode disruptiveness question with most likely class membership in the optimum 5-class model.

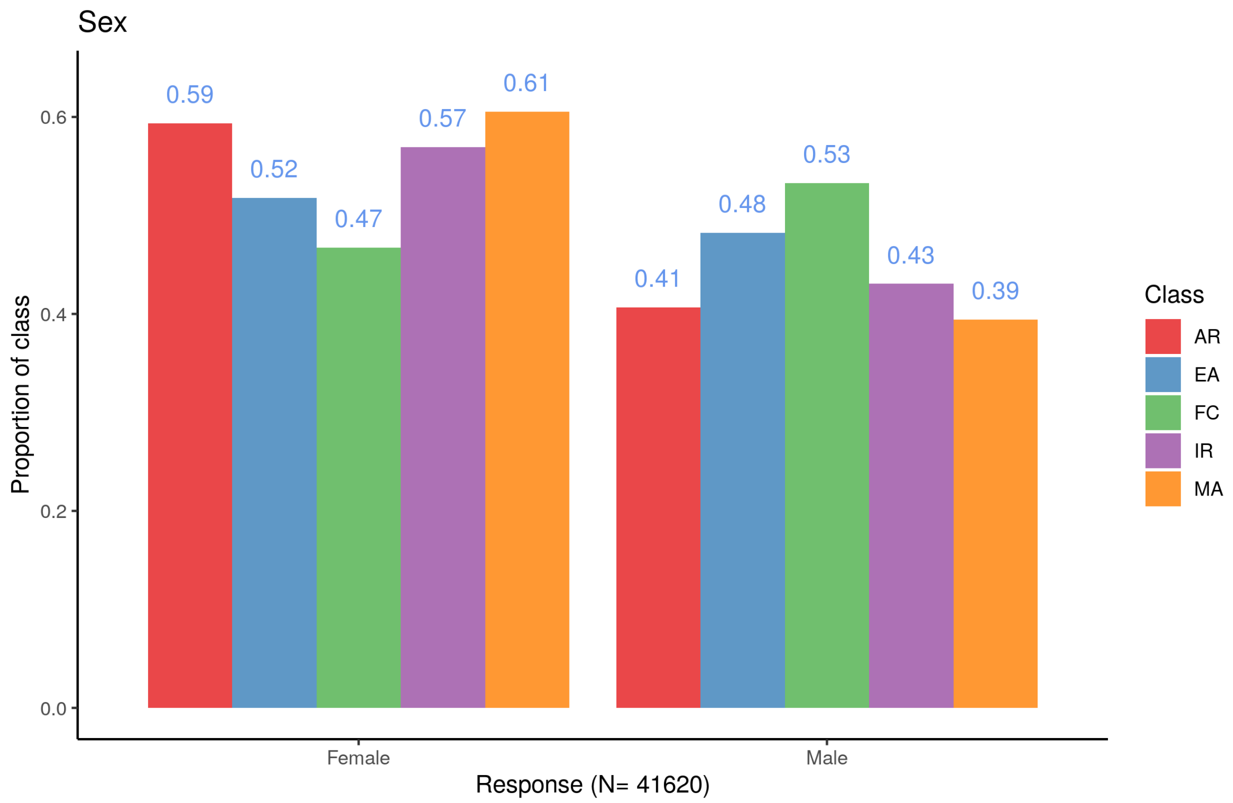


Figure S8 Distributions of sex by most likely class membership.

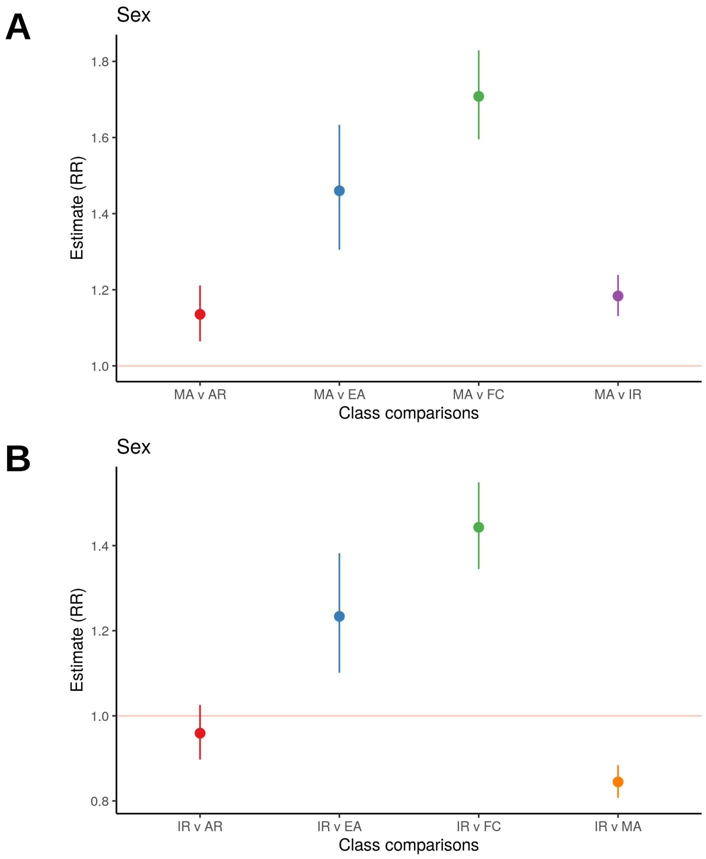


Figure S9 Associations of sex with most likely class membership in the optimum 5-class model.

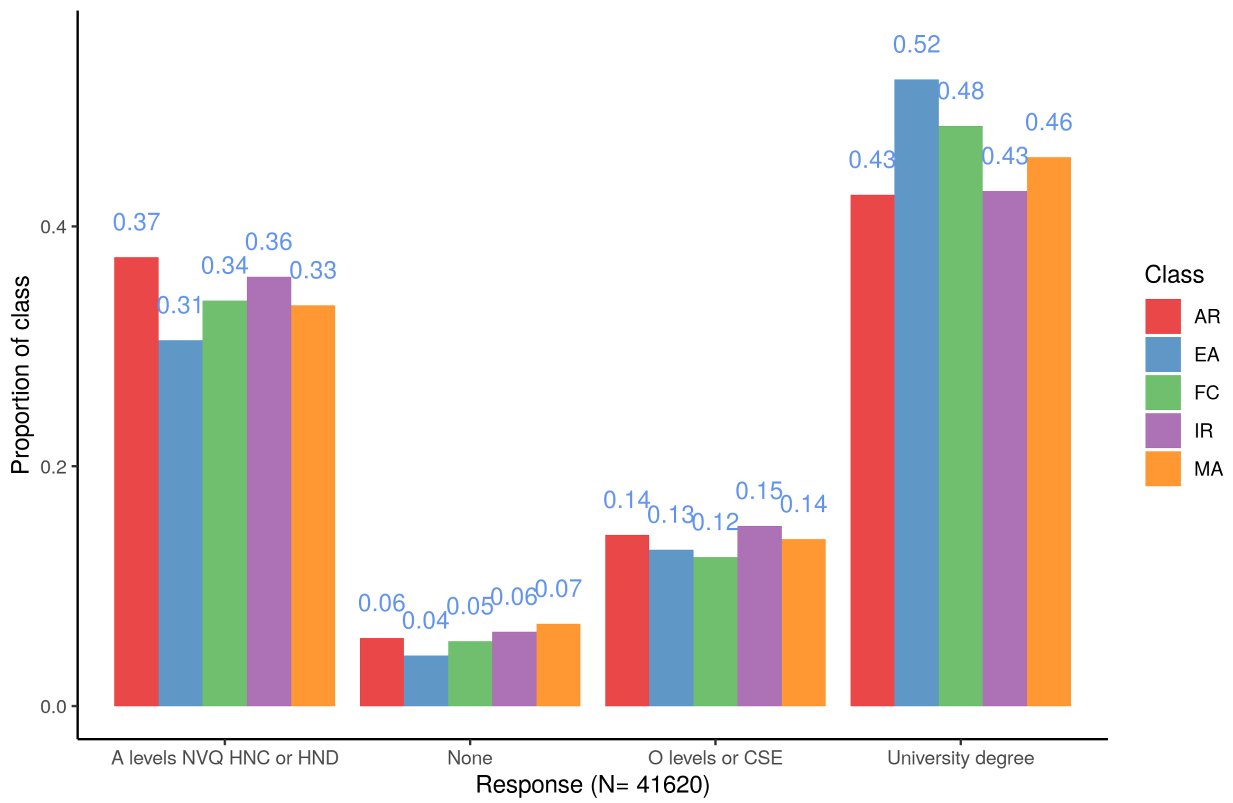


Figure S10 Distributions of educational attainment by most likely class membership.

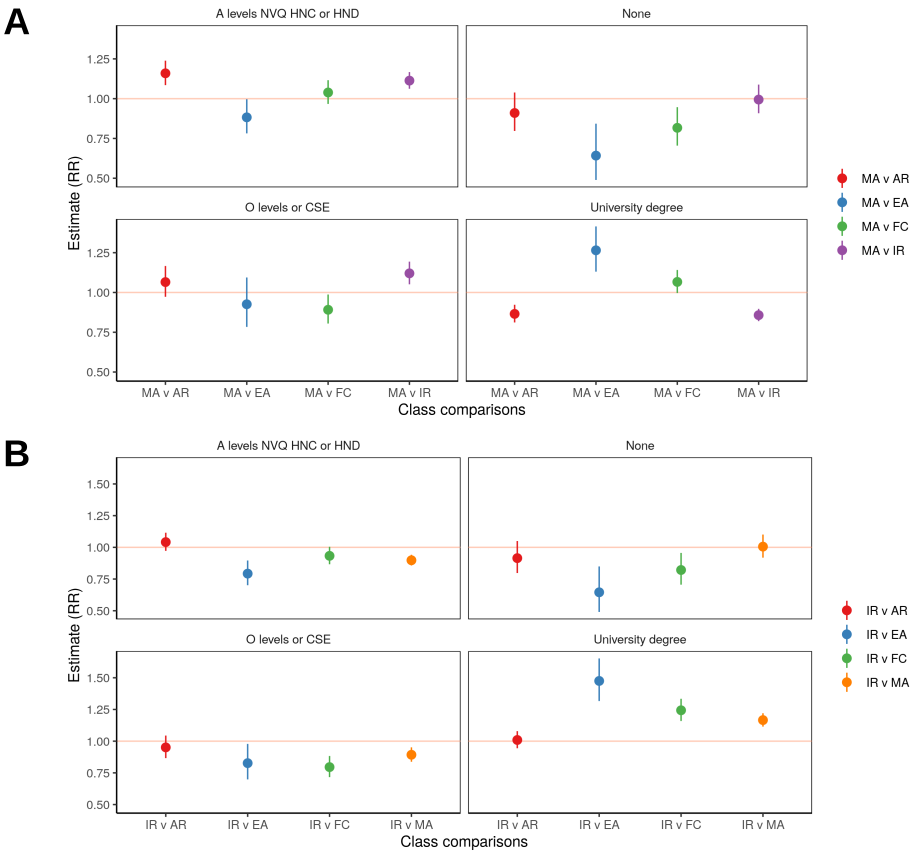


Figure S11 Associations of educational attainment with most likely class membership in the optimum 5-class model.

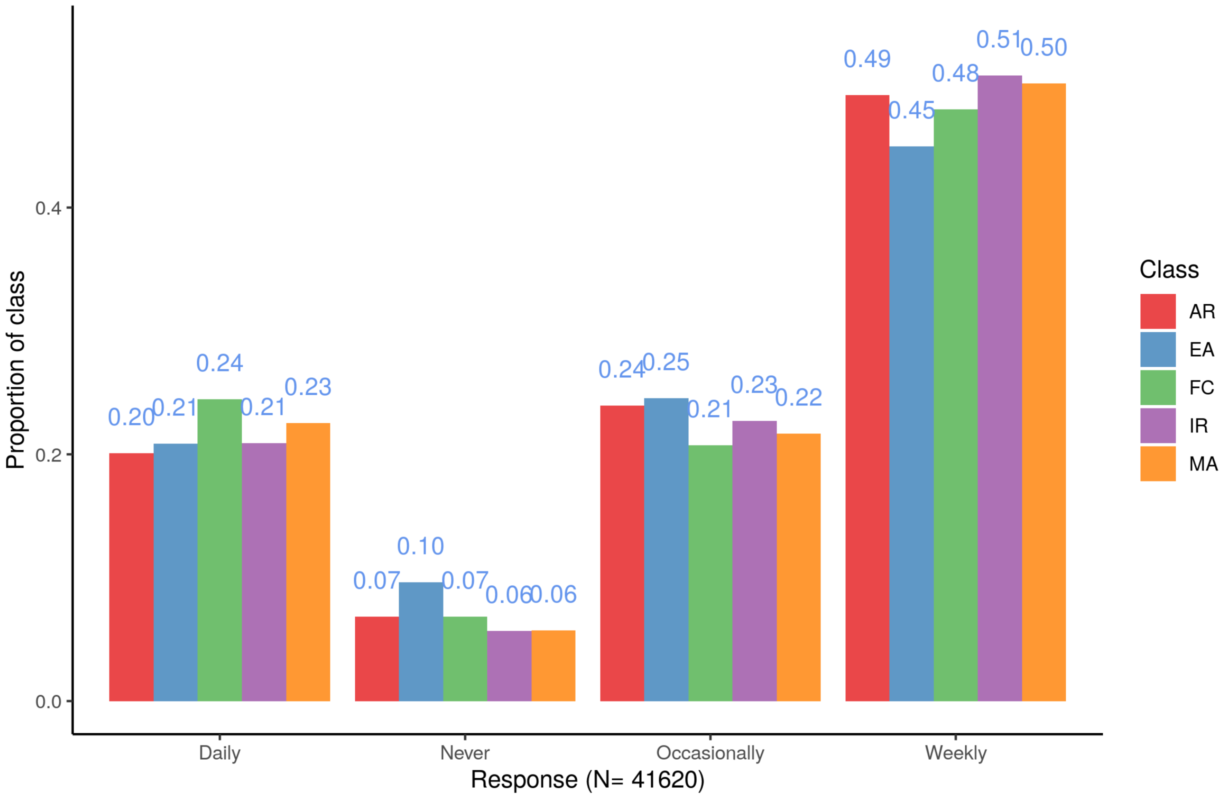


Figure S12 Distributions of alcohol consumption frequency by most likely class membership.

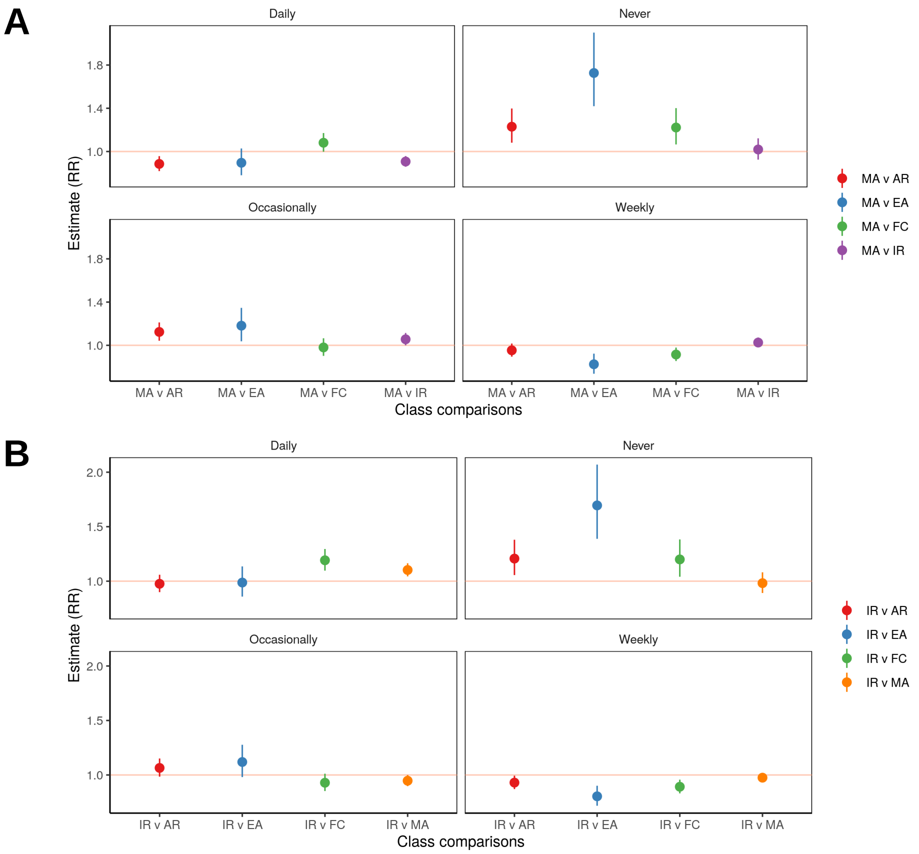


Figure S13 Associations of alcohol consumption frequency with most likely class membership in the optimum 5-class model.

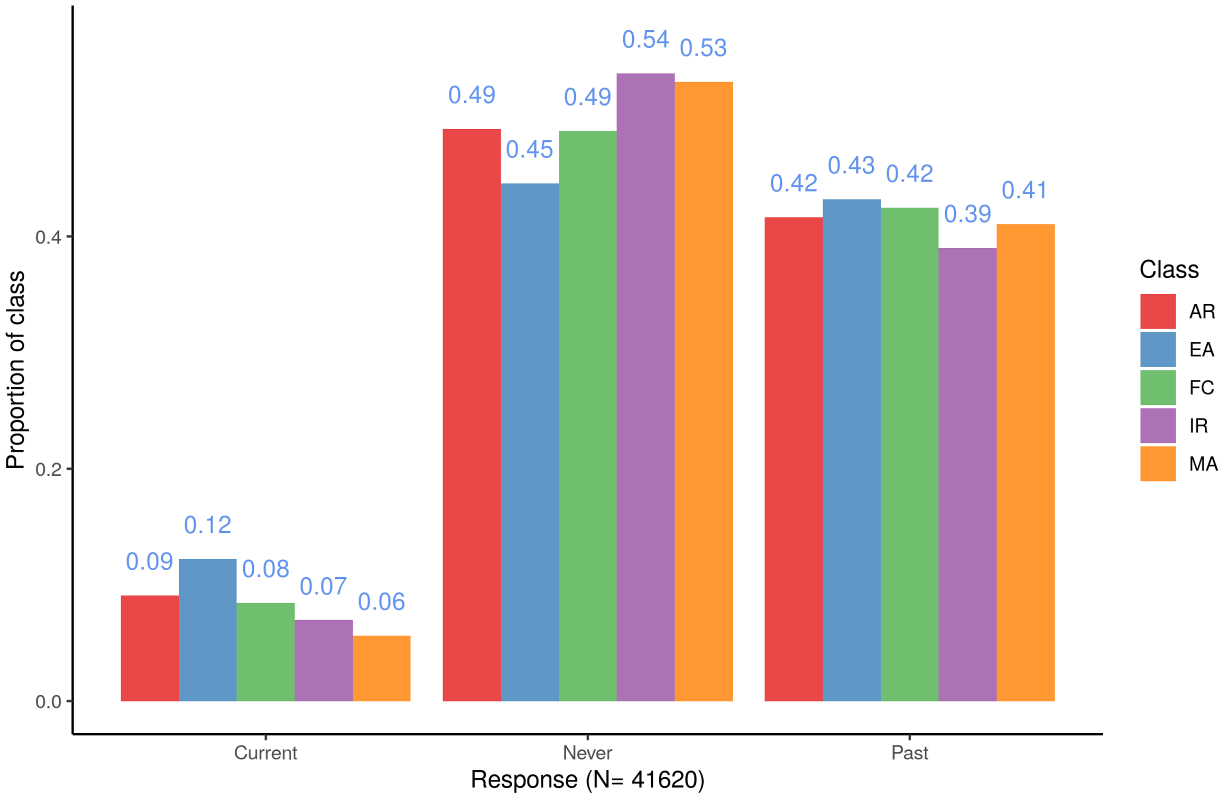


Figure S14 Distributions of smoking by most likely class membership.

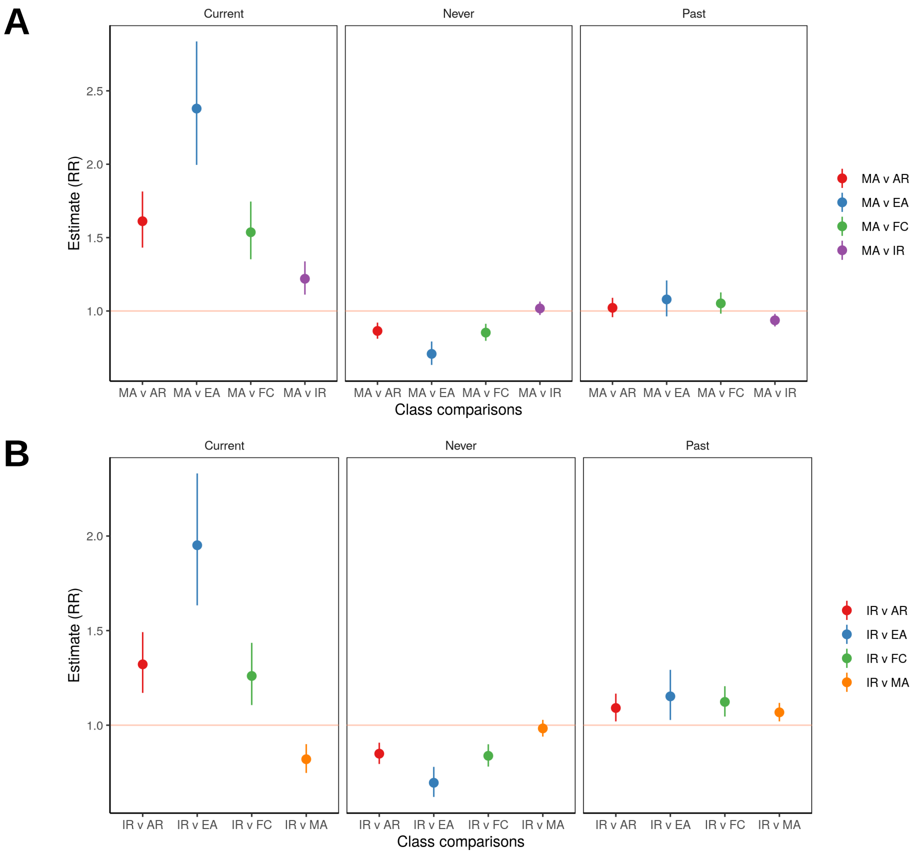


Figure S15 Associations of smoking with most likely class membership in the optimum 5-class model.

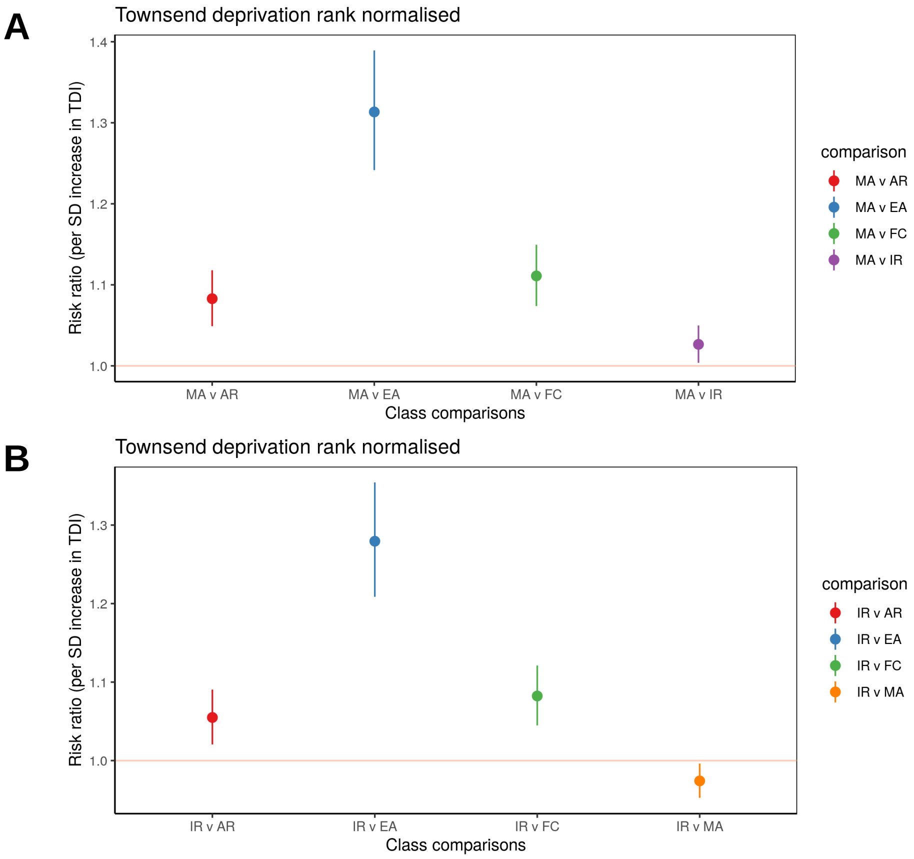


Figure S16 Associations of rank normalised Townsend deprivation index (TDI) with most likely class membership in the optimum 5-class model.

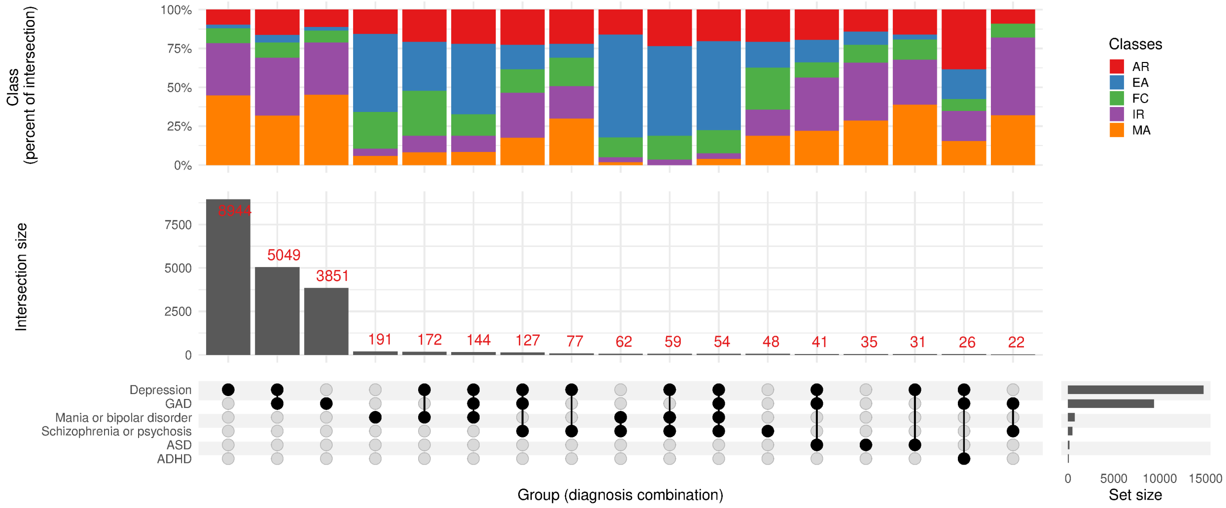


Figure S17 Distributions of different combinations of self-reported diagnoses of six psychiatric disorder (one or more diagnoses; N=19,017, 45.1% of analytical sample) by most likely class assignment. The lower panel indicates the intersection size (number of individuals reporting that combination of diagnoses). The upper panel indicates the percent of that intersection comprised by each class. Note that diagnosis combinations with fewer than 20 individuals are excluded (N=84 individuals). Classes are AR = active restless class, EA = extensively affected class, FC = focused creative class, IR = inactive restless class, MA = minimally affected class.

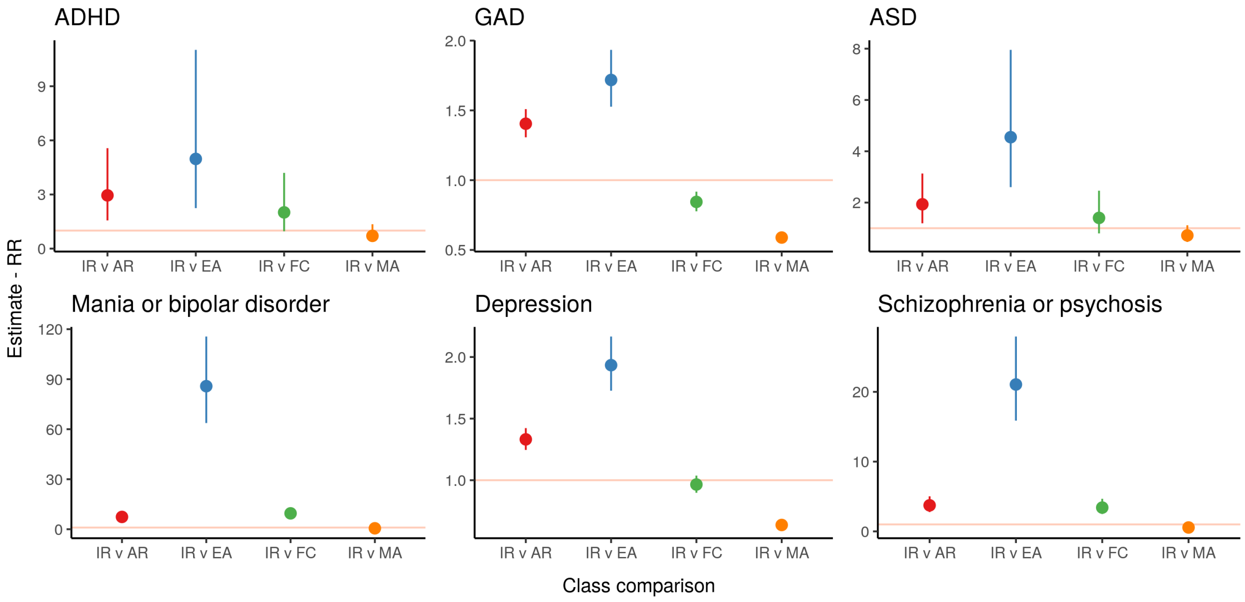


Figure S18 Associations of self-reported diagnoses of six disorders with most likely class membership in the optimum 5-class model.

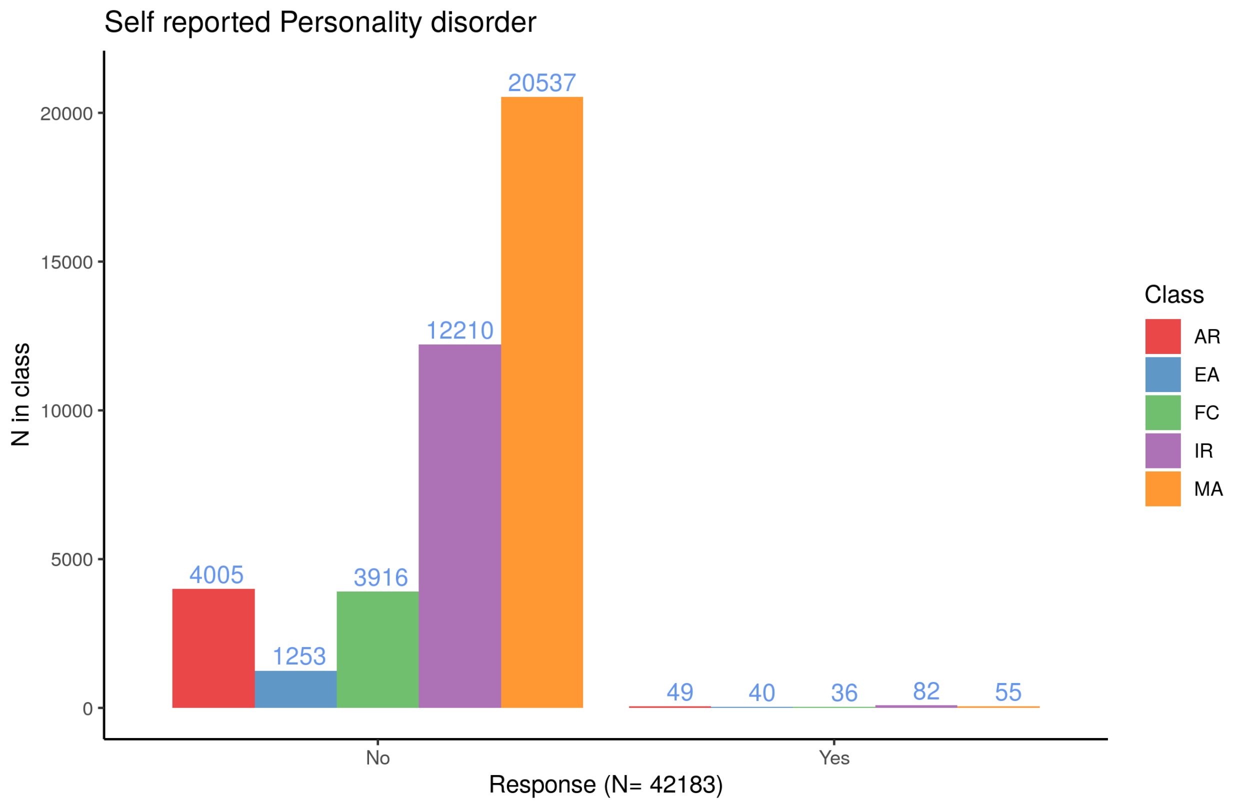


Figure S19 Distributions of self-reported diagnosis of personality disorder by most likely class membership.

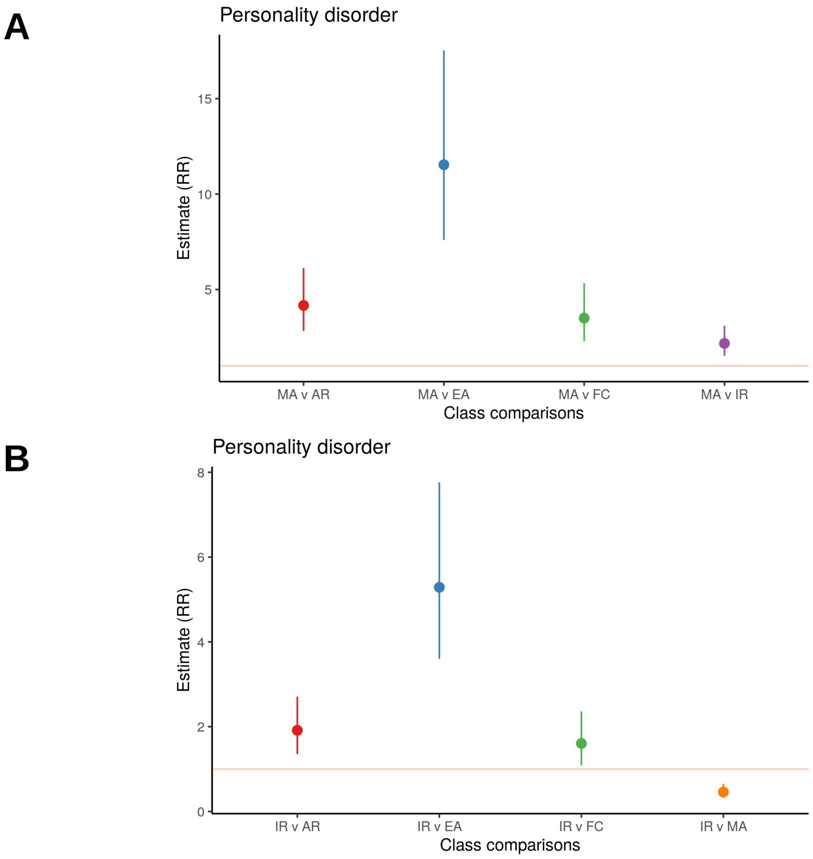


Figure S20 Associations of self-reported diagnosis of personality disorder with most likely class membership in the optimum 5-class model.

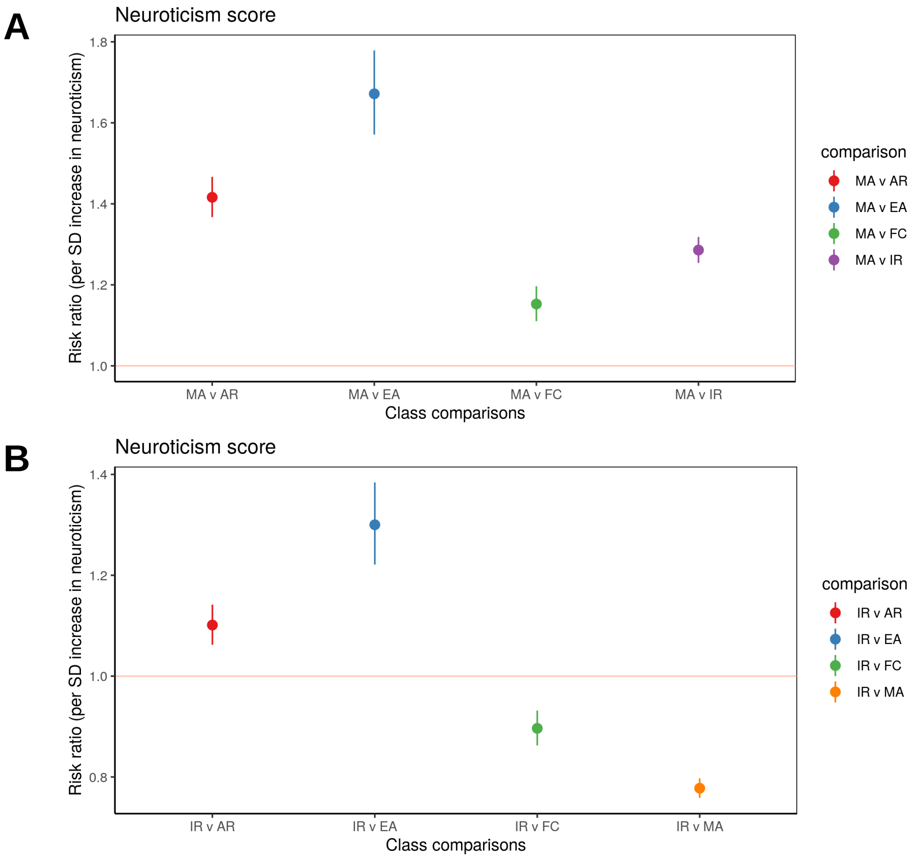


Figure S21 Associations of standardised neuroticism score with most likely class membership in the optimum 5-class model.

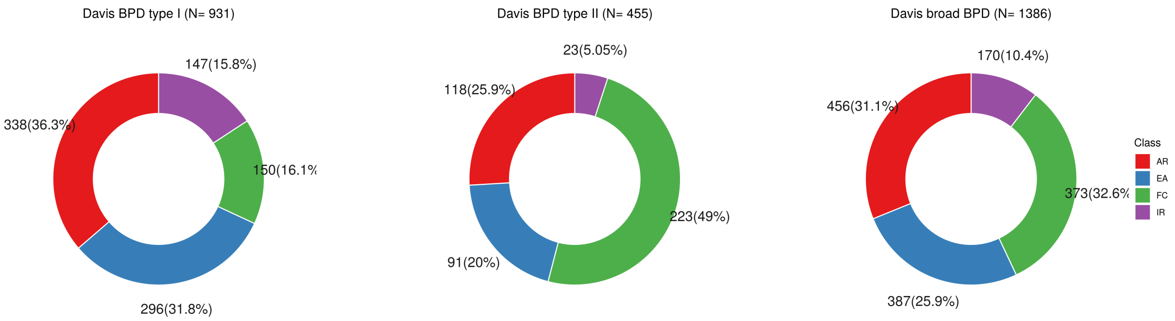


Figure S22 Comparison of the overlap of cases of probable bipolar disorder type I, bipolar disorder type II and broad bipolar disorder (combined type I and type II), as defined by Davis et al. with most likely latent class assigment.

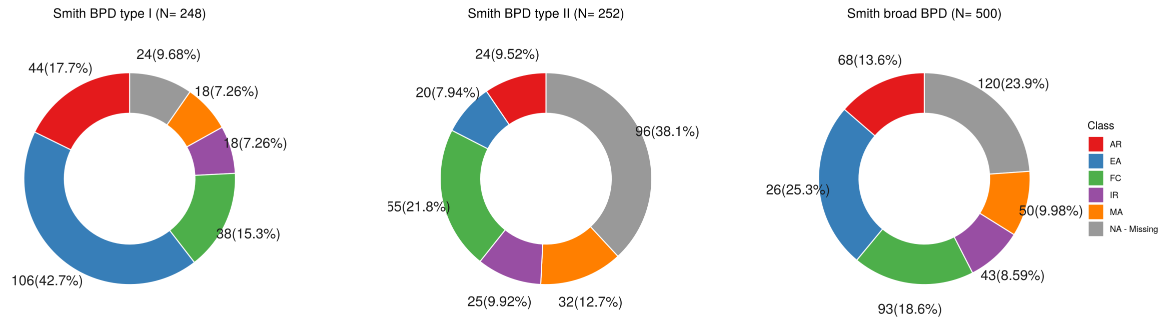


Figure S23 Comparison of the overlap of cases of probable bipolar disorder type I, bipolar disorder type II and broad bipolar disorder (combined type I and type II), as defined by Smith et al. with most likely latent class assigment.

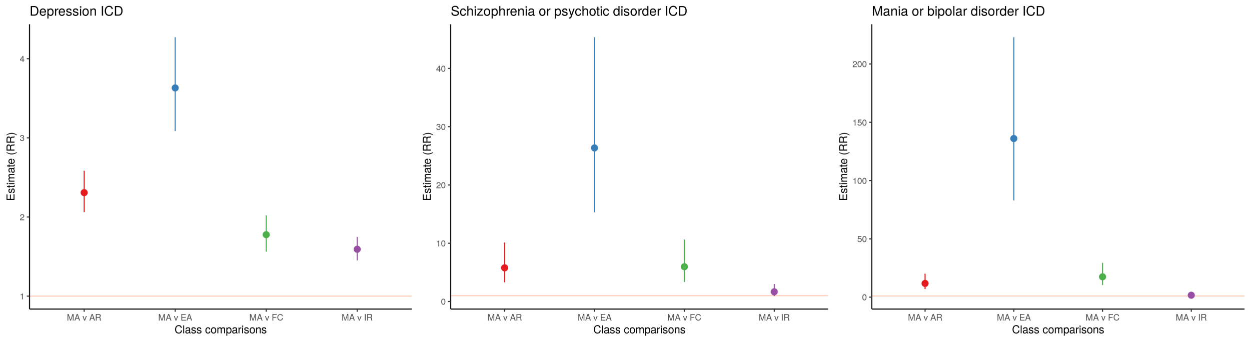


Figure S24 Associations of ICD diagnoses from hospital records of three disorders with most likely class membership in the optimum 5-class model.

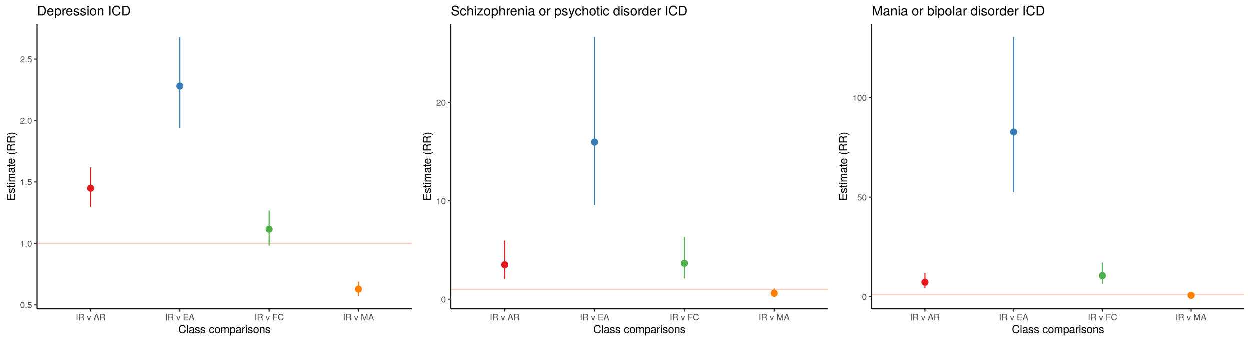


Figure S25 Associations of ICD diagnoses from hospital records of three disorders with most likely class membership in the optimum 5-class model.

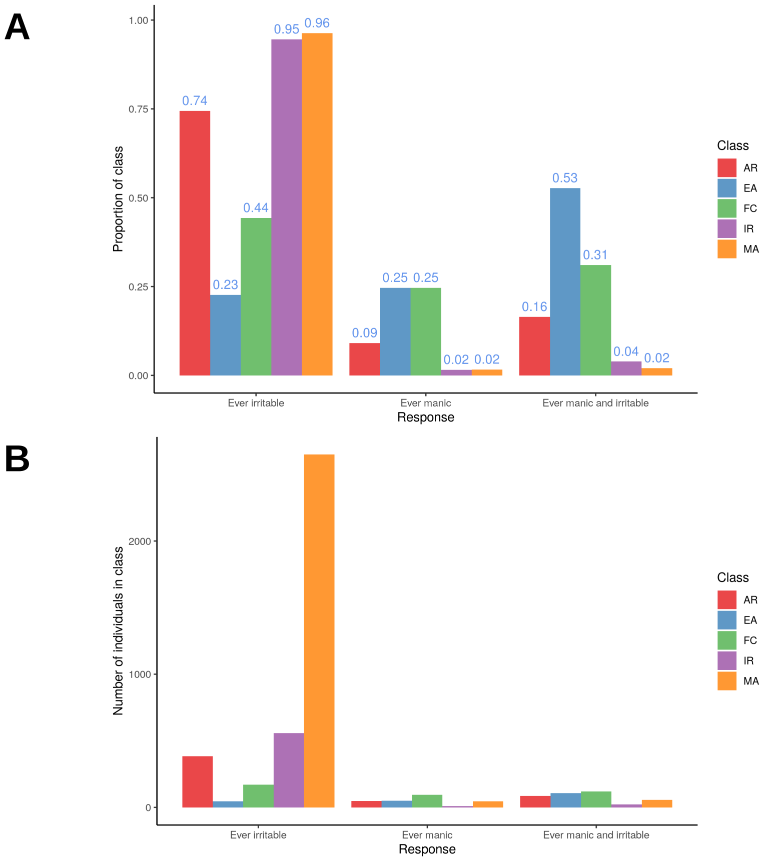


Figure S26 Distribution of responses to manic or irritable stem question by most likely class membership in the optimum 5-class model in PROTECT, where A) the proportion of responses in each class, and B) the absolute number of responses in each class.

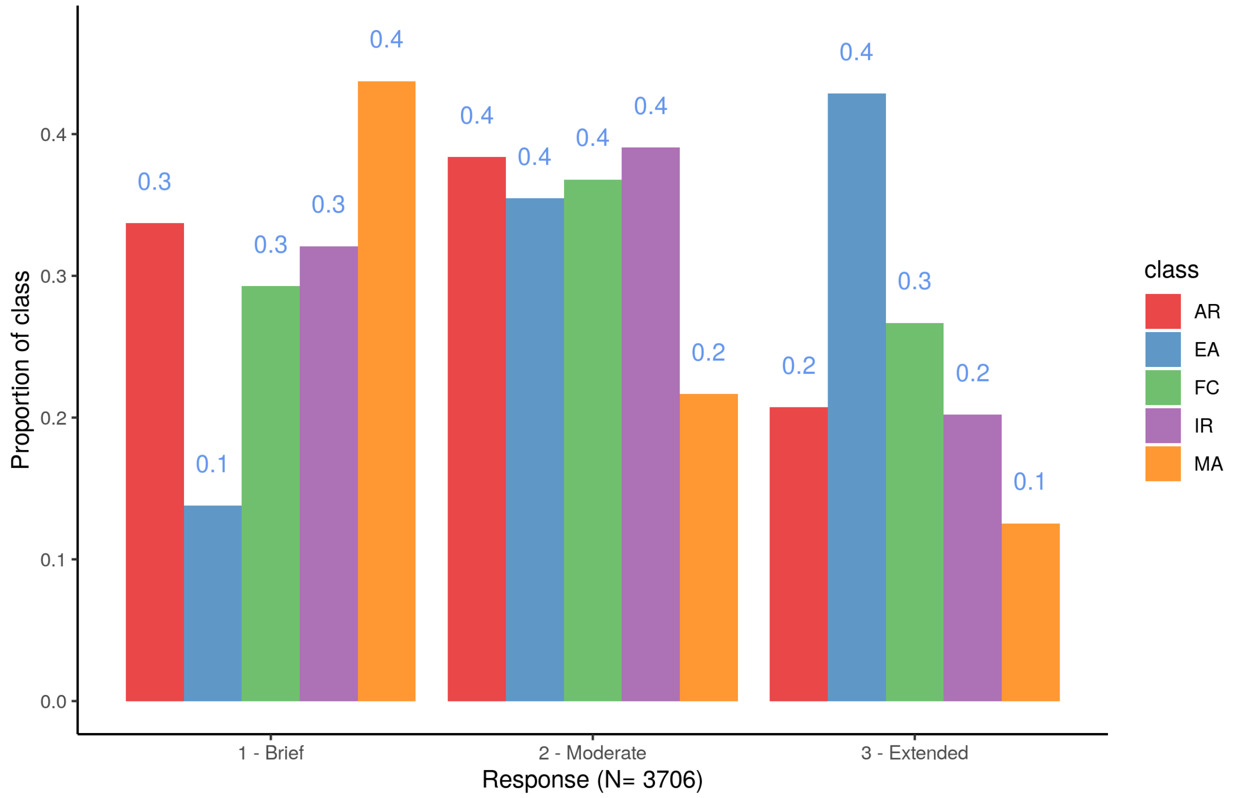


Figure S27 Distributions of responses to episode duration by most likely class membership, where “1 - Brief” <24 hours, “2 - Moderate”; duration >24 hours and < 1 week, and “3 - Extended”; duration > 1 week in PROTECT.

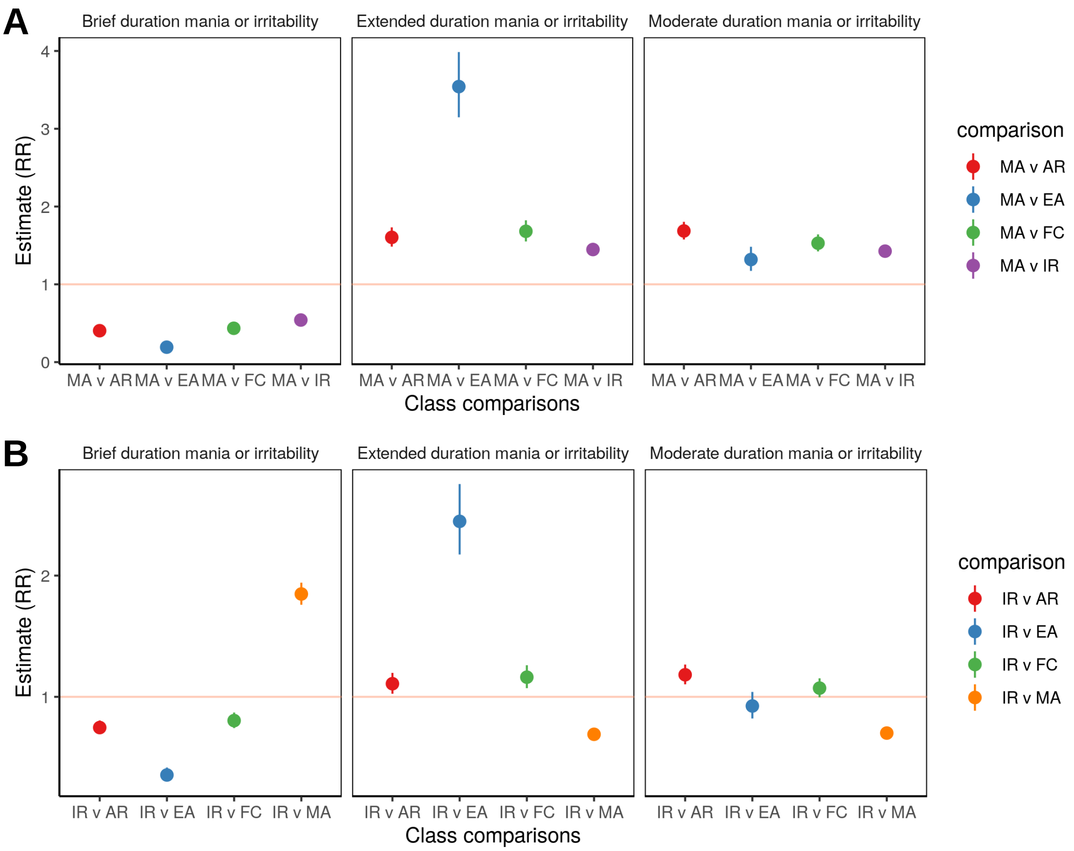


Figure S28 Associations of responses to episode duration question by most likely class membership in the optimum 5-class model in PROTECT.

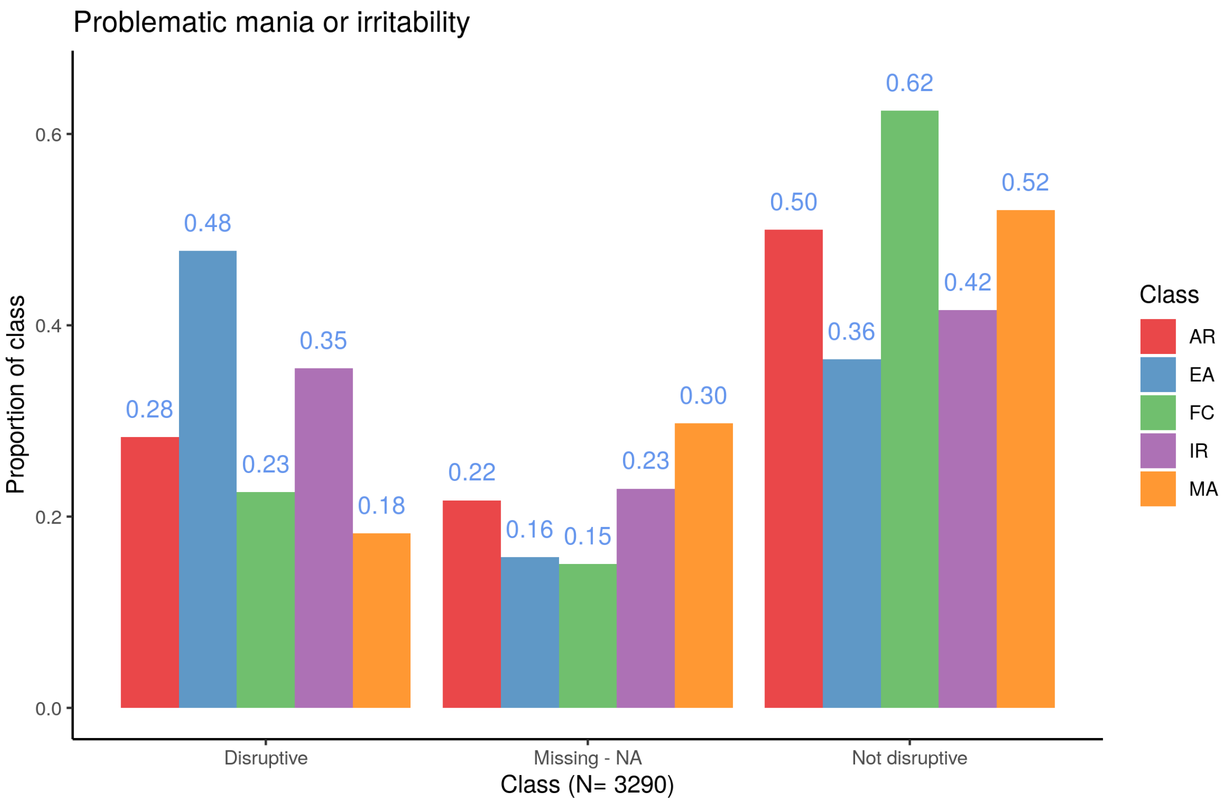


Figure S29 Distributions of responses to episode disruptiveness by most likely class membership in PROTECT.

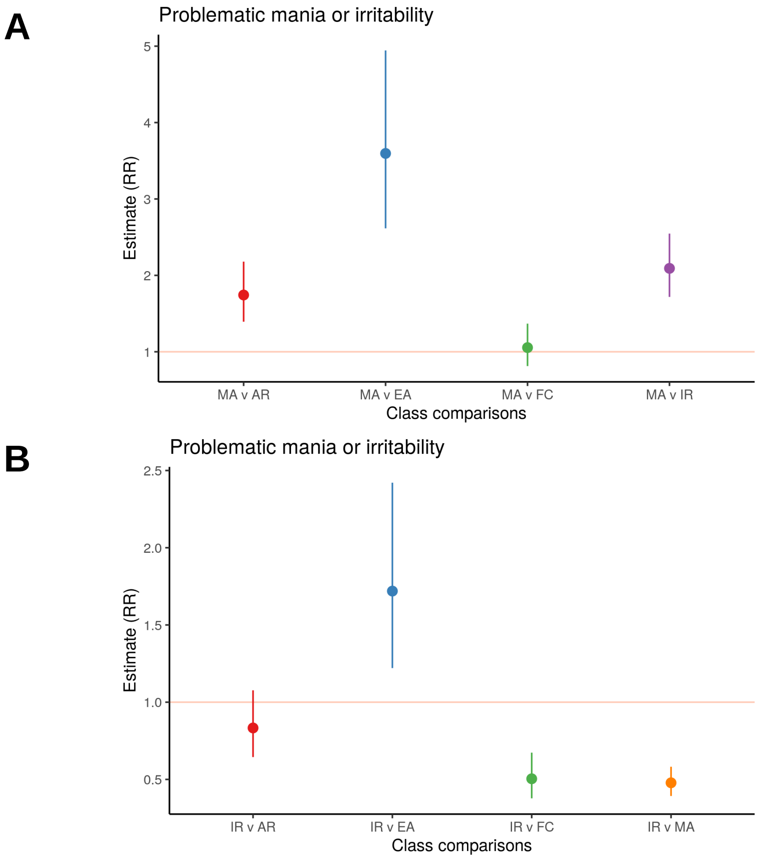


Figure S30 Associations of responses to episode disruptiveness question with most likely class membership in the optimum 5-class model in PROTECT.

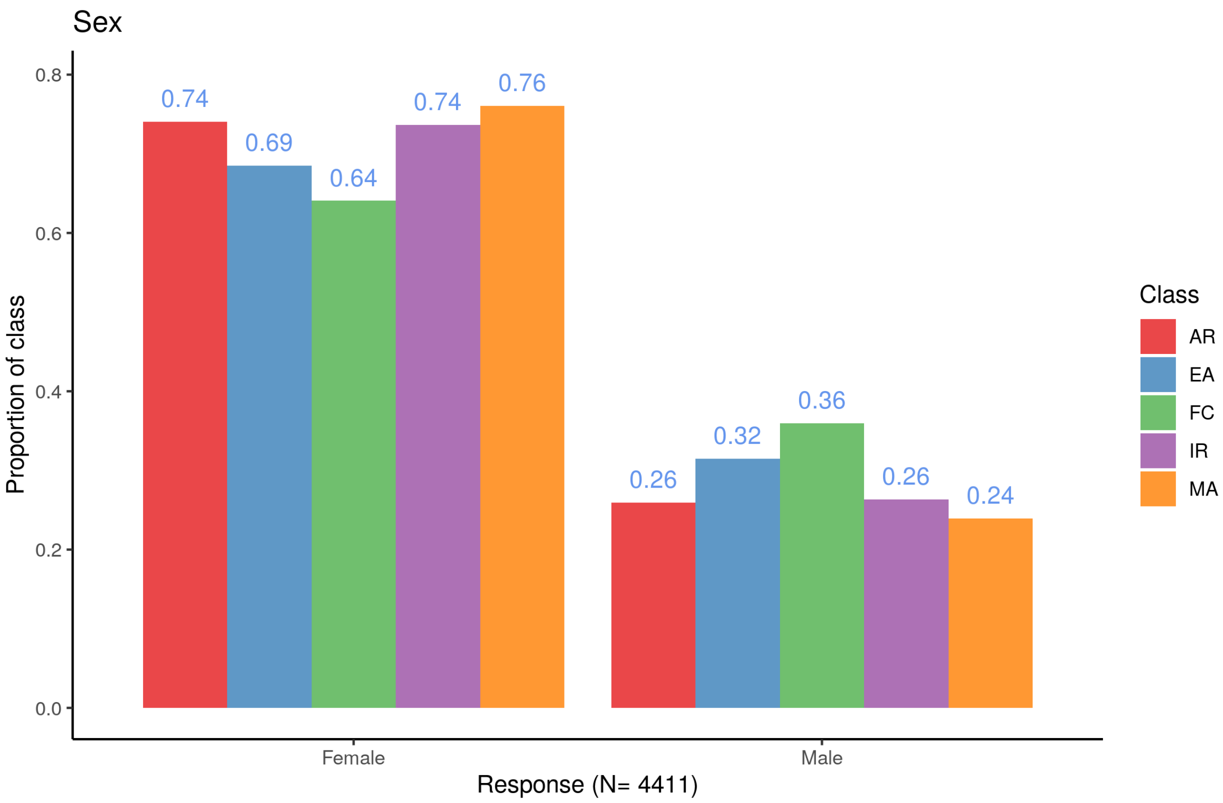

Figure S31 Distributions of sex by most likely class membership in PROTECT.

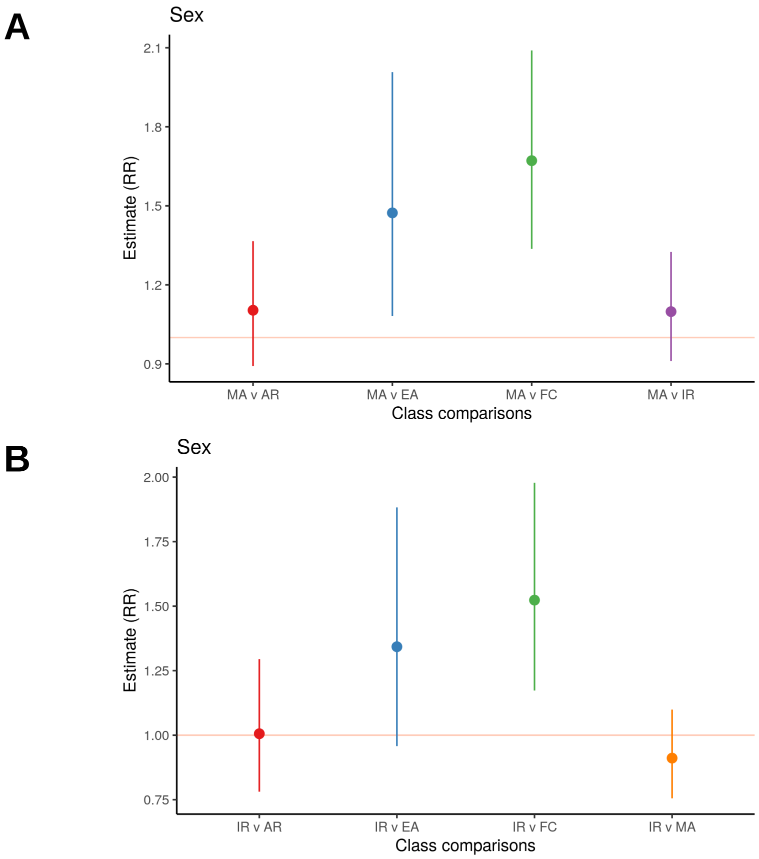


Figure S32 Associations of sex with most likely class membership in the optimum 5-class model in PROTECT.

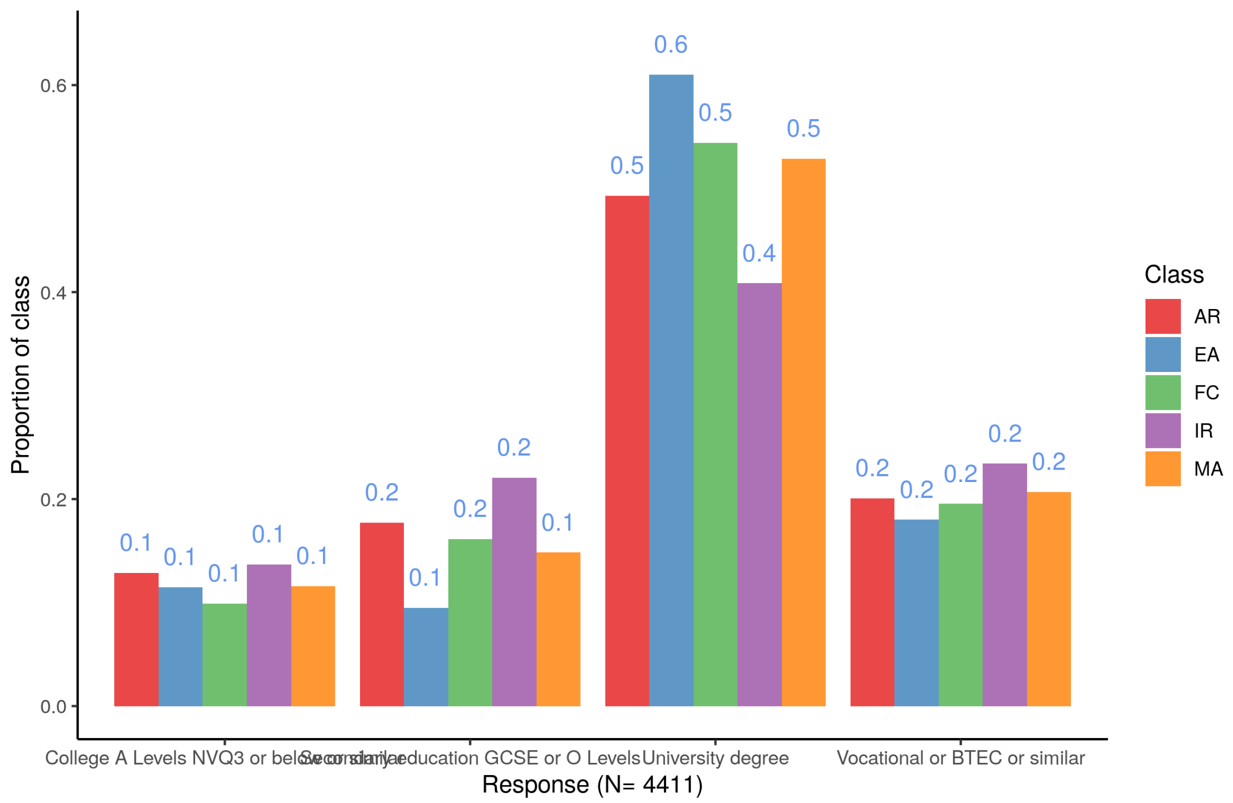


Figure S33 Distributions of educational attainment by most likely class membership in PROTECT.

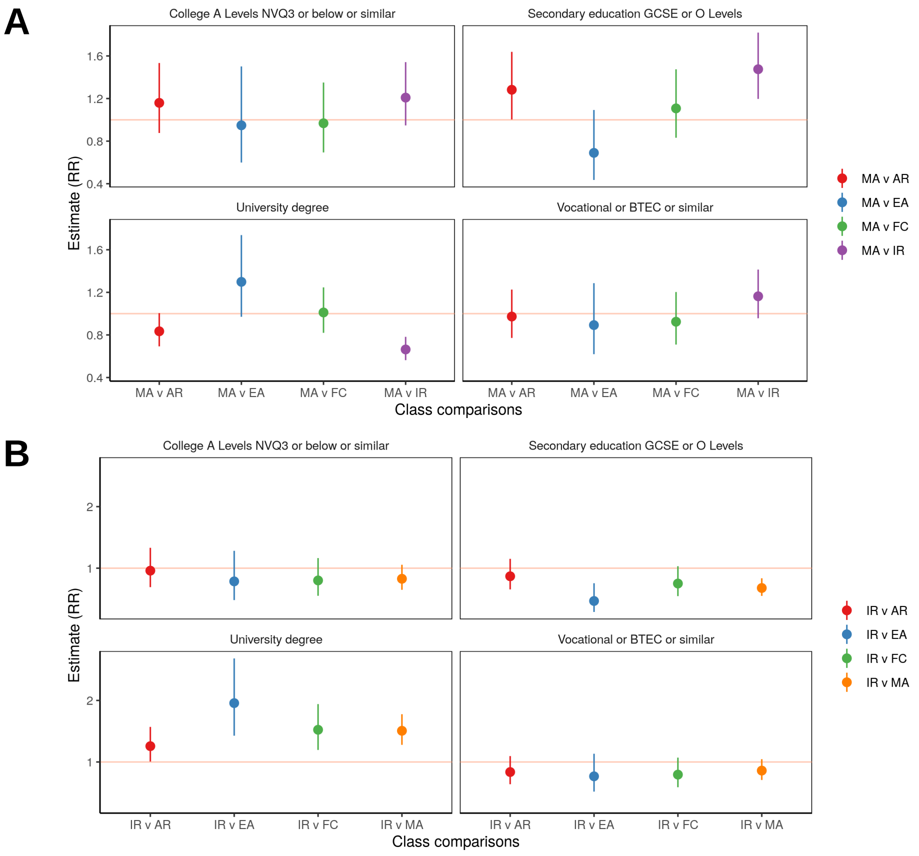


Figure S34 Associations of educational attainment with most likely class membership in the optimum 5-class model in PROTECT.

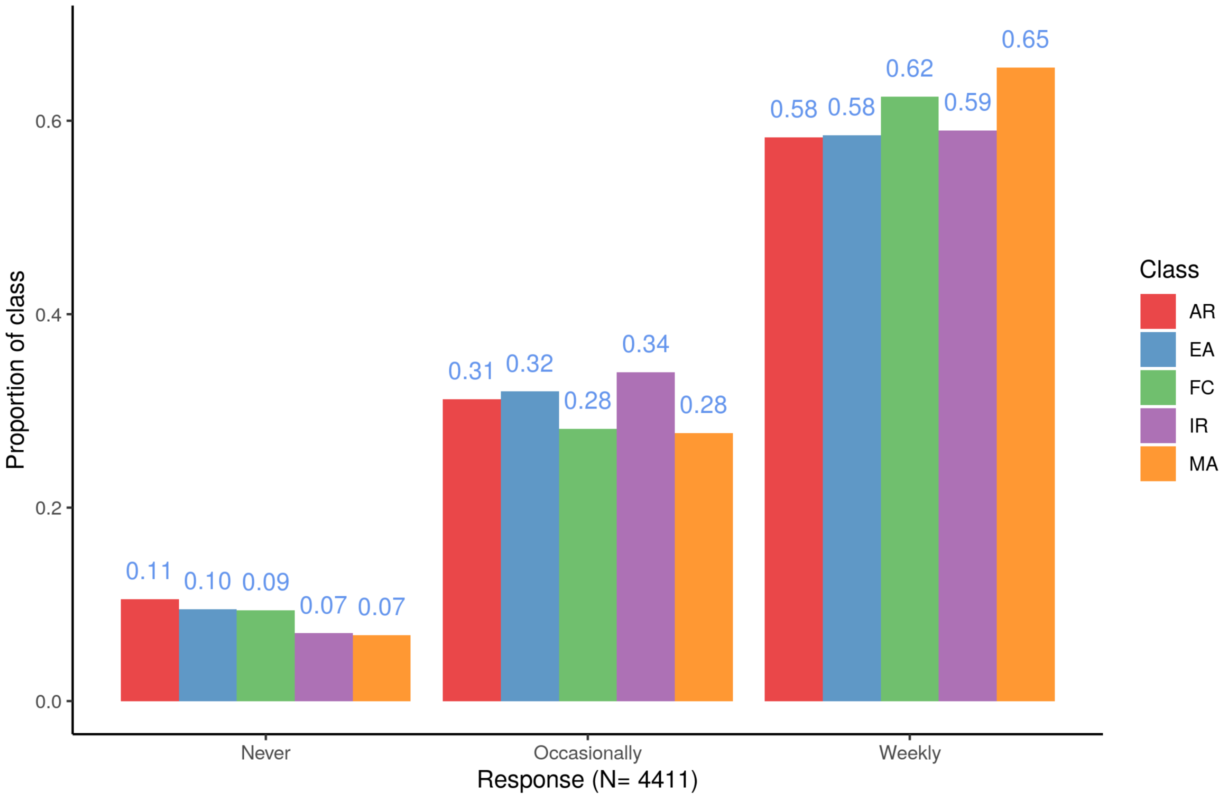


Figure S35 Distributions of alcohol consumption frequency by most likely class membership in PROTECT.

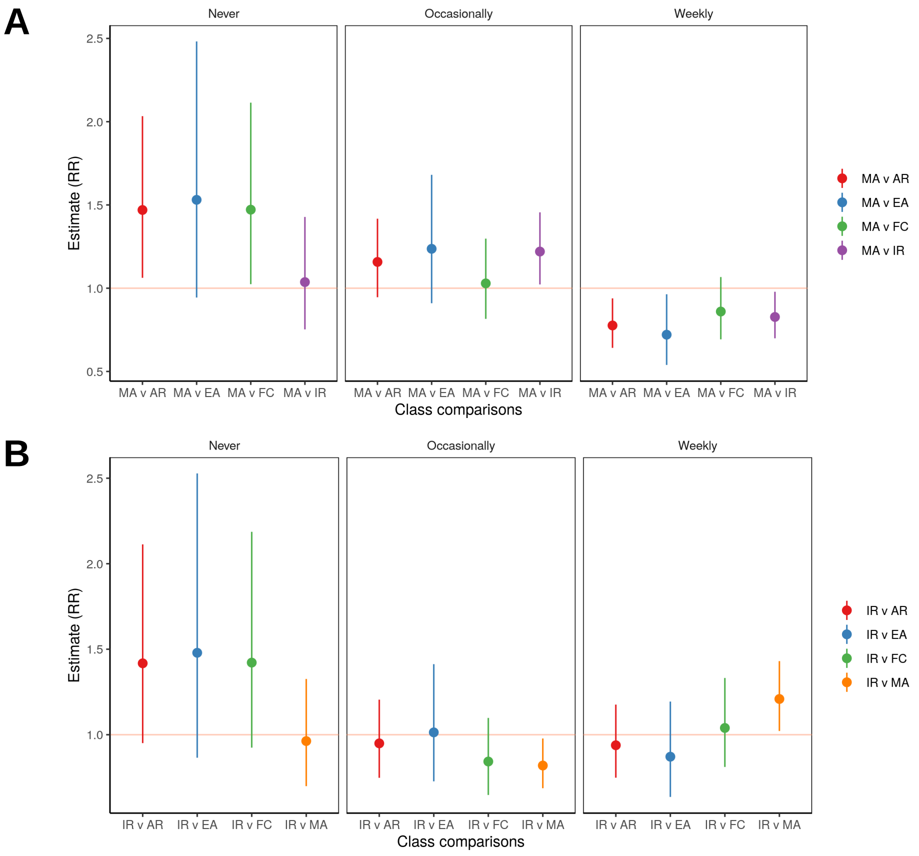


Figure S36 Associations of alcohol consumption frequency with most likely class membership in the optimum 5-class model in PROTECT.

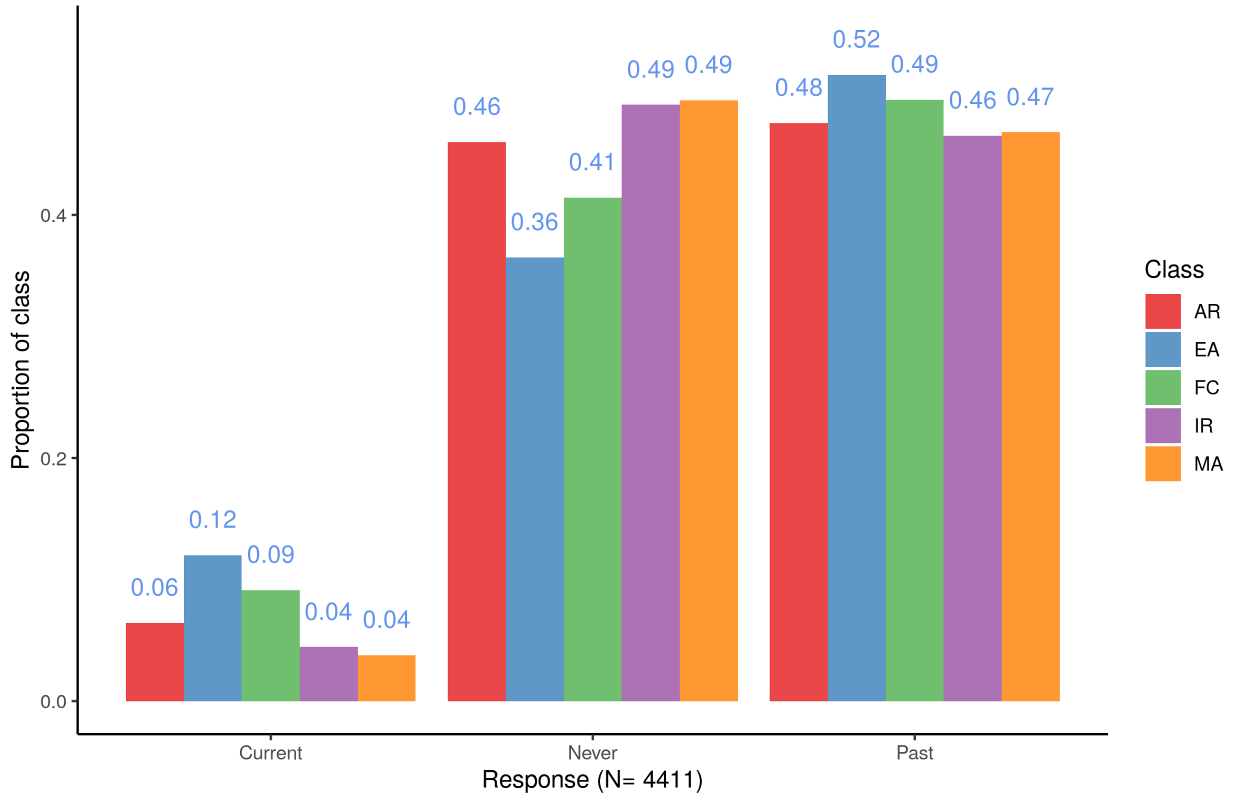


Figure S37 Distributions of smoking by most likely class membership.

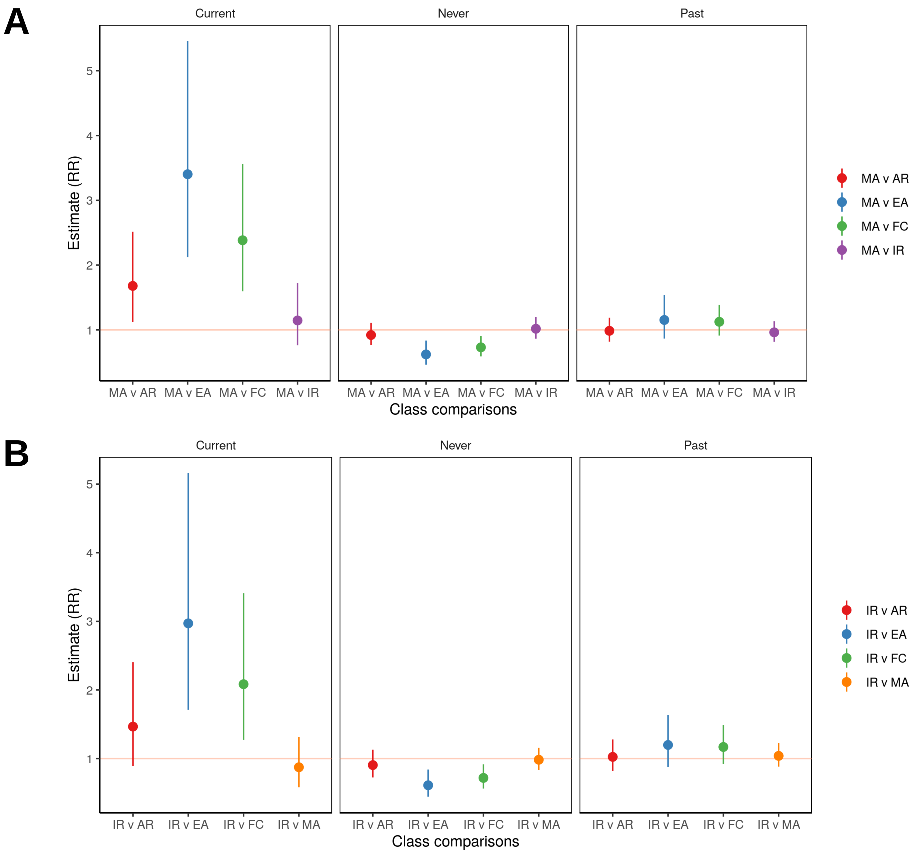


Figure S38 Associations of smoking with most likely class membership in the optimum 5-class model in PROTECT.

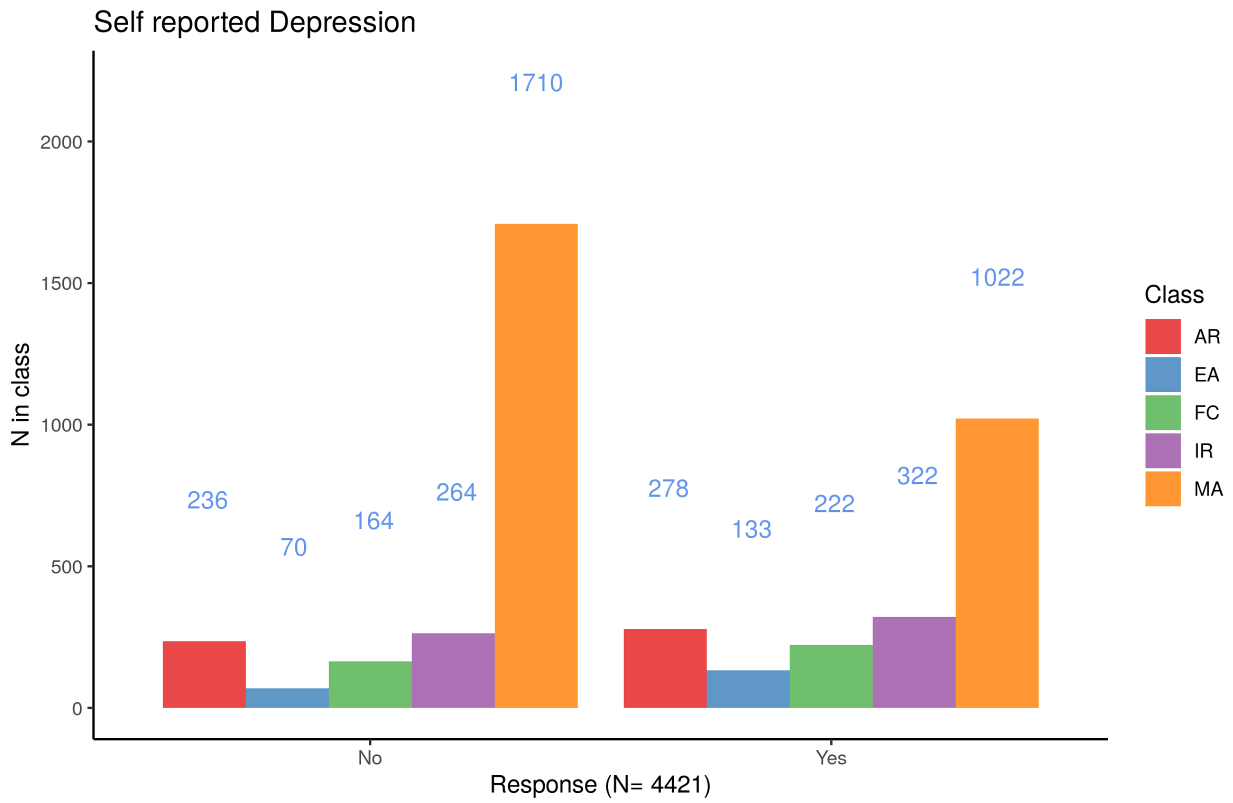

Figure S39 Distributions of self-reported diagnosis of depression by most likely class membership.

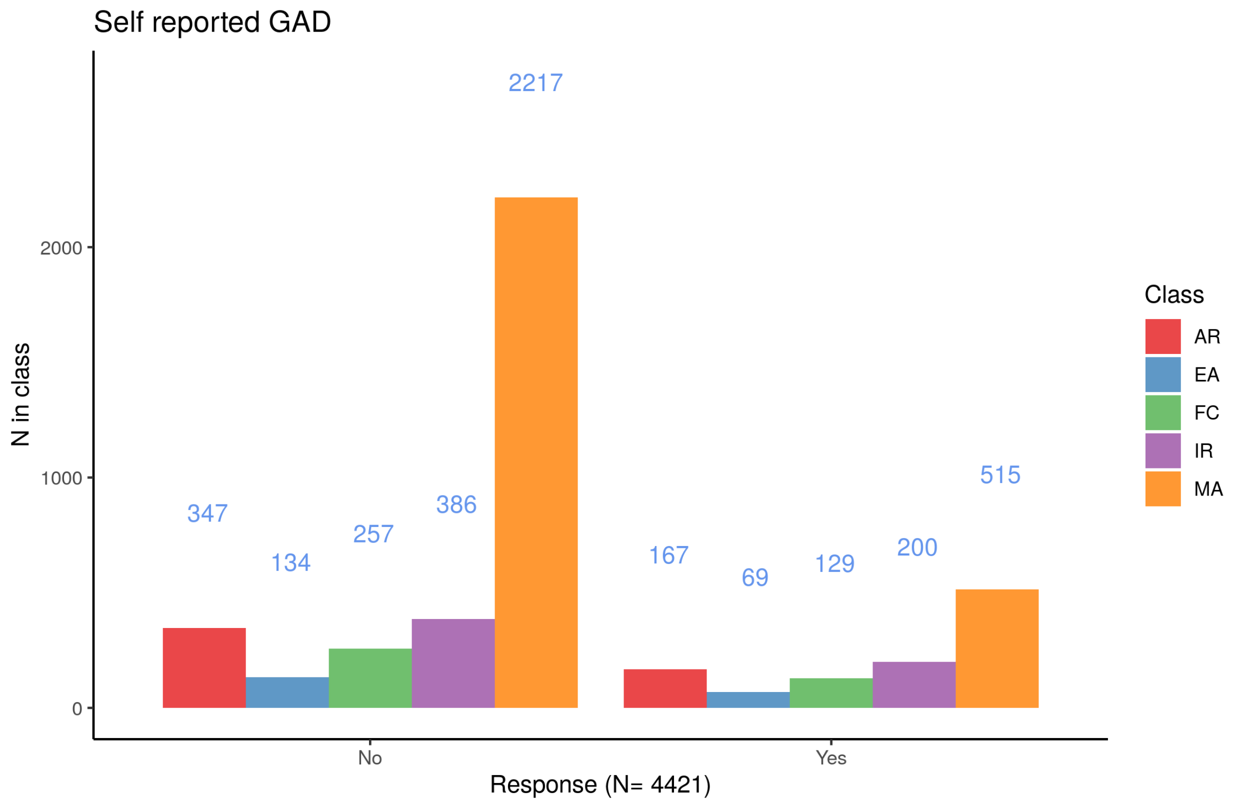

Figure S40 Distributions of self-reported GAD by most likely class membership.

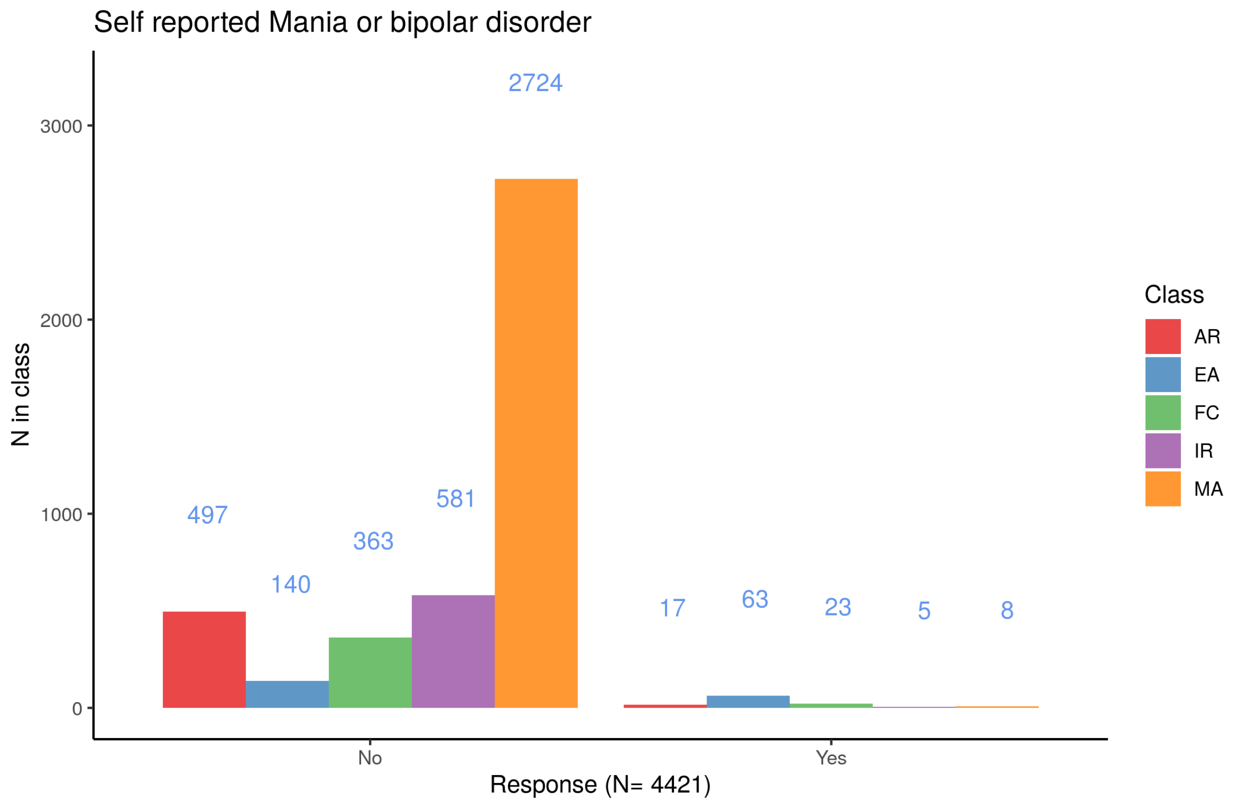


Figure S41 Distributions of self-reported mania or bipolar disorder by most likely class membership.

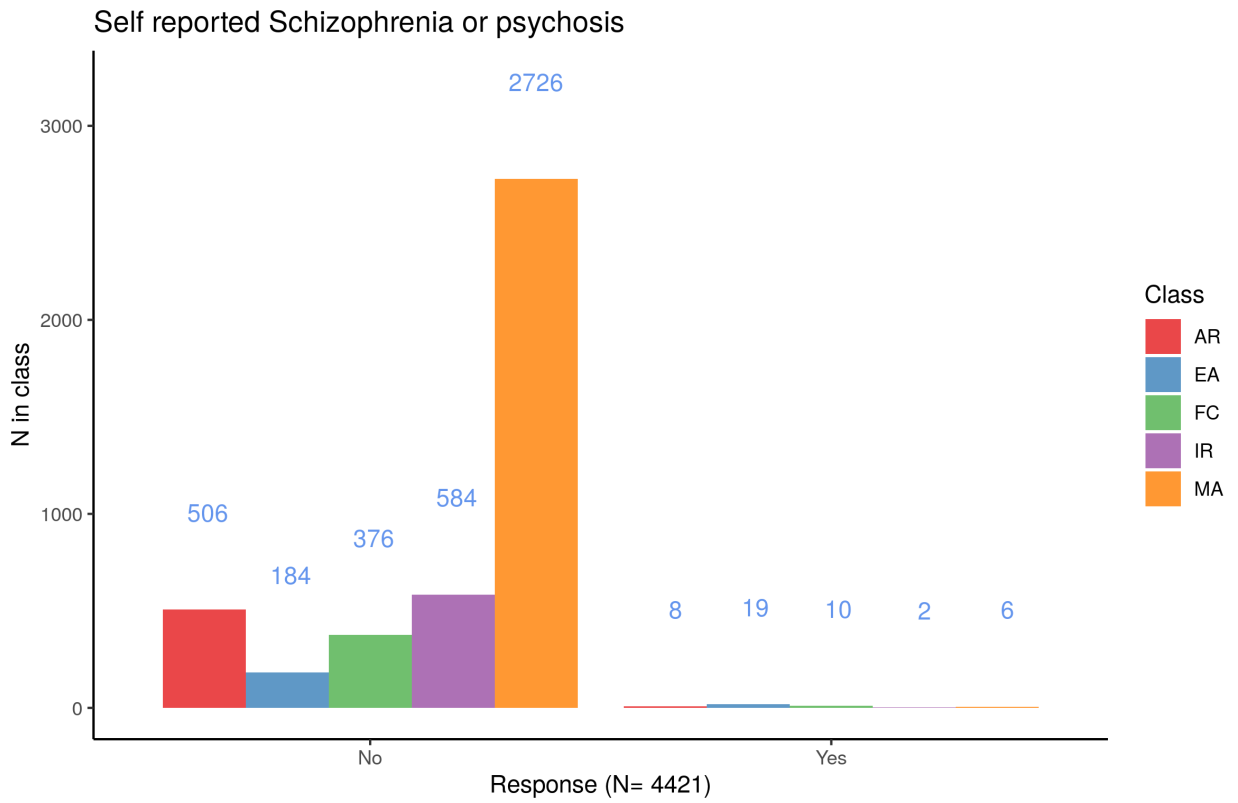


Figure S42 Distributions of self-reported diagnosis of schizophrenia or psychosis by most likely class membership.

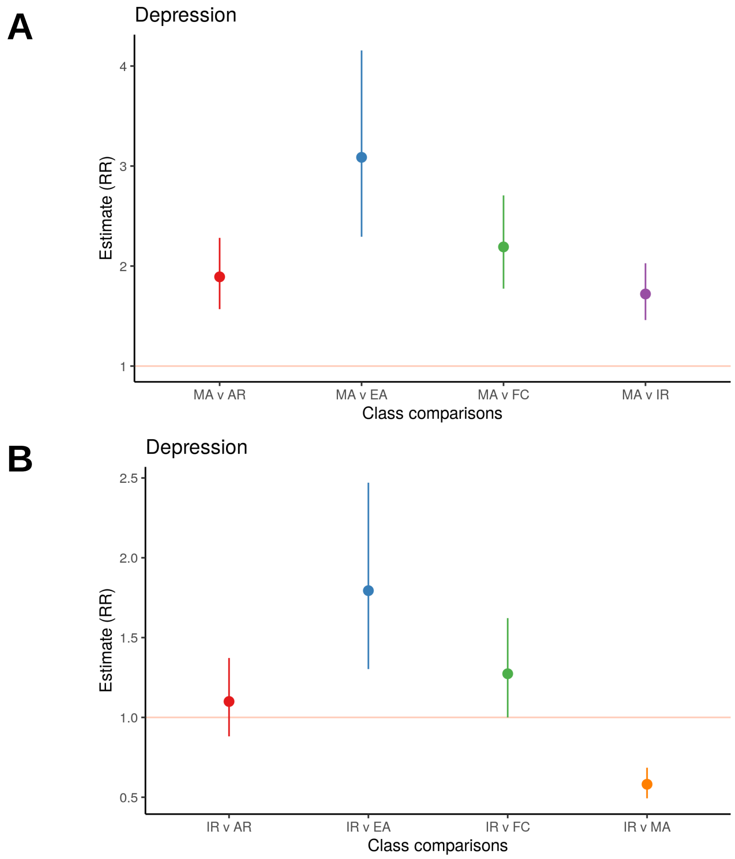


Figure S43 Associations of self-reported dignosis of depression with most likely class membership in the optimum 5-class model in PROTECT.

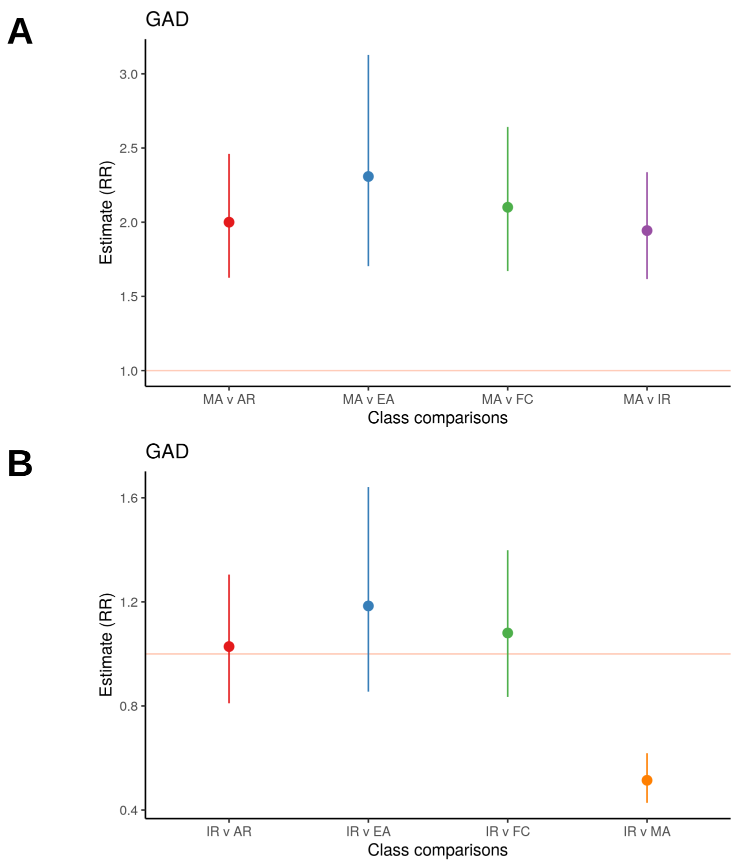


Figure S44 Associations of self-reported diagnosis of GAD with most likely class membership in the optimum 5-class model in PROTECT.

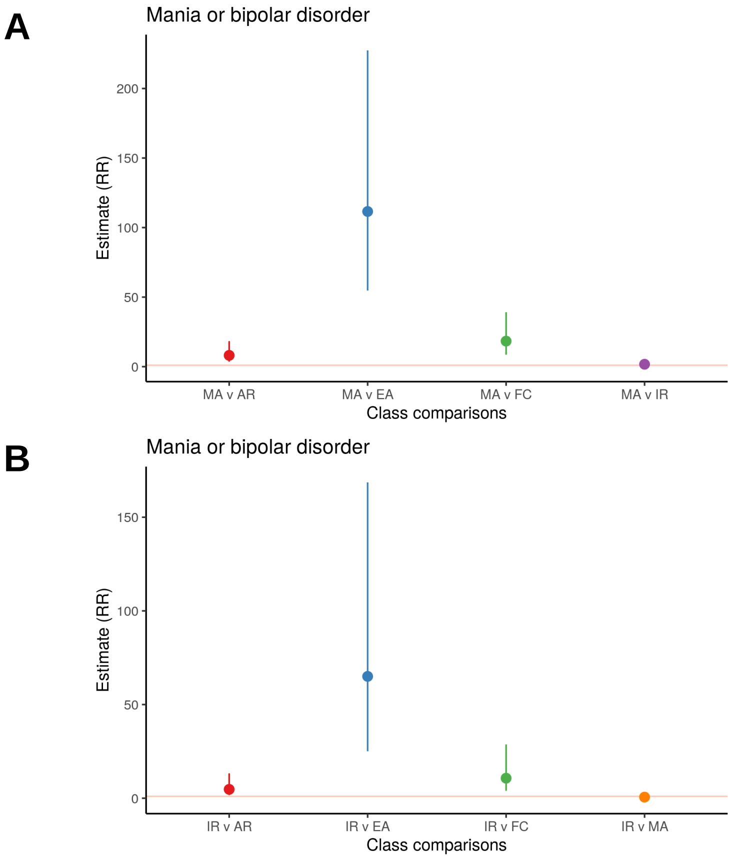


Figure S45 Associations of self-reported diagnosis of mania or bipolar disorder with most likely class membership in the optimum 5-class model in PROTECT.

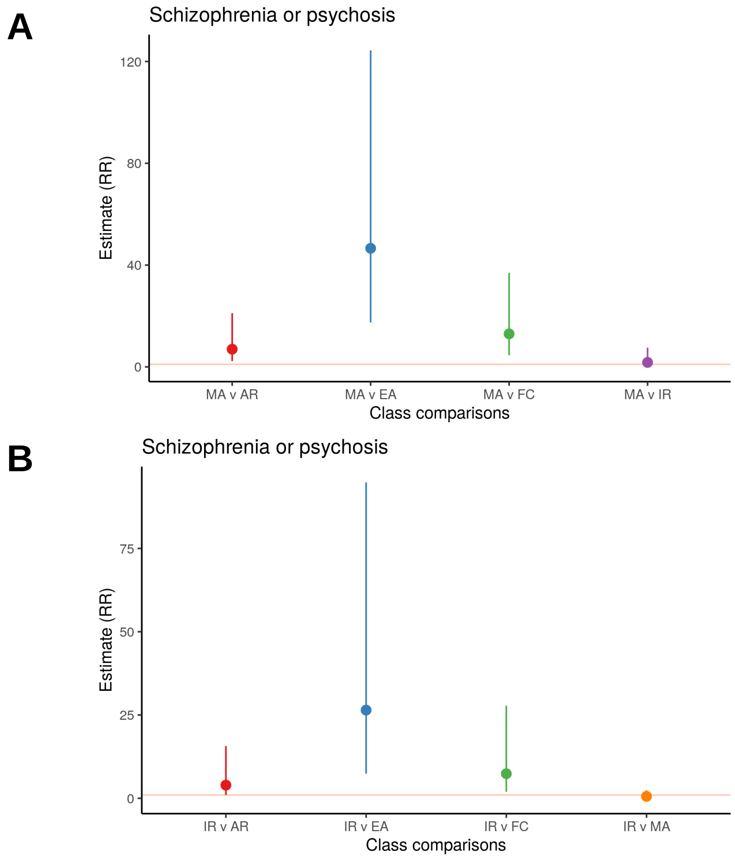


Figure S46 Associations of self-reported diagnosis of schizophrenia or psychosis with most likely class membership in the optimum 5-class model in PROTECT.

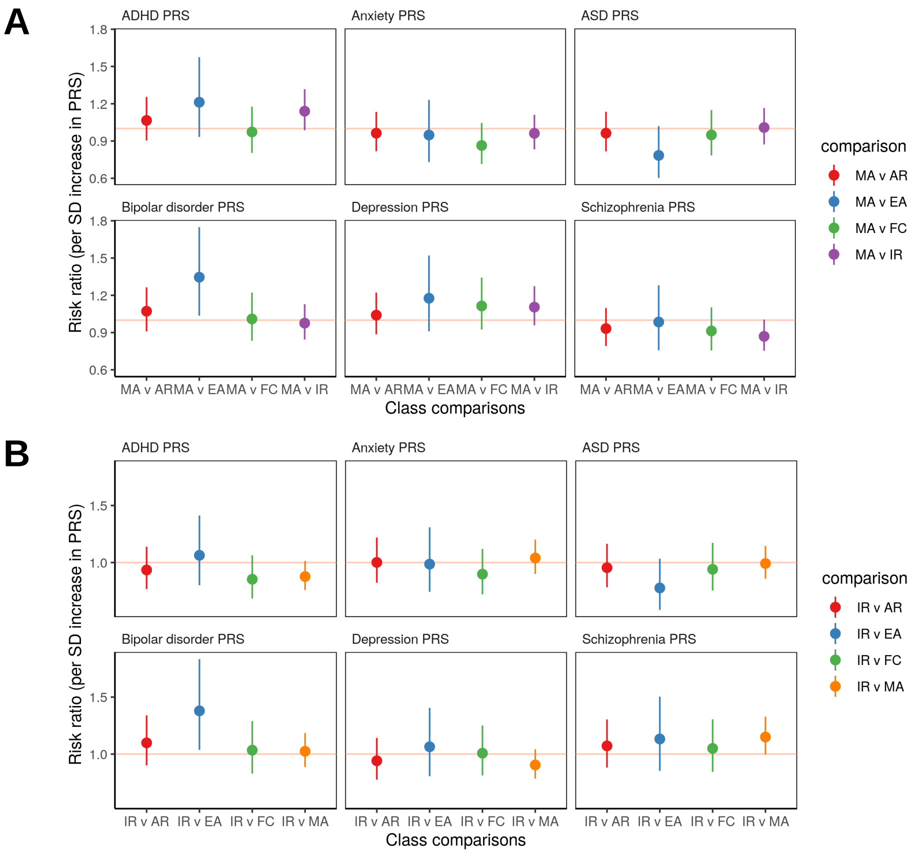


Figure S47 Associations of PRS of six disorders with most likely class membership in the optimum 5-class model in PROTECT.

### 4 Supplementary Tables

Table 1: Details of published genome-wide association studies from which summary results were used for calculating polygenic risk scores (PRS)

| Trait | Publication year | Study population ancestry | Trait data type | N cases | N controls | Sample size (discovery) | PMID |
|---|---|---|---|---|---|---|---|
| ADHD | 2017 | European | binary | 19099 | 34194 | 53293 | 30478444 |
| Anxiety disorders | 2016 | European | binary | NULL | NULL | 17310 | 26754954 |
| Autism spectrum disorder | 2017 | European | binary | 6197 | 7377 | 13574 | 28540026 |
| Bipolar disorder | 2011 | European | binary | 7481 | 9250 | 16731 | 21926972 |
| Major depressive disorder | 2018 | European | binary | 45591 | 97674 | 143265 | 29700475 |
| Schizophrenia | 2014 | European | binary | 33640 | 43456 | 77096 | 25056061 |

*Where sample size for cases and controls is NULL the sample size was not reported or was not available for the subset of the data that was analysed in the original study.*

Table 2: Numbers of responses to individual symptoms questions out of a total sample of N=42183

| Response | Value | N (percent) |
|---|---|---|
| More active | No | 34148 (0.81) |
| More active | Yes | 8035 (0.19) |
| More confident | No | 37526 (0.89) |
| More confident | Yes | 4657 (0.11) |
| Easily distracted | No | 29620 (0.7) |
| Easily distracted | Yes | 12563 (0.3) |
| More creative | No | 37770 (0.9) |
| More creative | Yes | 4413 (0.1) |
| Less sleep | No | 36801 (0.87) |
| Less sleep | Yes | 5382 (0.13) |
| Thoughts racing | No | 27772 (0.66) |
| Thoughts racing | Yes | 14411 (0.34) |
| More restless | No | 24729 (0.59) |
| More restless | Yes | 17454 (0.41) |
| More talkative | No | 34547 (0.82) |
| More talkative | Yes | 7636 (0.18) |

Table 3: Fit statistics for latent class models with 2:7 classes

| Number of classes | 2 | 3 | 4 | 5 | 6 | 7 |
|---|---|---|---|---|---|---|
| BIC | 300578.9 | 291581.9 | 288859.6 | 287883.9 | 287770.8 | 287732.7 |
| AIC | 300431.9 | 291357 | 288556.9 | 287503.4 | 287312.4 | 287196.4 |
| loglikelihood | -150198.947 | -145652.482 | -144243.426 | -143707.676 | -143603.199 | -143536.199 |
| N iterations | 86 | 198 | 187 | 686 | 5140 | 1892 |
| Entropy | 3.58 | 3.46 | 3.43 | 3.41 | 3.41 | 3.4 |
| Relative entropy | 0.761 | 0.665 | 0.665 | 0.626 | 0.629 | 0.592 |
| N | 42183 | 42183 | 42183 | 42183 | 42183 | 42183 |

Table 4: Distribution of responses to manic or irritable stem question by most likely class membership in the optimum 5-class model

| Class | Response | N (percent) |
|---|---|---|
| IR | Ever manic | 229 (1.86) |
| IR | Ever irritable | 11366 (92.47) |
| IR | Ever manic and irritable | 697 (5.67) |
| EA | Ever manic | 260 (20.11) |
| EA | Ever irritable | 226 (17.48) |
| EA | Ever manic and irritable | 807 (62.41) |
| MA | Ever manic | 366 (1.78) |
| MA | Ever irritable | 19670 (95.52) |
| MA | Ever manic and irritable | 556 (2.7) |
| FC | Ever manic | 940 (23.79) |
| FC | Ever irritable | 1629 (41.22) |
| FC | Ever manic and irritable | 1383 (34.99) |
| AR | Ever manic | 408 (10.06) |
| AR | Ever irritable | 2562 (63.2) |
| AR | Ever manic and irritable | 1084 (26.74) |

Table 5: Distribution of responses to the episode duration question by most likely class membership in the optimum 5-class model

| Class | Response | N (percent) |
|---|---|---|
| IR | 1 - Brief | 3417 (27.8) |
| IR | 2 - Moderate | 4878 (39.68) |
| IR | 3 - Extended | 2920 (23.76) |
| IR | Missing - NA | 1077 (8.76) |
| EA | 1 - Brief | 172 (13.3) |
| EA | 2 - Moderate | 493 (38.13) |
| EA | 3 - Extended | 560 (43.31) |
| EA | Missing - NA | 68 (5.26) |
| MA | 1 - Brief | 8303 (40.32) |
| MA | 2 - Moderate | 5915 (28.72) |
| MA | 3 - Extended | 3261 (15.84) |
| MA | Missing - NA | 3113 (15.12) |
| FC | 1 - Brief | 1032 (26.11) |
| FC | 2 - Moderate | 1654 (41.85) |
| FC | 3 - Extended | 1040 (26.32) |
| FC | Missing - NA | 226 (5.72) |
| AR | 1 - Brief | 957 (23.61) |
| AR | 2 - Moderate | 1784 (44.01) |
| AR | 3 - Extended | 1038 (25.6) |
| AR | Missing - NA | 275 (6.78) |

Table 6: Associations of episode duration with most likely class membership weighted for the probability of inclusion in each class.

| Response | Class comparison | Effect | 95% CI | SE | P-value |
|---|---|---|---|---|---|
| Brief duration mania or irritability | MA v IR | 0.541 | 0.515,0.568 | 0.025 | 4.56e-132 |
| Brief duration mania or irritability | MA v EA | 0.192 | 0.164,0.225 | 0.081 | 3.62e-92 |
| Brief duration mania or irritability | MA v FC | 0.435 | 0.403,0.47 | 0.039 | 1.49e-100 |
| Brief duration mania or irritability | MA v AR | 0.404 | 0.375,0.434 | 0.037 | 7.58e-131 |
| Moderate duration mania or irritability | MA v IR | 1.427 | 1.358,1.498 | 0.025 | 6.35e-46 |
| Moderate duration mania or irritability | MA v EA | 1.319 | 1.173,1.483 | 0.060 | 3.82e-06 |
| Moderate duration mania or irritability | MA v FC | 1.529 | 1.423,1.642 | 0.036 | 2.78e-31 |
| Moderate duration mania or irritability | MA v AR | 1.686 | 1.576,1.803 | 0.034 | 2.01e-52 |
| Extended duration mania or irritability | MA v IR | 1.447 | 1.366,1.533 | 0.029 | 2.03e-36 |
| Extended duration mania or irritability | MA v EA | 3.542 | 3.148,3.986 | 0.060 | 7.83e-98 |
| Extended duration mania or irritability | MA v FC | 1.682 | 1.551,1.824 | 0.041 | 1.91e-36 |
| Extended duration mania or irritability | MA v AR | 1.604 | 1.485,1.732 | 0.039 | 2.74e-33 |
| Brief duration mania or irritability | IR v EA | 0.355 | 0.303,0.417 | 0.082 | 9.72e-37 |
| Brief duration mania or irritability | IR v MA | 1.848 | 1.759,1.941 | 0.025 | 4.89e-132 |
| Brief duration mania or irritability | IR v FC | 0.804 | 0.742,0.87 | 0.041 | 7.61e-08 |
| Brief duration mania or irritability | IR v AR | 0.746 | 0.692,0.805 | 0.039 | 4.83e-14 |
| Moderate duration mania or irritability | IR v EA | 0.924 | 0.821,1.04 | 0.060 | 1.93e-01 |
| Moderate duration mania or irritability | IR v MA | 0.701 | 0.668,0.736 | 0.025 | 6.49e-46 |
| Moderate duration mania or irritability | IR v FC | 1.072 | 0.996,1.153 | 0.037 | 6.27e-02 |
| Moderate duration mania or irritability | IR v AR | 1.182 | 1.103,1.266 | 0.035 | 1.87e-06 |
| Extended duration mania or irritability | IR v EA | 2.447 | 2.174,2.755 | 0.060 | 1.22e-49 |
| Extended duration mania or irritability | IR v MA | 0.691 | 0.652,0.732 | 0.029 | 2.00e-36 |
| Extended duration mania or irritability | IR v FC | 1.162 | 1.071,1.261 | 0.041 | 2.95e-04 |
| Extended duration mania or irritability | IR v AR | 1.108 | 1.025,1.197 | 0.040 | 9.48e-03 |

Table 7: Distribution of responses to episode disruptiveness question by most likely class membership in the optimum 5-class model

| Class | Response | N (percent) |
|---|---|---|
| IR | Disruptive | 4018 (39.13) |
| IR | Not disruptive | 6251 (60.87) |
| IR | Missing - NA | 2023 (NA) |
| EA | Disruptive | 611 (53.18) |
| EA | Not disruptive | 538 (46.82) |
| EA | Missing - NA | 144 (NA) |
| MA | Disruptive | 3898 (22.34) |
| MA | Not disruptive | 13549 (77.66) |
| MA | Missing - NA | 3145 (NA) |
| FC | Disruptive | 745 (20.64) |
| FC | Not disruptive | 2865 (79.36) |
| FC | Missing - NA | 342 (NA) |
| AR | Disruptive | 1362 (39.38) |
| AR | Not disruptive | 2097 (60.62) |
| AR | Missing - NA | 595 (NA) |

Table 8: Associations of episode disruptiveness with most likely class membership weighted for the probability of inclusion in each class.

| Response | Class comparison | Effect | 95% CI | SE | P-value |
|---|---|---|---|---|---|
| Problematic mania or irritability | MA v IR | 1.938 | 1.837,2.045 | 0.027 | 1.01e-128 |
| Problematic mania or irritability | MA v EA | 3.675 | 3.259,4.144 | 0.061 | 3.19e-100 |
| Problematic mania or irritability | MA v FC | 0.967 | 0.887,1.053 | 0.044 | 4.40e-01 |
| Problematic mania or irritability | MA v AR | 2.156 | 2.004,2.319 | 0.037 | 8.85e-95 |
| Problematic mania or irritability | IR v EA | 1.895 | 1.68,2.138 | 0.061 | 2.28e-25 |
| Problematic mania or irritability | IR v MA | 0.516 | 0.489,0.544 | 0.027 | 1.06e-128 |
| Problematic mania or irritability | IR v FC | 0.499 | 0.458,0.544 | 0.044 | 3.23e-56 |
| Problematic mania or irritability | IR v AR | 1.112 | 1.033,1.197 | 0.037 | 4.53e-03 |

Table 9: Distribution of sex by most likely class membership in the optimum 5-class model

| Class | Response | N (percent) |
|---|---|---|
| IR | Female | 6916 (56.95) |
| IR | Male | 5229 (43.05) |
| EA | Female | 660 (51.76) |
| EA | Male | 615 (48.24) |
| MA | Female | 12293 (60.55) |
| MA | Male | 8009 (39.45) |
| FC | Female | 1819 (46.72) |
| FC | Male | 2074 (53.28) |
| AR | Female | 2377 (59.35) |
| AR | Male | 1628 (40.65) |

Table 10: Associations of sex with most likely class membership weighted for the probability of inclusion in each class.

| Response | Class comparison | Effect | 95% CI | SE | P-value |
|---|---|---|---|---|---|
| Sex | MA v IR | 1.184 | 1.131,1.239 | 0.023 | 4.01e-13 |
| Sex | MA v EA | 1.460 | 1.305,1.634 | 0.057 | 3.87e-11 |
| Sex | MA v FC | 1.708 | 1.595,1.829 | 0.035 | 2.95e-53 |
| Sex | MA v AR | 1.135 | 1.064,1.211 | 0.033 | 1.16e-04 |
| Sex | IR v EA | 1.234 | 1.101,1.382 | 0.058 | 2.91e-04 |
| Sex | IR v MA | 0.845 | 0.807,0.884 | 0.023 | 4.01e-13 |
| Sex | IR v FC | 1.443 | 1.345,1.549 | 0.036 | 2.12e-24 |
| Sex | IR v AR | 0.959 | 0.897,1.026 | 0.034 | 2.24e-01 |

Table 11: Distribution of educational attainment by most likely class membership in the optimum 5-class model

| Class | Response | N (percent) |
|---|---|---|
| IR | A levels NVQ HNC or HND | 4350 (35.82) |
| IR | None | 752 (6.19) |
| IR | O levels or CSE | 1826 (15.03) |
| IR | University degree | 5217 (42.96) |
| EA | A levels NVQ HNC or HND | 389 (30.51) |
| EA | None | 54 (4.24) |
| EA | O levels or CSE | 166 (13.02) |
| EA | University degree | 666 (52.24) |
| MA | A levels NVQ HNC or HND | 6786 (33.43) |
| MA | None | 1393 (6.86) |
| MA | O levels or CSE | 2829 (13.93) |
| MA | University degree | 9294 (45.78) |
| FC | A levels NVQ HNC or HND | 1317 (33.83) |
| FC | None | 210 (5.39) |
| FC | O levels or CSE | 483 (12.41) |
| FC | University degree | 1883 (48.37) |
| AR | A levels NVQ HNC or HND | 1499 (37.43) |
| AR | None | 227 (5.67) |
| AR | O levels or CSE | 571 (14.26) |
| AR | University degree | 1708 (42.65) |

Table 12: Associations of educational attainment with most likely class membership weighted for the probability of inclusion in each class.

| Response | Class comparison | Effect | 95% CI | SE | P-value |
|---|---|---|---|---|---|
| A levels NVQ HNC or HND | MA v IR | 1.113 | 1.062,1.167 | 0.024 | 7.80e-06 |
| A levels NVQ HNC or HND | MA v EA | 0.883 | 0.782,0.997 | 0.062 | 4.37e-02 |
| A levels NVQ HNC or HND | MA v FC | 1.039 | 0.967,1.116 | 0.037 | 2.98e-01 |
| A levels NVQ HNC or HND | MA v AR | 1.159 | 1.085,1.239 | 0.034 | 1.24e-05 |
| None | MA v IR | 0.994 | 0.908,1.088 | 0.046 | 9.02e-01 |
| None | MA v EA | 0.642 | 0.489,0.843 | 0.139 | 1.41e-03 |
| None | MA v FC | 0.817 | 0.705,0.947 | 0.075 | 7.13e-03 |
| None | MA v AR | 0.910 | 0.797,1.039 | 0.068 | 1.62e-01 |
| O levels or CSE | MA v IR | 1.120 | 1.051,1.194 | 0.032 | 4.66e-04 |
| O levels or CSE | MA v EA | 0.926 | 0.784,1.094 | 0.085 | 3.66e-01 |
| O levels or CSE | MA v FC | 0.891 | 0.805,0.987 | 0.052 | 2.73e-02 |
| O levels or CSE | MA v AR | 1.065 | 0.973,1.167 | 0.046 | 1.72e-01 |
| University degree | MA v IR | 0.858 | 0.82,0.897 | 0.023 | 2.52e-11 |
| University degree | MA v EA | 1.265 | 1.131,1.415 | 0.057 | 3.97e-05 |
| University degree | MA v FC | 1.066 | 0.996,1.141 | 0.035 | 6.41e-02 |
| University degree | MA v AR | 0.865 | 0.812,0.923 | 0.033 | 1.02e-05 |
| A levels NVQ HNC or HND | IR v EA | 0.793 | 0.701,0.896 | 0.063 | 2.08e-04 |
| A levels NVQ HNC or HND | IR v MA | 0.898 | 0.857,0.941 | 0.024 | 7.82e-06 |
| A levels NVQ HNC or HND | IR v FC | 0.933 | 0.866,1.005 | 0.038 | 6.60e-02 |
| A levels NVQ HNC or HND | IR v AR | 1.041 | 0.972,1.115 | 0.035 | 2.47e-01 |
| None | IR v EA | 0.646 | 0.491,0.849 | 0.140 | 1.78e-03 |
| None | IR v MA | 1.006 | 0.919,1.101 | 0.046 | 9.02e-01 |
| None | IR v FC | 0.821 | 0.706,0.956 | 0.078 | 1.11e-02 |
| None | IR v AR | 0.915 | 0.797,1.05 | 0.070 | 2.05e-01 |
| O levels or CSE | IR v EA | 0.827 | 0.698,0.978 | 0.086 | 2.68e-02 |
| O levels or CSE | IR v MA | 0.893 | 0.838,0.951 | 0.032 | 4.65e-04 |
| O levels or CSE | IR v FC | 0.795 | 0.716,0.883 | 0.053 | 1.79e-05 |
| O levels or CSE | IR v AR | 0.951 | 0.866,1.044 | 0.048 | 2.91e-01 |
| University degree | IR v EA | 1.475 | 1.316,1.652 | 0.058 | 2.07e-11 |
| University degree | IR v MA | 1.166 | 1.115,1.22 | 0.023 | 2.51e-11 |
| University degree | IR v FC | 1.244 | 1.159,1.334 | 0.036 | 1.40e-09 |
| University degree | IR v AR | 1.009 | 0.944,1.079 | 0.034 | 7.88e-01 |

Table 13: Distribution of alcohol consumption frequency by most likely class membership in the optimum 5-class model

| Class | Response | N (percent) |
|---|---|---|
| IR | Daily | 2538 (20.9) |
| IR | Never | 693 (5.71) |
| IR | Occasionally | 2758 (22.71) |
| IR | Weekly | 6156 (50.69) |
| EA | Daily | 266 (20.86) |
| EA | Never | 123 (9.65) |
| EA | Occasionally | 313 (24.55) |
| EA | Weekly | 573 (44.94) |
| MA | Daily | 4577 (22.54) |
| MA | Never | 1165 (5.74) |
| MA | Occasionally | 4400 (21.67) |
| MA | Weekly | 10160 (50.04) |
| FC | Daily | 953 (24.48) |
| FC | Never | 266 (6.83) |
| FC | Occasionally | 807 (20.73) |
| FC | Weekly | 1867 (47.96) |
| AR | Daily | 804 (20.07) |
| AR | Never | 274 (6.84) |
| AR | Occasionally | 959 (23.95) |
| AR | Weekly | 1968 (49.14) |

Table 14: Associations of alcohol consumption frequency with most likely class membership weighted for the probability of inclusion in each class.

| Response | Class comparison | Effect | 95% CI | SE | P-value |
|---|---|---|---|---|---|
| Daily | MA v IR | 0.907 | 0.859,0.958 | 0.028 | 4.46e-04 |
| Daily | MA v EA | 0.896 | 0.78,1.028 | 0.070 | 1.17e-01 |
| Daily | MA v FC | 1.081 | 0.998,1.171 | 0.041 | 5.59e-02 |
| Daily | MA v AR | 0.885 | 0.819,0.957 | 0.040 | 2.23e-03 |
| Never | MA v IR | 1.019 | 0.925,1.122 | 0.049 | 7.07e-01 |
| Never | MA v EA | 1.726 | 1.419,2.1 | 0.100 | 4.66e-08 |
| Never | MA v FC | 1.222 | 1.065,1.401 | 0.070 | 4.15e-03 |
| Never | MA v AR | 1.230 | 1.081,1.398 | 0.065 | 1.61e-03 |
| Occasionally | MA v IR | 1.056 | 1,1.114 | 0.028 | 4.91e-02 |
| Occasionally | MA v EA | 1.182 | 1.037,1.346 | 0.067 | 1.22e-02 |
| Occasionally | MA v FC | 0.980 | 0.902,1.065 | 0.042 | 6.39e-01 |
| Occasionally | MA v AR | 1.124 | 1.042,1.212 | 0.038 | 2.40e-03 |
| Weekly | MA v IR | 1.026 | 0.981,1.073 | 0.023 | 2.69e-01 |
| Weekly | MA v EA | 0.825 | 0.737,0.923 | 0.057 | 7.78e-04 |
| Weekly | MA v FC | 0.915 | 0.855,0.979 | 0.035 | 1.02e-02 |
| Weekly | MA v AR | 0.954 | 0.895,1.016 | 0.032 | 1.44e-01 |
| Daily | IR v EA | 0.987 | 0.858,1.135 | 0.071 | 8.57e-01 |
| Daily | IR v MA | 1.102 | 1.044,1.164 | 0.028 | 4.42e-04 |
| Daily | IR v FC | 1.192 | 1.096,1.295 | 0.042 | 3.66e-05 |
| Daily | IR v AR | 0.976 | 0.899,1.059 | 0.042 | 5.57e-01 |
| Never | IR v EA | 1.695 | 1.389,2.07 | 0.102 | 2.16e-07 |
| Never | IR v MA | 0.982 | 0.891,1.081 | 0.049 | 7.09e-01 |
| Never | IR v FC | 1.200 | 1.041,1.383 | 0.073 | 1.22e-02 |
| Never | IR v AR | 1.207 | 1.056,1.38 | 0.068 | 5.86e-03 |
| Occasionally | IR v EA | 1.119 | 0.98,1.277 | 0.067 | 9.54e-02 |
| Occasionally | IR v MA | 0.947 | 0.897,1 | 0.028 | 4.90e-02 |
| Occasionally | IR v FC | 0.929 | 0.853,1.012 | 0.044 | 8.98e-02 |
| Occasionally | IR v AR | 1.065 | 0.985,1.151 | 0.040 | 1.16e-01 |
| Weekly | IR v EA | 0.804 | 0.718,0.901 | 0.058 | 1.76e-04 |
| Weekly | IR v MA | 0.975 | 0.932,1.02 | 0.023 | 2.69e-01 |
| Weekly | IR v FC | 0.892 | 0.831,0.957 | 0.036 | 1.44e-03 |
| Weekly | IR v AR | 0.930 | 0.87,0.994 | 0.034 | 3.13e-02 |

Table 15: Distribution of smoking by most likely class membership in the optimum 5-class model

| Class | Response | N (percent) |
|---|---|---|
| IR | Current | 847 (6.97) |
| IR | Never | 6561 (54.02) |
| IR | Past | 4737 (39) |
| EA | Current | 156 (12.24) |
| EA | Never | 568 (44.55) |
| EA | Past | 551 (43.22) |
| MA | Current | 1140 (5.62) |
| MA | Never | 10823 (53.31) |
| MA | Past | 8339 (41.07) |
| FC | Current | 329 (8.45) |
| FC | Never | 1910 (49.06) |
| FC | Past | 1654 (42.49) |
| AR | Current | 364 (9.09) |
| AR | Never | 1973 (49.26) |
| AR | Past | 1668 (41.65) |

Table 16: Associations of smoking with most likely class membership weighted for the probability of inclusion in each class.

| Response | Class comparison | Effect | 95% CI | SE | P-value |
|---|---|---|---|---|---|
| Current | MA v IR | 1.219 | 1.111,1.338 | 0.047 | 2.75e-05 |
| Current | MA v EA | 2.379 | 1.995,2.837 | 0.090 | 4.52e-22 |
| Current | MA v FC | 1.536 | 1.352,1.746 | 0.065 | 4.22e-11 |
| Current | MA v AR | 1.612 | 1.432,1.814 | 0.060 | 2.78e-15 |
| Never | MA v IR | 1.017 | 0.973,1.064 | 0.023 | 4.52e-01 |
| Never | MA v EA | 0.708 | 0.632,0.792 | 0.057 | 1.75e-09 |
| Never | MA v FC | 0.853 | 0.797,0.913 | 0.035 | 4.28e-06 |
| Never | MA v AR | 0.864 | 0.811,0.921 | 0.032 | 6.83e-06 |
| Past | MA v IR | 0.936 | 0.895,0.98 | 0.023 | 4.91e-03 |
| Past | MA v EA | 1.079 | 0.963,1.208 | 0.058 | 1.89e-01 |
| Past | MA v FC | 1.052 | 0.982,1.127 | 0.035 | 1.51e-01 |
| Past | MA v AR | 1.022 | 0.958,1.09 | 0.033 | 5.17e-01 |
| Current | IR v EA | 1.951 | 1.634,2.331 | 0.091 | 1.69e-13 |
| Current | IR v MA | 0.820 | 0.747,0.9 | 0.047 | 2.76e-05 |
| Current | IR v FC | 1.260 | 1.106,1.435 | 0.066 | 4.99e-04 |
| Current | IR v AR | 1.322 | 1.171,1.492 | 0.062 | 6.49e-06 |
| Never | IR v EA | 0.696 | 0.621,0.78 | 0.058 | 4.37e-10 |
| Never | IR v MA | 0.983 | 0.94,1.028 | 0.023 | 4.51e-01 |
| Never | IR v FC | 0.838 | 0.781,0.899 | 0.036 | 8.53e-07 |
| Never | IR v AR | 0.849 | 0.795,0.907 | 0.034 | 1.32e-06 |
| Past | IR v EA | 1.152 | 1.027,1.293 | 0.059 | 1.54e-02 |
| Past | IR v MA | 1.068 | 1.02,1.118 | 0.023 | 4.91e-03 |
| Past | IR v FC | 1.123 | 1.046,1.206 | 0.036 | 1.44e-03 |
| Past | IR v AR | 1.091 | 1.02,1.167 | 0.034 | 1.13e-02 |

Table 17: Associations of Townsend deprivation index (TDI) with most likely class membership weighted for the probability of inclusion in each class.

| Response | Class comparison | Effect | 95% CI | SE | P-value |
|---|---|---|---|---|---|
| Townsend deprivation rank normalised | MA v IR | 1.027 | 1.004,1.05 | 0.011 | 2.23e-02 |
| Townsend deprivation rank normalised | MA v EA | 1.313 | 1.242,1.389 | 0.029 | 2.02e-21 |
| Townsend deprivation rank normalised | MA v FC | 1.111 | 1.074,1.149 | 0.017 | 1.39e-09 |
| Townsend deprivation rank normalised | MA v AR | 1.083 | 1.049,1.118 | 0.016 | 9.75e-07 |
| Townsend deprivation rank normalised | IR v EA | 1.279 | 1.209,1.354 | 0.029 | 2.16e-17 |
| Townsend deprivation rank normalised | IR v MA | 0.974 | 0.953,0.996 | 0.011 | 2.22e-02 |
| Townsend deprivation rank normalised | IR v FC | 1.082 | 1.045,1.121 | 0.018 | 1.08e-05 |
| Townsend deprivation rank normalised | IR v AR | 1.055 | 1.021,1.09 | 0.017 | 1.56e-03 |

Table 18: Numbers of cases of self-reported diagnoses of six disorders by most likely class membership.

| Class | Outcome | Response | N (percent) |
|---|---|---|---|
| IR | ADHD | Case | 18 (0.15) |
| EA | ADHD | Case | 8 (0.62) |
| MA | ADHD | Case | 19 (0.09) |
| FC | ADHD | Case | 11 (0.28) |
| AR | ADHD | Case | 21 (0.52) |
| IR | GAD | Case | 3282 (26.7) |
| EA | GAD | Case | 478 (36.97) |
| MA | GAD | Case | 3398 (16.5) |
| FC | GAD | Case | 851 (21.53) |
| AR | GAD | Case | 1362 (33.6) |
| IR | ASD | Case | 43 (0.35) |
| EA | ASD | Case | 18 (1.39) |
| MA | ASD | Case | 41 (0.2) |
| FC | ASD | Case | 16 (0.4) |
| AR | ASD | Case | 26 (0.64) |
| IR | Mania or bipolar | Case | 51 (0.41) |
| EA | Mania or bipolar | Case | 339 (26.22) |
| MA | Mania or bipolar | Case | 44 (0.21) |
| FC | Mania or bipolar | Case | 145 (3.67) |
| AR | Mania or bipolar | Case | 139 (3.43) |
| IR | Depression | Case | 5020 (40.84) |
| EA | Depression | Case | 698 (53.98) |
| MA | Depression | Case | 5698 (27.67) |
| FC | Depression | Case | 1468 (37.15) |
| AR | Depression | Case | 1877 (46.3) |
| IR | Schizophrenia or psychosis | Case | 80 (0.65) |
| EA | Schizophrenia or psychosis | Case | 146 (11.29) |
| MA | Schizophrenia or psychosis | Case | 67 (0.33) |
| FC | Schizophrenia or psychosis | Case | 75 (1.9) |
| AR | Schizophrenia or psychosis | Case | 98 (2.42) |

Table 19: Associations of self-reported diagnoses of six disorders with most likely class membership weighted for the probability of inclusion in each class.

| Response | Class comparison | Effect | 95% CI | SE | P-value |
|---|---|---|---|---|---|
| Self reported ADHD | MA v IR | 1.411 | 0.738,2.697 | 0.331 | 2.98e-01 |
| Self reported ADHD | MA v EA | 6.973 | 3.135,15.511 | 0.408 | 1.93e-06 |
| Self reported ADHD | MA v FC | 2.852 | 1.363,5.968 | 0.377 | 5.41e-03 |
| Self reported ADHD | MA v AR | 4.172 | 2.206,7.892 | 0.325 | 1.12e-05 |
| Self reported GAD | MA v IR | 1.698 | 1.607,1.795 | 0.028 | 3.01e-79 |
| Self reported GAD | MA v EA | 2.918 | 2.593,3.284 | 0.060 | 1.23e-70 |
| Self reported GAD | MA v FC | 1.434 | 1.32,1.558 | 0.042 | 1.86e-17 |
| Self reported GAD | MA v AR | 2.385 | 2.22,2.562 | 0.037 | 7.24e-125 |
| Self reported ASD | MA v IR | 1.398 | 0.903,2.163 | 0.223 | 1.33e-01 |
| Self reported ASD | MA v EA | 6.279 | 3.59,10.982 | 0.285 | 1.18e-10 |
| Self reported ASD | MA v FC | 1.928 | 1.097,3.391 | 0.288 | 2.26e-02 |
| Self reported ASD | MA v AR | 2.660 | 1.641,4.314 | 0.247 | 7.27e-05 |
| Self reported Mania or bipolar disorder | MA v IR | 1.793 | 1.191,2.7 | 0.209 | 5.19e-03 |
| Self reported Mania or bipolar disorder | MA v EA | 153.661 | 110.369,213.934 | 0.169 | 2.17e-195 |
| Self reported Mania or bipolar disorder | MA v FC | 17.048 | 12.043,24.135 | 0.177 | 1.46e-57 |
| Self reported Mania or bipolar disorder | MA v AR | 13.219 | 9.309,18.771 | 0.179 | 3.32e-47 |
| Self reported Depression | MA v IR | 1.571 | 1.498,1.647 | 0.024 | 7.65e-78 |
| Self reported Depression | MA v EA | 3.037 | 2.714,3.399 | 0.057 | 2.25e-83 |
| Self reported Depression | MA v FC | 1.516 | 1.412,1.627 | 0.036 | 9.08e-31 |
| Self reported Depression | MA v AR | 2.091 | 1.96,2.232 | 0.033 | 9.28e-110 |
| Self reported Schizophrenia or psychosis | MA v IR | 1.792 | 1.278,2.513 | 0.173 | 7.24e-04 |
| Self reported Schizophrenia or psychosis | MA v EA | 37.778 | 27.805,51.329 | 0.156 | 2.68e-119 |
| Self reported Schizophrenia or psychosis | MA v FC | 6.111 | 4.366,8.552 | 0.172 | 4.93e-26 |
| Self reported Schizophrenia or psychosis | MA v AR | 6.701 | 4.867,9.227 | 0.163 | 2.03e-31 |

Table 20: Associations of self-reported diagnoses of six disorders with most likely class membership weighted for the probability of inclusion in each class.

| Response | Class comparison | Effect | 95% CI | SE | P-value |
|---|---|---|---|---|---|
| Self reported ADHD | IR v EA | 4.972 | 2.244,11.015 | 0.406 | 7.78e-05 |
| Self reported ADHD | IR v MA | 0.707 | 0.37,1.351 | 0.331 | 2.93e-01 |
| Self reported ADHD | IR v FC | 2.008 | 0.961,4.199 | 0.376 | 6.38e-02 |
| Self reported ADHD | IR v AR | 2.951 | 1.564,5.568 | 0.324 | 8.38e-04 |
| Self reported GAD | IR v EA | 1.718 | 1.526,1.934 | 0.060 | 2.98e-19 |
| Self reported GAD | IR v MA | 0.589 | 0.557,0.622 | 0.028 | 3.12e-79 |
| Self reported GAD | IR v FC | 0.844 | 0.777,0.918 | 0.042 | 6.78e-05 |
| Self reported GAD | IR v AR | 1.404 | 1.307,1.509 | 0.037 | 1.99e-20 |
| Self reported ASD | IR v EA | 4.549 | 2.602,7.953 | 0.285 | 1.06e-07 |
| Self reported ASD | IR v MA | 0.720 | 0.465,1.115 | 0.223 | 1.41e-01 |
| Self reported ASD | IR v FC | 1.402 | 0.798,2.463 | 0.287 | 2.40e-01 |
| Self reported ASD | IR v AR | 1.933 | 1.193,3.133 | 0.246 | 7.46e-03 |
| Self reported Mania or bipolar disorder | IR v EA | 85.812 | 63.709,115.582 | 0.152 | 1.07e-188 |
| Self reported Mania or bipolar disorder | IR v MA | 0.559 | 0.371,0.842 | 0.209 | 5.33e-03 |
| Self reported Mania or bipolar disorder | IR v FC | 9.521 | 6.94,13.062 | 0.161 | 2.51e-44 |
| Self reported Mania or bipolar disorder | IR v AR | 7.382 | 5.363,10.162 | 0.163 | 1.50e-34 |
| Self reported Depression | IR v EA | 1.934 | 1.727,2.166 | 0.058 | 4.37e-30 |
| Self reported Depression | IR v MA | 0.637 | 0.607,0.668 | 0.024 | 7.39e-78 |
| Self reported Depression | IR v FC | 0.965 | 0.898,1.037 | 0.037 | 3.34e-01 |
| Self reported Depression | IR v AR | 1.331 | 1.246,1.423 | 0.034 | 3.13e-17 |
| Self reported Schizophrenia or psychosis | IR v EA | 21.055 | 15.873,27.927 | 0.144 | 3.32e-99 |
| Self reported Schizophrenia or psychosis | IR v MA | 0.558 | 0.398,0.783 | 0.172 | 7.29e-04 |
| Self reported Schizophrenia or psychosis | IR v FC | 3.411 | 2.491,4.671 | 0.160 | 1.99e-14 |
| Self reported Schizophrenia or psychosis | IR v AR | 3.735 | 2.775,5.025 | 0.151 | 3.31e-18 |

Table 21: Numbers of cases of ICD hospital diagnoses of three disorders by most likely class membership.

| Class | Outcome | Response | N (percent) |
|---|---|---|---|
| IR | Mania or bipolar disorder ICD | Case | 22 (0.21) |
| EA | Mania or bipolar disorder ICD | Case | 156 (13.74) |
| MA | Mania or bipolar disorder ICD | Case | 19 (0.11) |
| FC | Mania or bipolar disorder ICD | Case | 66 (1.95) |
| AR | Mania or bipolar disorder ICD | Case | 57 (1.6) |
| IR | Depression ICD | Case | 1032 (9.67) |
| EA | Depression ICD | Case | 205 (18.06) |
| MA | Depression ICD | Case | 998 (5.7) |
| FC | Depression ICD | Case | 347 (10.26) |
| AR | Depression ICD | Case | 455 (12.76) |
| IR | Schizophrenia or psychotic disorder ICD | Case | 28 (0.26) |
| EA | Schizophrenia or psychotic disorder ICD | Case | 39 (3.44) |
| MA | Schizophrenia or psychotic disorder ICD | Case | 21 (0.12) |
| FC | Schizophrenia or psychotic disorder ICD | Case | 24 (0.71) |
| AR | Schizophrenia or psychotic disorder ICD | Case | 29 (0.81) |

Table 22: Associations of ICD hospital diagnoses of three disorders with most likely class membership weighted for the probability of inclusion in each class.

| Response | Class comparison | Effect | 95% CI | SE | P-value |
|---|---|---|---|---|---|
| Mania or bipolar disorder ICD | MA v IR | 1.654 | 0.882,3.099 | 0.320 | 1.16e-01 |
| Mania or bipolar disorder ICD | MA v EA | 136.051 | 83.01,222.982 | 0.252 | 1.33e-84 |
| Mania or bipolar disorder ICD | MA v FC | 17.448 | 10.35,29.412 | 0.266 | 7.24e-27 |
| Mania or bipolar disorder ICD | MA v AR | 11.784 | 6.907,20.105 | 0.273 | 1.43e-19 |
| Depression ICD | MA v IR | 1.592 | 1.452,1.747 | 0.047 | 6.26e-23 |
| Depression ICD | MA v EA | 3.631 | 3.086,4.272 | 0.083 | 1.74e-54 |
| Depression ICD | MA v FC | 1.776 | 1.561,2.021 | 0.066 | 2.47e-18 |
| Depression ICD | MA v AR | 2.308 | 2.061,2.584 | 0.058 | 1.08e-47 |
| Schizophrenia or psychotic disorder ICD | MA v IR | 1.671 | 0.927,3.012 | 0.301 | 8.79e-02 |
| Schizophrenia or psychotic disorder ICD | MA v EA | 26.354 | 15.312,45.357 | 0.277 | 3.47e-32 |
| Schizophrenia or psychotic disorder ICD | MA v FC | 5.968 | 3.347,10.642 | 0.295 | 1.42e-09 |
| Schizophrenia or psychotic disorder ICD | MA v AR | 5.784 | 3.3,10.137 | 0.286 | 8.81e-10 |

Table 23: Associations of ICD hospital diagnoses of three disorders with most likely class membership weighted for the probability of inclusion in each class.

| Response | Class comparison | Effect | 95% CI | SE | P-value |
|---|---|---|---|---|---|
| Mania or bipolar disorder ICD | IR v EA | 82.752 | 52.451,130.556 | 0.233 | 2.42e-80 |
| Mania or bipolar disorder ICD | IR v MA | 0.610 | 0.325,1.143 | 0.320 | 1.23e-01 |
| Mania or bipolar disorder ICD | IR v FC | 10.509 | 6.459,17.099 | 0.248 | 2.78e-21 |
| Mania or bipolar disorder ICD | IR v AR | 7.194 | 4.369,11.848 | 0.255 | 9.00e-15 |
| Depression ICD | IR v EA | 2.280 | 1.94,2.68 | 0.082 | 1.45e-23 |
| Depression ICD | IR v MA | 0.628 | 0.573,0.689 | 0.047 | 6.39e-23 |
| Depression ICD | IR v FC | 1.116 | 0.982,1.267 | 0.065 | 9.28e-02 |
| Depression ICD | IR v AR | 1.449 | 1.296,1.62 | 0.057 | 6.65e-11 |
| Schizophrenia or psychotic disorder ICD | IR v EA | 15.962 | 9.564,26.638 | 0.261 | 2.93e-26 |
| Schizophrenia or psychotic disorder ICD | IR v MA | 0.605 | 0.335,1.093 | 0.301 | 9.58e-02 |
| Schizophrenia or psychotic disorder ICD | IR v FC | 3.643 | 2.105,6.306 | 0.280 | 3.86e-06 |
| Schizophrenia or psychotic disorder ICD | IR v AR | 3.501 | 2.058,5.957 | 0.271 | 3.81e-06 |

Table 24: Associations of self-reported personality disorder with most likely class membership weighted for the probability of inclusion in each class.

| Response | Class comparison | Effect | 95% CI | SE | P-value |
|---|---|---|---|---|---|
| Self reported Personality disorder | MA v IR | 2.175 | 1.525,3.103 | 0.181 | 1.80e-05 |
| Self reported Personality disorder | MA v EA | 11.538 | 7.595,17.527 | 0.213 | 2.00e-30 |
| Self reported Personality disorder | MA v FC | 3.503 | 2.302,5.331 | 0.214 | 4.92e-09 |
| Self reported Personality disorder | MA v AR | 4.165 | 2.833,6.122 | 0.197 | 3.89e-13 |
| Self reported Personality disorder | IR v EA | 5.286 | 3.6,7.761 | 0.196 | 1.95e-17 |
| Self reported Personality disorder | IR v MA | 0.458 | 0.321,0.653 | 0.181 | 1.63e-05 |
| Self reported Personality disorder | IR v FC | 1.604 | 1.09,2.36 | 0.197 | 1.65e-02 |
| Self reported Personality disorder | IR v AR | 1.913 | 1.352,2.709 | 0.177 | 2.53e-04 |

Table 25: Associations of neuroticism score with most likely class membership weighted for the probability of inclusion in each class.

| Response | Class comparison | Effect | 95% CI | SE | P-value |
|---|---|---|---|---|---|
| Neuroticism score | MA v IR | 1.286 | 1.254,1.318 | 0.013 | 3.05e-87 |
| Neuroticism score | MA v EA | 1.672 | 1.571,1.779 | 0.032 | 4.09e-59 |
| Neuroticism score | MA v FC | 1.153 | 1.11,1.197 | 0.019 | 9.91e-14 |
| Neuroticism score | MA v AR | 1.416 | 1.367,1.467 | 0.018 | 3.86e-84 |
| Neuroticism score | IR v EA | 1.300 | 1.221,1.384 | 0.032 | 2.10e-16 |
| Neuroticism score | IR v MA | 0.778 | 0.759,0.797 | 0.013 | 3.04e-87 |
| Neuroticism score | IR v FC | 0.896 | 0.863,0.932 | 0.020 | 2.69e-08 |
| Neuroticism score | IR v AR | 1.101 | 1.062,1.142 | 0.018 | 1.61e-07 |

Table 26: Associations of polygenic risk scores (PRS) of six disorders with most likely class membership weighted for the probability of inclusion in each class.

| Response | Class comparison | Effect | 95% CI | SE | P-value |
|---|---|---|---|---|---|
| Bipolar disorder PRS | MA v IR | 1.000 | 0.975,1.025 | 0.013 | 9.89e-01 |
| Bipolar disorder PRS | MA v EA | 1.172 | 1.101,1.247 | 0.032 | 5.74e-07 |
| Bipolar disorder PRS | MA v FC | 1.078 | 1.038,1.12 | 0.019 | 1.06e-04 |
| Bipolar disorder PRS | MA v AR | 1.014 | 0.979,1.051 | 0.018 | 4.35e-01 |
| Schizophrenia PRS | MA v IR | 0.992 | 0.967,1.017 | 0.013 | 5.18e-01 |
| Schizophrenia PRS | MA v EA | 1.246 | 1.17,1.327 | 0.032 | 7.04e-12 |
| Schizophrenia PRS | MA v FC | 1.109 | 1.068,1.153 | 0.020 | 1.14e-07 |
| Schizophrenia PRS | MA v AR | 1.065 | 1.027,1.103 | 0.018 | 5.88e-04 |
| Depression PRS | MA v IR | 1.022 | 0.997,1.048 | 0.013 | 8.69e-02 |
| Depression PRS | MA v EA | 1.144 | 1.075,1.217 | 0.032 | 2.10e-05 |
| Depression PRS | MA v FC | 1.053 | 1.014,1.093 | 0.019 | 7.73e-03 |
| Depression PRS | MA v AR | 1.045 | 1.009,1.083 | 0.018 | 1.46e-02 |
| ADHD PRS | MA v IR | 1.028 | 1.002,1.054 | 0.013 | 3.21e-02 |
| ADHD PRS | MA v EA | 1.064 | 1,1.132 | 0.032 | 5.06e-02 |
| ADHD PRS | MA v FC | 1.010 | 0.972,1.049 | 0.019 | 6.21e-01 |
| ADHD PRS | MA v AR | 1.051 | 1.014,1.089 | 0.018 | 6.21e-03 |
| ASD PRS | MA v IR | 0.987 | 0.963,1.013 | 0.013 | 3.26e-01 |
| ASD PRS | MA v EA | 1.001 | 0.94,1.065 | 0.032 | 9.80e-01 |
| ASD PRS | MA v FC | 1.015 | 0.977,1.055 | 0.019 | 4.33e-01 |
| ASD PRS | MA v AR | 0.991 | 0.956,1.027 | 0.018 | 6.21e-01 |
| Anxiety PRS | MA v IR | 1.002 | 0.978,1.028 | 0.013 | 8.53e-01 |
| Anxiety PRS | MA v EA | 0.978 | 0.919,1.04 | 0.032 | 4.79e-01 |
| Anxiety PRS | MA v FC | 1.000 | 0.963,1.038 | 0.019 | 9.91e-01 |
| Anxiety PRS | MA v AR | 1.014 | 0.978,1.05 | 0.018 | 4.55e-01 |

Table 27: Associations of polygenic risk scores (PRS) of six disorders with most likely class membership weighted for the probability of inclusion in each class.

| Response | Class comparison | Effect | 95% CI | SE | P-value |
|---|---|---|---|---|---|
| Bipolar disorder PRS | IR v EA | 1.172 | 1.101,1.248 | 0.032 | 7.86e-07 |
| Bipolar disorder PRS | IR v MA | 1.000 | 0.975,1.026 | 0.013 | 9.90e-01 |
| Bipolar disorder PRS | IR v FC | 1.078 | 1.037,1.122 | 0.020 | 1.75e-04 |
| Bipolar disorder PRS | IR v AR | 1.014 | 0.978,1.053 | 0.019 | 4.47e-01 |
| Schizophrenia PRS | IR v EA | 1.256 | 1.179,1.339 | 0.032 | 2.16e-12 |
| Schizophrenia PRS | IR v MA | 1.008 | 0.983,1.034 | 0.013 | 5.17e-01 |
| Schizophrenia PRS | IR v FC | 1.118 | 1.075,1.164 | 0.020 | 3.12e-08 |
| Schizophrenia PRS | IR v AR | 1.074 | 1.034,1.114 | 0.019 | 1.82e-04 |
| Depression PRS | IR v EA | 1.119 | 1.051,1.192 | 0.032 | 4.30e-04 |
| Depression PRS | IR v MA | 0.978 | 0.954,1.003 | 0.013 | 8.69e-02 |
| Depression PRS | IR v FC | 1.030 | 0.991,1.071 | 0.020 | 1.37e-01 |
| Depression PRS | IR v AR | 1.022 | 0.986,1.061 | 0.019 | 2.36e-01 |
| ADHD PRS | IR v EA | 1.035 | 0.972,1.102 | 0.032 | 2.81e-01 |
| ADHD PRS | IR v MA | 0.973 | 0.949,0.998 | 0.013 | 3.21e-02 |
| ADHD PRS | IR v FC | 0.982 | 0.945,1.022 | 0.020 | 3.76e-01 |
| ADHD PRS | IR v AR | 1.022 | 0.985,1.061 | 0.019 | 2.39e-01 |
| ASD PRS | IR v EA | 1.013 | 0.951,1.079 | 0.032 | 6.95e-01 |
| ASD PRS | IR v MA | 1.013 | 0.988,1.038 | 0.013 | 3.25e-01 |
| ASD PRS | IR v FC | 1.028 | 0.988,1.07 | 0.020 | 1.66e-01 |
| ASD PRS | IR v AR | 1.004 | 0.967,1.041 | 0.019 | 8.47e-01 |
| Anxiety PRS | IR v EA | 0.976 | 0.916,1.039 | 0.032 | 4.39e-01 |
| Anxiety PRS | IR v MA | 0.998 | 0.973,1.023 | 0.013 | 8.51e-01 |
| Anxiety PRS | IR v FC | 0.997 | 0.959,1.037 | 0.020 | 8.97e-01 |
| Anxiety PRS | IR v AR | 1.011 | 0.975,1.049 | 0.019 | 5.53e-01 |

Table 28: Comparison of sociodemographic characteristics in the subset of participants in the latent class analysis (LCA subset) with responses to the stem question on ever having experienced a period of manic or irritable mood as well as the subsequent questions of symptoms, and the full cohort participants, in the PROTECT replication study.

| | | LCA subset | | Full cohort | |
|---|---|---|---|---|---|
| | | N | percent | N | percent |
| Total | | 4445 | | 23930 | |
| Sex | Female | 3282 | 74 | 17526 | 73 |
| | Male | 1147 | 26 | 6338 | 26 |
| | NA/Missing | 16 | 0.36 | 66 | 0.28 |
| Education | University degree | 2272 | 51 | 12364 | 52 |
| | College, A Levels NVQ3 or similar | 527 | 12 | 2788 | 12 |
| | Secondary education, GCSE or O Levels | 708 | 16 | 3955 | 17 |
| | Vocational, BTEC or similar | 922 | 21 | 4757 | 20 |
| | NA/ Missing | 16 | 0.36 | 66 | 0.28 |
| Alcohol consumption | Weekly | 2789 | 63 | 15144 | 63 |
| | Occasionally | 1292 | 29 | 6295 | 26 |
| | Never | 336 | 7.6 | 1587 | 6.6 |
| | NA/Missing | 28 | 0.63 | 904 | 3.8 |
| Smoking | Current | 221 | 5 | 874 | 3.7 |
| | Past | 2087 | 47 | 10103 | 42 |
| | Never | 2109 | 47 | 12049 | 50 |
| | NA/Missing | 28 | 0.63 | 904 | 3.8 |
| | | Mean | SD | Mean | SD |
| Age | | 60.9 | 7.1 | 62.3 | 7.4 |
| | NA/ Missing | 16 | | 66 | |

Table 29: Numbers of responses to individual symptoms questions out of a total sample of N=4445 in PROTECT

| Response | Value | N (percent) |
|---|---|---|
| More active | No | 3664 (0.82) |
| More active | Yes | 781 (0.18) |
| More confident | No | 3934 (0.89) |
| More confident | Yes | 511 (0.11) |
| Easily distracted | No | 3384 (0.76) |
| Easily distracted | Yes | 1061 (0.24) |
| More creative | No | 3901 (0.88) |
| More creative | Yes | 544 (0.12) |
| Less sleep | No | 3938 (0.89) |
| Less sleep | Yes | 507 (0.11) |
| Thoughts racing | No | 3139 (0.71) |
| Thoughts racing | Yes | 1306 (0.29) |
| More restless | No | 3076 (0.69) |
| More restless | Yes | 1369 (0.31) |
| More talkative | No | 3653 (0.82) |
| More talkative | Yes | 792 (0.18) |

Table 30: Fit statistics for latent class models with 2:7 classes in PROTECT

| Number of classes | 2 | 3 | 4 | 5 | 6 | 7 |
|---|---|---|---|---|---|---|
| BIC | 29026.1 | 28121.7 | 27871.6 | 27853.9 | 27905.4 | 27964.2 |
| AIC | 28917.3 | 27955.3 | 27647.6 | 27572.3 | 27566.2 | 27567.4 |
| loglikelihood | -14441.639 | -13951.652 | -13788.8 | -13742.16 | -13730.123 | -13721.694 |
| N iterations | 62 | 129 | 234 | 1272 | 7770 | 1556 |
| Entropy | 3.27 | 3.15 | 3.11 | 3.1 | 3.09 | 3.09 |
| Relative entropy | 0.82 | 0.709 | 0.708 | 0.67 | 0.669 | 0.649 |

Table 31: Distribution of responses to manic or irritable stem question by most likely class membership in the optimum 5-class model in PROTECT

| Class | Response | N (percent) |
|---|---|---|
| IR | Ever manic | 9 (1.53) |
| IR | Ever irritable | 557 (94.57) |
| IR | Ever manic and irritable | 23 (3.9) |
| EA | Ever manic | 50 (24.63) |
| EA | Ever irritable | 46 (22.66) |
| EA | Ever manic and irritable | 107 (52.71) |
| MA | Ever manic | 46 (1.67) |
| MA | Ever irritable | 2649 (96.29) |
| MA | Ever manic and irritable | 56 (2.04) |
| FC | Ever manic | 95 (24.61) |
| FC | Ever irritable | 171 (44.3) |
| FC | Ever manic and irritable | 120 (31.09) |
| AR | Ever manic | 47 (9.11) |
| AR | Ever irritable | 384 (74.42) |
| AR | Ever manic and irritable | 85 (16.47) |

Table 32: Distribution of responses to the episode duration question by most likely class membership in the optimum 5-class model in PROTECT

| Class | Response | N (percent) |
|---|---|---|
| IR | 1 - Brief | 189 (32.088) |
| IR | 2 - Moderate | 230 (39.049) |
| IR | 3 - Extended | 119 (20.204) |
| EA | 1 - Brief | 28 (13.793) |
| EA | 2 - Moderate | 72 (35.468) |
| EA | 3 - Extended | 87 (42.857) |
| MA | 1 - Brief | 1203 (43.73) |
| MA | 2 - Moderate | 596 (21.665) |
| MA | 3 - Extended | 345 (12.541) |
| FC | 1 - Brief | 113 (29.275) |
| FC | 2 - Moderate | 142 (36.788) |
| FC | 3 - Extended | 103 (26.684) |
| AR | 1 - Brief | 174 (33.721) |
| AR | 2 - Moderate | 198 (38.372) |
| AR | 3 - Extended | 107 (20.736) |

Table 33: Distribution of responses to the episode disruptiveness question by most likely class membership in the optimum 5-class model in PROTECT

| Class | Response | N (percent) |
|---|---|---|
| IR | Disruptive | 209 (46.04) |
| IR | Not disruptive | 245 (53.96) |
| IR | Missing - NA | 135 (NA) |
| EA | Disruptive | 97 (56.73) |
| EA | Not disruptive | 74 (43.27) |
| EA | Missing - NA | 32 (NA) |
| MA | Disruptive | 502 (25.97) |
| MA | Not disruptive | 1431 (74.03) |
| MA | Missing - NA | 818 (NA) |
| FC | Disruptive | 87 (26.52) |
| FC | Not disruptive | 241 (73.48) |
| FC | Missing - NA | 58 (NA) |
| AR | Disruptive | 146 (36.14) |
| AR | Not disruptive | 258 (63.86) |
| AR | Missing - NA | 112 (NA) |

Table 34: Associations of episode duration with most likely class membership weighted for the probability of inclusion in each class in PROTECT.

| Response | Class comparison | Effect | 95% CI | SE | P-value |
|---|---|---|---|---|---|
| Brief duration mania or irritability | MA v IR | 0.525 | 0.44,0.627 | 0.091 | 1.16e-12 |
| Brief duration mania or irritability | MA v EA | 0.140 | 0.093,0.211 | 0.207 | 2.43e-21 |
| Brief duration mania or irritability | MA v FC | 0.389 | 0.308,0.489 | 0.118 | 1.00e-15 |
| Brief duration mania or irritability | MA v AR | 0.456 | 0.372,0.559 | 0.104 | 3.57e-14 |
| Moderate duration mania or irritability | MA v IR | 1.291 | 1.178,1.415 | 0.047 | 4.46e-08 |
| Moderate duration mania or irritability | MA v EA | 1.264 | 1.083,1.474 | 0.079 | 2.94e-03 |
| Moderate duration mania or irritability | MA v FC | 1.300 | 1.161,1.457 | 0.058 | 6.03e-06 |
| Moderate duration mania or irritability | MA v AR | 1.326 | 1.197,1.469 | 0.052 | 6.30e-08 |
| Extended duration mania or irritability | MA v IR | 1.109 | 1.03,1.194 | 0.038 | 6.08e-03 |
| Extended duration mania or irritability | MA v EA | 1.659 | 1.496,1.839 | 0.053 | 6.19e-22 |
| Extended duration mania or irritability | MA v FC | 1.252 | 1.15,1.364 | 0.043 | 2.14e-07 |
| Extended duration mania or irritability | MA v AR | 1.154 | 1.065,1.251 | 0.041 | 4.99e-04 |
| Brief duration mania or irritability | IR v EA | 0.267 | 0.175,0.408 | 0.217 | 1.08e-09 |
| Brief duration mania or irritability | IR v MA | 1.903 | 1.594,2.273 | 0.091 | 1.20e-12 |
| Brief duration mania or irritability | IR v FC | 0.739 | 0.569,0.961 | 0.134 | 2.41e-02 |
| Brief duration mania or irritability | IR v AR | 0.868 | 0.684,1.102 | 0.122 | 2.46e-01 |
| Moderate duration mania or irritability | IR v EA | 0.979 | 0.83,1.155 | 0.084 | 8.01e-01 |
| Moderate duration mania or irritability | IR v MA | 0.775 | 0.707,0.849 | 0.047 | 4.46e-08 |
| Moderate duration mania or irritability | IR v FC | 1.007 | 0.886,1.145 | 0.065 | 9.12e-01 |
| Moderate duration mania or irritability | IR v AR | 1.027 | 0.913,1.156 | 0.060 | 6.56e-01 |
| Extended duration mania or irritability | IR v EA | 1.496 | 1.336,1.676 | 0.058 | 3.13e-12 |
| Extended duration mania or irritability | IR v MA | 0.902 | 0.838,0.971 | 0.038 | 6.07e-03 |
| Extended duration mania or irritability | IR v FC | 1.130 | 1.025,1.245 | 0.050 | 1.41e-02 |
| Extended duration mania or irritability | IR v AR | 1.041 | 0.948,1.143 | 0.048 | 4.01e-01 |

Table 35: Associations of episode disruptiveness with most likely class membership weighted for the probability of inclusion in each class in PROTECT.

| Response | Class comparison | Effect | 95% CI | SE | P-value |
|---|---|---|---|---|---|
| Problematic mania or irritability | MA v IR | 2.092 | 1.718,2.547 | 0.100 | 1.97e-13 |
| Problematic mania or irritability | MA v EA | 3.596 | 2.615,4.944 | 0.162 | 3.32e-15 |
| Problematic mania or irritability | MA v FC | 1.055 | 0.813,1.368 | 0.133 | 6.89e-01 |
| Problematic mania or irritability | MA v AR | 1.743 | 1.394,2.18 | 0.114 | 1.11e-06 |
| Problematic mania or irritability | IR v EA | 1.719 | 1.221,2.421 | 0.175 | 1.92e-03 |
| Problematic mania or irritability | IR v MA | 0.478 | 0.393,0.582 | 0.100 | 1.97e-13 |
| Problematic mania or irritability | IR v FC | 0.504 | 0.378,0.673 | 0.148 | 3.47e-06 |
| Problematic mania or irritability | IR v AR | 0.833 | 0.645,1.077 | 0.131 | 1.63e-01 |

Table 36: Associations of sex with most likely class membership weighted for the probability of inclusion in each class in PROTECT.

| Response | Class comparison | Effect | 95% CI | SE | P-value |
|---|---|---|---|---|---|
| Sex | MA v IR | 1.098 | 0.91,1.325 | 0.096 | 3.27e-01 |
| Sex | MA v EA | 1.473 | 1.081,2.007 | 0.158 | 1.41e-02 |
| Sex | MA v FC | 1.671 | 1.337,2.09 | 0.114 | 6.64e-06 |
| Sex | MA v AR | 1.103 | 0.892,1.366 | 0.109 | 3.66e-01 |
| Sex | IR v EA | 1.343 | 0.958,1.882 | 0.172 | 0.0874 |
| Sex | IR v MA | 0.911 | 0.755,1.099 | 0.096 | 0.3309 |
| Sex | IR v FC | 1.523 | 1.173,1.978 | 0.133 | 0.0016 |
| Sex | IR v AR | 1.006 | 0.781,1.295 | 0.129 | 0.9651 |

Table 37: Associations of education levels with most likely class membership weighted for the probability of inclusion in each class in PROTECT.

| Response | Class comparison | Effect | 95% CI | SE | P-value |
|---|---|---|---|---|---|
| College A Levels NVQ3 or below or similar | MA v IR | 1.209 | 0.948,1.542 | 0.124 | 1.27e-01 |
| College A Levels NVQ3 or below or similar | MA v EA | 0.948 | 0.599,1.502 | 0.234 | 8.21e-01 |
| College A Levels NVQ3 or below or similar | MA v FC | 0.968 | 0.694,1.35 | 0.170 | 8.48e-01 |
| College A Levels NVQ3 or below or similar | MA v AR | 1.159 | 0.876,1.534 | 0.143 | 3.00e-01 |
| Secondary education GCSE or O Levels | MA v IR | 1.475 | 1.196,1.819 | 0.107 | 2.75e-04 |
| Secondary education GCSE or O Levels | MA v EA | 0.690 | 0.435,1.093 | 0.235 | 1.13e-01 |
| Secondary education GCSE or O Levels | MA v FC | 1.107 | 0.832,1.475 | 0.146 | 4.85e-01 |
| Secondary education GCSE or O Levels | MA v AR | 1.282 | 1.003,1.639 | 0.125 | 4.68e-02 |
| University degree | MA v IR | 0.664 | 0.563,0.782 | 0.084 | 1.02e-06 |
| University degree | MA v EA | 1.299 | 0.97,1.738 | 0.149 | 7.88e-02 |
| University degree | MA v FC | 1.011 | 0.82,1.247 | 0.107 | 9.19e-01 |
| University degree | MA v AR | 0.834 | 0.693,1.005 | 0.095 | 5.62e-02 |
| Vocational or BTEC or similar | MA v IR | 1.163 | 0.957,1.414 | 0.100 | 1.30e-01 |
| Vocational or BTEC or similar | MA v EA | 0.892 | 0.619,1.286 | 0.186 | 5.42e-01 |
| Vocational or BTEC or similar | MA v FC | 0.924 | 0.71,1.203 | 0.135 | 5.57e-01 |
| Vocational or BTEC or similar | MA v AR | 0.973 | 0.772,1.226 | 0.118 | 8.15e-01 |
| College A Levels NVQ3 or below or similar | IR v EA | 0.785 | 0.481,1.282 | 0.250 | 3.34e-01 |
| College A Levels NVQ3 or below or similar | IR v MA | 0.827 | 0.649,1.056 | 0.124 | 1.27e-01 |
| College A Levels NVQ3 or below or similar | IR v FC | 0.801 | 0.551,1.164 | 0.191 | 2.44e-01 |
| College A Levels NVQ3 or below or similar | IR v AR | 0.959 | 0.692,1.331 | 0.167 | 8.04e-01 |
| Secondary education GCSE or O Levels | IR v EA | 0.467 | 0.289,0.755 | 0.245 | 1.91e-03 |
| Secondary education GCSE or O Levels | IR v MA | 0.678 | 0.55,0.836 | 0.107 | 2.75e-04 |
| Secondary education GCSE or O Levels | IR v FC | 0.750 | 0.546,1.032 | 0.163 | 7.72e-02 |
| Secondary education GCSE or O Levels | IR v AR | 0.868 | 0.655,1.152 | 0.144 | 3.27e-01 |
| University degree | IR v EA | 1.957 | 1.427,2.683 | 0.161 | 3.08e-05 |
| University degree | IR v MA | 1.507 | 1.278,1.776 | 0.084 | 1.03e-06 |
| University degree | IR v FC | 1.523 | 1.196,1.941 | 0.124 | 6.59e-04 |
| University degree | IR v AR | 1.257 | 1.007,1.57 | 0.113 | 4.34e-02 |
| Vocational or BTEC or similar | IR v EA | 0.767 | 0.519,1.134 | 0.200 | 1.84e-01 |
| Vocational or BTEC or similar | IR v MA | 0.860 | 0.707,1.045 | 0.100 | 1.30e-01 |
| Vocational or BTEC or similar | IR v FC | 0.795 | 0.59,1.071 | 0.152 | 1.31e-01 |
| Vocational or BTEC or similar | IR v AR | 0.836 | 0.638,1.096 | 0.138 | 1.95e-01 |

Table 38: Associations of alcohol consumption frequency with most likely class membership weighted for the probability of inclusion in each class in PROTECT.

| Response | Class comparison | Effect | 95% CI | SE | P-value |
|---|---|---|---|---|---|
| Never | MA v IR | 1.037 | 0.753,1.428 | 0.163 | 0.82539 |
| Never | MA v EA | 1.531 | 0.944,2.482 | 0.247 | 0.08430 |
| Never | MA v FC | 1.472 | 1.024,2.114 | 0.185 | 0.03665 |
| Never | MA v AR | 1.470 | 1.062,2.033 | 0.166 | 0.02005 |
| Occasionally | MA v IR | 1.220 | 1.023,1.456 | 0.090 | 0.02719 |
| Occasionally | MA v EA | 1.237 | 0.91,1.681 | 0.157 | 0.17521 |
| Occasionally | MA v FC | 1.029 | 0.816,1.298 | 0.118 | 0.80945 |
| Occasionally | MA v AR | 1.158 | 0.946,1.417 | 0.103 | 0.15579 |
| Weekly | MA v IR | 0.827 | 0.699,0.979 | 0.086 | 0.02701 |
| Weekly | MA v EA | 0.721 | 0.539,0.964 | 0.148 | 0.02709 |
| Weekly | MA v FC | 0.860 | 0.693,1.067 | 0.110 | 0.17026 |
| Weekly | MA v AR | 0.776 | 0.642,0.939 | 0.097 | 0.00909 |
| Never | IR v EA | 1.479 | 0.865,2.528 | 0.273 | 0.1523 |
| Never | IR v MA | 0.963 | 0.699,1.326 | 0.163 | 0.8170 |
| Never | IR v FC | 1.422 | 0.924,2.186 | 0.220 | 0.1092 |
| Never | IR v AR | 1.418 | 0.951,2.113 | 0.204 | 0.0867 |
| Occasionally | IR v EA | 1.014 | 0.727,1.413 | 0.169 | 0.9359 |
| Occasionally | IR v MA | 0.820 | 0.687,0.978 | 0.090 | 0.0274 |
| Occasionally | IR v FC | 0.843 | 0.648,1.098 | 0.135 | 0.2059 |
| Occasionally | IR v AR | 0.949 | 0.748,1.205 | 0.122 | 0.6700 |
| Weekly | IR v EA | 0.871 | 0.636,1.194 | 0.161 | 0.3916 |
| Weekly | IR v MA | 1.209 | 1.022,1.43 | 0.086 | 0.0270 |
| Weekly | IR v FC | 1.040 | 0.811,1.332 | 0.126 | 0.7589 |
| Weekly | IR v AR | 0.938 | 0.749,1.176 | 0.115 | 0.5811 |

Table 39: Associations of smoking with most likely class membership weighted for the probability of inclusion in each class in PROTECT.

| Response | Class comparison | Effect | 95% CI | SE | P-value |
|---|---|---|---|---|---|
| Current | MA v IR | 1.145 | 0.763,1.72 | 0.207 | 5.13e-01 |
| Current | MA v EA | 3.403 | 2.122,5.456 | 0.241 | 3.70e-07 |
| Current | MA v FC | 2.383 | 1.595,3.56 | 0.205 | 2.23e-05 |
| Current | MA v AR | 1.679 | 1.12,2.515 | 0.206 | 1.20e-02 |
| Never | MA v IR | 1.017 | 0.864,1.198 | 0.083 | 8.36e-01 |
| Never | MA v EA | 0.622 | 0.463,0.835 | 0.150 | 1.58e-03 |
| Never | MA v FC | 0.731 | 0.591,0.903 | 0.108 | 3.70e-03 |
| Never | MA v AR | 0.921 | 0.764,1.109 | 0.095 | 3.85e-01 |
| Past | MA v IR | 0.962 | 0.817,1.133 | 0.083 | 6.44e-01 |
| Past | MA v EA | 1.153 | 0.866,1.535 | 0.146 | 3.30e-01 |
| Past | MA v FC | 1.125 | 0.912,1.386 | 0.107 | 2.71e-01 |
| Past | MA v AR | 0.986 | 0.818,1.188 | 0.095 | 8.80e-01 |
| Current | IR v EA | 2.971 | 1.711,5.159 | 0.282 | 0.000111 |
| Current | IR v MA | 0.874 | 0.582,1.312 | 0.207 | 0.514611 |
| Current | IR v FC | 2.084 | 1.273,3.41 | 0.251 | 0.003495 |
| Current | IR v AR | 1.466 | 0.893,2.405 | 0.253 | 0.130199 |
| Never | IR v EA | 0.611 | 0.445,0.84 | 0.162 | 0.002421 |
| Never | IR v MA | 0.983 | 0.835,1.157 | 0.083 | 0.835101 |
| Never | IR v FC | 0.718 | 0.563,0.916 | 0.124 | 0.007692 |
| Never | IR v AR | 0.905 | 0.725,1.129 | 0.113 | 0.376405 |
| Past | IR v EA | 1.198 | 0.878,1.634 | 0.158 | 0.253981 |
| Past | IR v MA | 1.039 | 0.882,1.224 | 0.083 | 0.644564 |
| Past | IR v FC | 1.169 | 0.918,1.488 | 0.123 | 0.205004 |
| Past | IR v AR | 1.024 | 0.821,1.279 | 0.113 | 0.830648 |

Table 40: Numbers of cases of self-reported diagnoses of six disorders by most likely class membership in PROTECT.

| Class | Outcome | Response | N |
|---|---|---|---|
| IR | GAD | Case | 200 |
| EA | GAD | Case | 69 |
| MA | GAD | Case | 515 |
| FC | GAD | Case | 129 |
| AR | GAD | Case | 167 |
| IR | Depression | Case | 322 |
| EA | Depression | Case | 133 |
| MA | Depression | Case | 1022 |
| FC | Depression | Case | 222 |
| AR | Depression | Case | 278 |
| IR | Mania or bipolar | Case | 5 |
| EA | Mania or bipolar | Case | 63 |
| MA | Mania or bipolar | Case | 8 |
| FC | Mania or bipolar | Case | 23 |
| AR | Mania or bipolar | Case | 17 |
| IR | Schizophrenia or psychosis | Case | 2 |
| EA | Schizophrenia or psychosis | Case | 19 |
| MA | Schizophrenia or psychosis | Case | 6 |
| FC | Schizophrenia or psychosis | Case | 10 |
| AR | Schizophrenia or psychosis | Case | 8 |

Table 41: Associations of self-reported diagnoses of three disorders with most likely class membership weighted for the probability of inclusion in each class in PROTECT.

| Response | Class comparison | Effect | 95% CI | SE | P-value |
|---|---|---|---|---|---|
| Self reported GAD | MA v IR | 1.944 | 1.616,2.337 | 0.094 | 1.59e-12 |
| Self reported GAD | MA v EA | 2.308 | 1.703,3.127 | 0.155 | 6.80e-08 |
| Self reported GAD | MA v FC | 2.101 | 1.67,2.642 | 0.117 | 2.18e-10 |
| Self reported GAD | MA v AR | 2.000 | 1.626,2.46 | 0.106 | 5.37e-11 |
| Self reported Schizophrenia or psychosis | MA v IR | 1.760 | 0.409,7.575 | 0.745 | 4.48e-01 |
| Self reported Schizophrenia or psychosis | MA v EA | 46.605 | 17.466,124.356 | 0.501 | 1.69e-14 |
| Self reported Schizophrenia or psychosis | MA v FC | 12.984 | 4.559,36.978 | 0.534 | 1.58e-06 |
| Self reported Schizophrenia or psychosis | MA v AR | 6.954 | 2.289,21.123 | 0.567 | 6.24e-04 |
| Self reported Depression | MA v IR | 1.721 | 1.46,2.028 | 0.084 | 9.48e-11 |
| Self reported Depression | MA v EA | 3.087 | 2.293,4.156 | 0.152 | 1.09e-13 |
| Self reported Depression | MA v FC | 2.191 | 1.774,2.707 | 0.108 | 3.27e-13 |
| Self reported Depression | MA v AR | 1.892 | 1.569,2.282 | 0.095 | 2.40e-11 |
| Self reported Mania or bipolar disorder | MA v IR | 1.715 | 0.565,5.21 | 0.567 | 3.41e-01 |
| Self reported Mania or bipolar disorder | MA v EA | 111.555 | 54.728,227.39 | 0.363 | 1.69e-38 |
| Self reported Mania or bipolar disorder | MA v FC | 18.352 | 8.603,39.146 | 0.387 | 5.15e-14 |
| Self reported Mania or bipolar disorder | MA v AR | 8.114 | 3.583,18.373 | 0.417 | 5.15e-07 |

Table 42: Associations of PRS of six disorders with most likely class membership weighted for the probability of inclusion in each class in PROTECT.

| Response | Class comparison | Effect | 95% CI | SE | P-value |
|---|---|---|---|---|---|
| Bipolar disorder PRS | MA v IR | 0.976 | 0.844,1.129 | 0.074 | 0.7417 |
| Bipolar disorder PRS | MA v EA | 1.346 | 1.035,1.749 | 0.134 | 0.0264 |
| Bipolar disorder PRS | MA v FC | 1.010 | 0.834,1.222 | 0.098 | 0.9227 |
| Bipolar disorder PRS | MA v AR | 1.072 | 0.909,1.264 | 0.084 | 0.4072 |
| Schizophrenia PRS | MA v IR | 0.870 | 0.753,1.004 | 0.073 | 0.0567 |
| Schizophrenia PRS | MA v EA | 0.985 | 0.758,1.28 | 0.134 | 0.9098 |
| Schizophrenia PRS | MA v FC | 0.913 | 0.755,1.103 | 0.097 | 0.3460 |
| Schizophrenia PRS | MA v AR | 0.932 | 0.791,1.098 | 0.083 | 0.4007 |
| Depression PRS | MA v IR | 1.105 | 0.959,1.274 | 0.072 | 0.1676 |
| Depression PRS | MA v EA | 1.176 | 0.91,1.521 | 0.131 | 0.2157 |
| Depression PRS | MA v FC | 1.114 | 0.924,1.343 | 0.095 | 0.2567 |
| Depression PRS | MA v AR | 1.040 | 0.886,1.222 | 0.082 | 0.6302 |
| ADHD PRS | MA v IR | 1.140 | 0.986,1.318 | 0.074 | 0.0764 |
| ADHD PRS | MA v EA | 1.212 | 0.933,1.575 | 0.134 | 0.1502 |
| ADHD PRS | MA v FC | 0.973 | 0.804,1.177 | 0.097 | 0.7759 |
| ADHD PRS | MA v AR | 1.066 | 0.904,1.256 | 0.084 | 0.4478 |
| ASD PRS | MA v IR | 1.009 | 0.872,1.166 | 0.074 | 0.9078 |
| ASD PRS | MA v EA | 0.784 | 0.603,1.02 | 0.134 | 0.0700 |
| ASD PRS | MA v FC | 0.949 | 0.784,1.149 | 0.098 | 0.5909 |
| ASD PRS | MA v AR | 0.963 | 0.816,1.135 | 0.084 | 0.6514 |
| Anxiety PRS | MA v IR | 0.962 | 0.833,1.111 | 0.074 | 0.5952 |
| Anxiety PRS | MA v EA | 0.948 | 0.73,1.231 | 0.133 | 0.6902 |
| Anxiety PRS | MA v FC | 0.864 | 0.714,1.045 | 0.097 | 0.1326 |
| Anxiety PRS | MA v AR | 0.963 | 0.818,1.134 | 0.083 | 0.6505 |
